# Impact of patient gender on low back pain management before and after the COVID-19 pandemic in commercially insured and Medicare Advantage cohorts. A retrospective cohort study

**DOI:** 10.1101/2023.06.05.23290968

**Authors:** David Elton, Meng Zhang

**Author notes:** Corresponding author: David Elton Optum 11000 Optum Circle, Eden Prairie, MN 55344.

## Abstract

**Background:** Variability in the management of LBP has been extensively studied, however the degree to which this variability is associated with patient gender is less well understood. The purpose of this retrospective cohort study was to examine variability in the management of LBP associated with patient gender in commercially insured (CI) and Medicare Advantage (MA) cohorts before and after the COVID-19 pandemic.

**Methods:** A US national sample of LBP episodes with a duration of less than 91 days experienced during 2019-2021 was analyzed. Independent variables included patient gender, whether an individual had CI or MA coverage, and the timing of LBP onset during pre-, early, and late COVID time periods. Dependent measures included the percent of individuals initially contacting eighteen types of health care provider (HCP) and receiving twenty-two types of health care services, and total episode cost. Measures associated with female patients were compared with a male patient baseline to examine patient gender related differences.

**Results:** The study included 222,043 CI and 466,125 MA complete episodes of LBP. 114,322 (51.5%) of the CI and 281,597 (60.4%) of MA episodes were associated with female patients. Individual home address zip code population attributes were nearly identical in both CI and MA cohorts.

During the pre-, early, and late COVID time periods, in both CI and MA cohorts, female patients were less likely than male patients to initially contact DCs (risk ratio (RR) CI pre-COVID 0.88, CI early COVID 0.90, CI late COVID 0.86, MA pre 0.70, MA early 0.70, MA late 0.73) and were more likely to initially contact rheumatologists (2.72, 2.62, 3.20, 2.15, 2.59, 2.08). In the CI cohort during the pre-, early, and late COVID time periods female patients more likely than male patients to initially contact physical therapists (PT) (RR pre-COVID 1.24, early COVID 1.17, late COVID 1.16) and licensed acupuncturists (LAC) (1.75, 1.53, 2.21).

In both the CI and MA cohorts plain film radiology was the most provided service for both female (32-40% of episodes) and male (31-40%) patients. During all time periods in both CI and MA cohorts female patients were less likely than male patients to receive spinal surgery (risk ratio (RR) CI pre-COVID 0.53, CI early COVID 0.54, CI late COVID 0.53, MA pre-0.45, MA early 0.46, MA late 0.42), prescription oral steroids (0.75, 0.73, 0.77, 0.82, 0.79, 0.83), and chiropractic manipulative therapy (CMT) (0.87, 0.89, 0.85, 0.70, 0.71, 0.73). In the CI cohort during all time periods female patients more likely than male patients to receive acupuncture (RR pre-1.41, early 1.48, late 1.48).

**Conclusions:** In both CI and MA cohorts, and compared to males, females with LBP were less likely to seek treatment from DCs and more likely to seek treatment from Rheumatologists. In the CI cohort females were more likely than males to seek treatment from PTs and LAcs. Females with LBP were less likely than males to undergo spinal surgery, receive a prescription oral steroid, or receive CMT.

## Background

The prevalence, variability, disability, and costs associated with low back pain (LBP) have been thoroughly explored.^1–7^ LBP is associated with almost half of low-value spending, defined as costs incurred with little to no associated benefit.^8, 9^ In an effort to improve the value of LBP management, high-quality clinical practice guidelines (CPG) have been developed.^1, 5, 6^ LBP CPGs emphasize natural history, self-care, and non-pharmaceutical services as first-line approaches.

Addressing disparities in availability and access to healthcare resources, and health outcomes, is an important public health focus.^10–12^ The association between patient-provider gender concordance, and clinical and experience outcomes, has revealed mixed results.^13–19^ Specific to the management of LBP, and musculoskeletal conditions generally, gender differences have been identified in prevalence, care seeking, and management.^20–27^ Females have been found to have higher prevalence of LBP than age matched males.^22, 25^ Only 33% of LBP CPGs incorporate individual gender into diagnosis or treatment recommendations.^23^ Prescription analgesic use for LBP has been found to be similar for females and males.^20^ For LBP, females have reported responding more favorably than males to acupuncture, while males reported responding more favorably than females to spinal manipulative therapy.^15^ While the degree of improvement following spinal surgery has been reported to be similar between females and males, females receive surgery at a more advanced state.^26, 27^

The type of HCP initially contacted, and subsequent care pathways has been utilized to explore variation in management of LBP with PCPs and DCs the most common types of HCP initially contacted by an individual with LBP.^28–30^ The gender distribution among of types of HCP managing LBP is highly variable. Among non-pharmaceutical, non-interventional providers chiropractors (DC)^31^ have the lowest proportion of female providers (32%), while physical therapists (PT)^32^ and licensed acupuncturists (LAc)^33^ are approximately 65% female. Among primary care HCP types, primary care physicians (PCP - 40%)^34^ and Doctors of Osteopathy (DO - 43%)^35^ have the lowest proportion of female practitioners, while nurse practitioners (87%)^36^ and physician assistants (PA - 67%)^37^ have the highest. Among physician specialist HCP types, orthopedic surgeons (OS – 7%), neurosurgeons (NS – 9%), and pain management physicians (PM – 19%) have the lowest proportion of female practitioners, while rheumatologists (46%), physical medicine and rehabilitation physicians (PMR – 36%), and neurologists (31%) have the highest.^34^ Radiologists (26%) and emergency medicine physicians (EM – 28%) also consist of a low proportion of female practitioners.^34^ [Supplement – Provider Type Gender] For individuals with LBP, the degree to which the selection of an initial HCP is influenced by gender concordance has not been investigated.

COVID-19 (COVID), declared a global pandemic on March 11, 2020 by the World Health Organization (WHO)^38–41^, has been associated with variable public policy responses^42–44^ and there are indications that the management of LBP was impacted by COVID.^45^ The degree to which COVID related changes were influenced by the gender of the individual with LBP has not been explored.

The aim of this study was to explore how patient gender is associated with differences in care-seeking for, and the management of, LBP in commercially insured (CI) and Medicare Advantage (MA) cohorts before and after the COVID pandemic. The hypothesis was female patients would prefer seeking care from HCP types with a higher percentage of female providers, and that this would be most evident in HCP types providing hands-on care like DCs, PTs and LAcs.

## Methods

This study’s cohort, methods, and authors are identical to an earlier study of COVID related changes in the management of LBP in CI and MA cohorts.^45^ Combining the earlier and current paper into a single paper was not feasible. The authors of the earlier study have granted permission to use the following Methods description language.

### Study design, population, setting and data sources

This is a retrospective cohort study of individuals seen by one or more HCPs for a complete episode of LBP. Individuals were from a single national insurer administering CI and MA benefit plans. An enrollee database included de-identified enrollment records, and administrative claims data for all inpatient and outpatient services, and pharmacy prescriptions. De-identified in and out-of-network HCP demographic information and professional licensure status were available in an HCP database. ZIP code level 2020 population race and ethnicity data was obtained from the US Census Bureau,^46^ 2019 adjusted gross income (AGI) data from the Internal Revenue Service,^47^ and 2020 socioeconomic Area Deprivation Index (ADI) data from the University of Wisconsin Neighborhood Atlas^®^ database.^48, 49^

With data being de-identified or a Limited Data Set in compliance with the Health Insurance Portability and Accountability Act and customer requirements, the UnitedHealth Group Office of Human Research Affairs Institutional Review Board determined that this study was exempt from ongoing Institutional Review Boards oversight. The study was conducted and reported based on the Strengthening the Reporting of Observational Studies in Epidemiology (STROBE) guidelines [Supplement – STROBE Checklist].^50^

The impact of numerous unmeasurable and unknown confounders, and confounders of measurable hypothesized confounders were likely amplified by the impact of the COVID pandemic and variable public policy responses.^43, 44^ [Supplement – Public Policy] As an alternative to adopting the standard practice of using potentially inadequate approaches such as propensity score matching^51^ to control for available yet incomplete potential confounders derived from administrative claims data to attempt to generate causal insights^52, 53^, the study was designed to address a literature gap of presenting actual, unadjusted associations between individual demographic attributes, HCP selection, and episodic characteristics for the management of LBP during the COVID pandemic. These confounders, significant cohort differences, insurance benefit design differences, and a different distribution of episodes among States resulted in an inability to directly compare CI and MA measures, and no such comparison should be attempted with study data.

### Unit of analysis and cohort selection

Episode of care has been shown to be a valid way to organize administrative claims data to summarize care pathways and analyze the rate and timing of use of services provided for a condition.^30, 54^ The Symmetry^®^ Episode Treatment Groups^®^ (ETG^®^) and Episode Risk Groups^®^ (ERG^®^) version 9.5 methodologies and definitions were used to translate administrative claims data into discrete episodes of care.^55^ A complete episode was defined as having at least 91-day pre- and 61-day post-episode clean periods, during which no services were provided by any HCP for any LBP diagnosis. The episode duration was the number of days between the first and last date of service for an episode.

The cohort consisted of individuals aged 18 years and older with a complete episode of LBP commencing and ending during the calendar years 2019-2021. To align with the timing of the WHO pandemic declaration^38–41^, the pre-COVID period was defined as episodes starting between 3/1/2019 and 2/29/2020, the early COVID period was 3/1/2020 to 2/28/2021, and the late COVID period was 3/1/2021 to 1/28/2022.

Episodes starting in the pre-COVID period had up to 35 months of post-onset duration potential, episodes starting in the early COVID period had up to 23 months and episodes starting in the late COVID period had up to 11 months. The resulting different episode volumes and duration distributions in the pre, early and late COVID periods and within and between the CI and MA cohorts was a potential confounder of a variety of study variables. As one example, MA episode durations were approximately double CI durations indicating a greater prevalence of chronic LBP in the MA cohort. To address this potential study limitation, a sensitivity analysis was conducted to evaluate limiting the cohort to episodes with a duration of less than 61 or less than 91 days. Based on the sensitivity analysis, the study was limited to episodes with a duration of less than 91 days. This approach balanced episode volume across pre, early and late COVID periods and reduced or eliminated the original significant difference in episode duration between pre, early and late COVID periods, between the CI and MI cohorts and between male and female patients. This approach may have partially addressed differing LBP clinical complexity in CI and MA cohorts and reduced but did not eliminate episodes crossing the pre, early and post COVID measurement periods. [Supplement – Episode Duration]

Individuals with diagnoses of malignant and non-malignant neoplasms, fractures and other spinal trauma, infection, congenital deformities and scoliosis, autoimmune disorders, osteoporosis, and advanced arthritis were excluded from the analysis. This approach was used to address potential confounders associated the COVID pandemic impacting care seeking behaviors of individuals with a more complex LBP condition differently from individuals with less complex LBP.

### Variables

Data preprocessing, table generation, and initial analyses were performed in Python (Python Language Reference, Version 3.7.5., n.d.). A goodness-of-fit analysis was performed using D’Agostino’s K-squared test. Non-normally distributed data were reported using the median, interquartile range (IQR), quartile 1 (Q1), and quartile 3 (Q3). Where utilized, p-values do not control for the false discovery rate. Due to the tendency for odds ratios to exaggerate risk in situations where an outcome is relatively common, and as a measure more widely understood in associational analyses, risk ratios (RR) and associated 95% confidence intervals were reported.^56^

The primary independent variables were the individual’s gender, timing of LBP onset and type of insurance coverage. Secondary independent variables were the individual home address 5-digit zip code population percent non-Hispanic white (NHW) and ADI. The primary dependent variables were the percent of individuals with LBP initially contacting one of eighteen types of HCP, and the percent of episodes including twenty-two types of health care services. The main secondary dependent variable was the total cost of care for all reimbursed services provided by any HCP during an episode. Total episode cost included costs associated with all services provided for LBP during an episode, including those not specifically identified in the service categories used in the analyses. Costs for services for which an insurance claim was not submitted, and indirect costs associated with missed days at work or reduced productivity, were not available. Additional secondary dependent variables included episode duration and the number of different HCPs seen during an episode. Due to numerous confounders no attempt was made to calculate the prevalence of LBP during the pre, early and late COVID time periods.

## Results

The study included 688,168 complete episodes of LBP, with 222,043 CI and 466,125 MA episodes. Depending on the pre/early/late COVID measurement period, the CI cohort was 51-52% female with a median age of 45-46, and the MA cohort was 60-61% female with a median age of 72-73. While the scaling for the ERG® Risk Score, a measure of illness burden, was different for CI and MA cohorts, within each cohort the median ERG® Risk Score was higher for females across pre/early/late COVID periods. Within the CI and MA cohorts and for all time-periods population attributes for cohort 5-digit home address zip code were nearly identical for females and males. CI median % NHW population was 66%-67%, ADI was 37-41, and AGI was $74k-$76k. For MA median % NHW was 67%-70%, ADI was 52-54, and AGI was $61k-$63k. Similarly, the number of different HCP seen during an episode and episode duration were similar in CI and MA cohorts, for all time periods and for male and female patients. Within CI and MA cohorts, and for each time-period, male and female patients had similar total episode cost, with both male and female patients experiencing a reduction in total episode cost during the early COVID period. For both male and female patients and for all time periods MA total episode cost was lower than CI. Episodes were from all 50 States; however, this was not a US representative sample and the distribution of episodes among States was different in CI and MA cohorts. [Supplement – States] [Table 1]

**Table 1.**
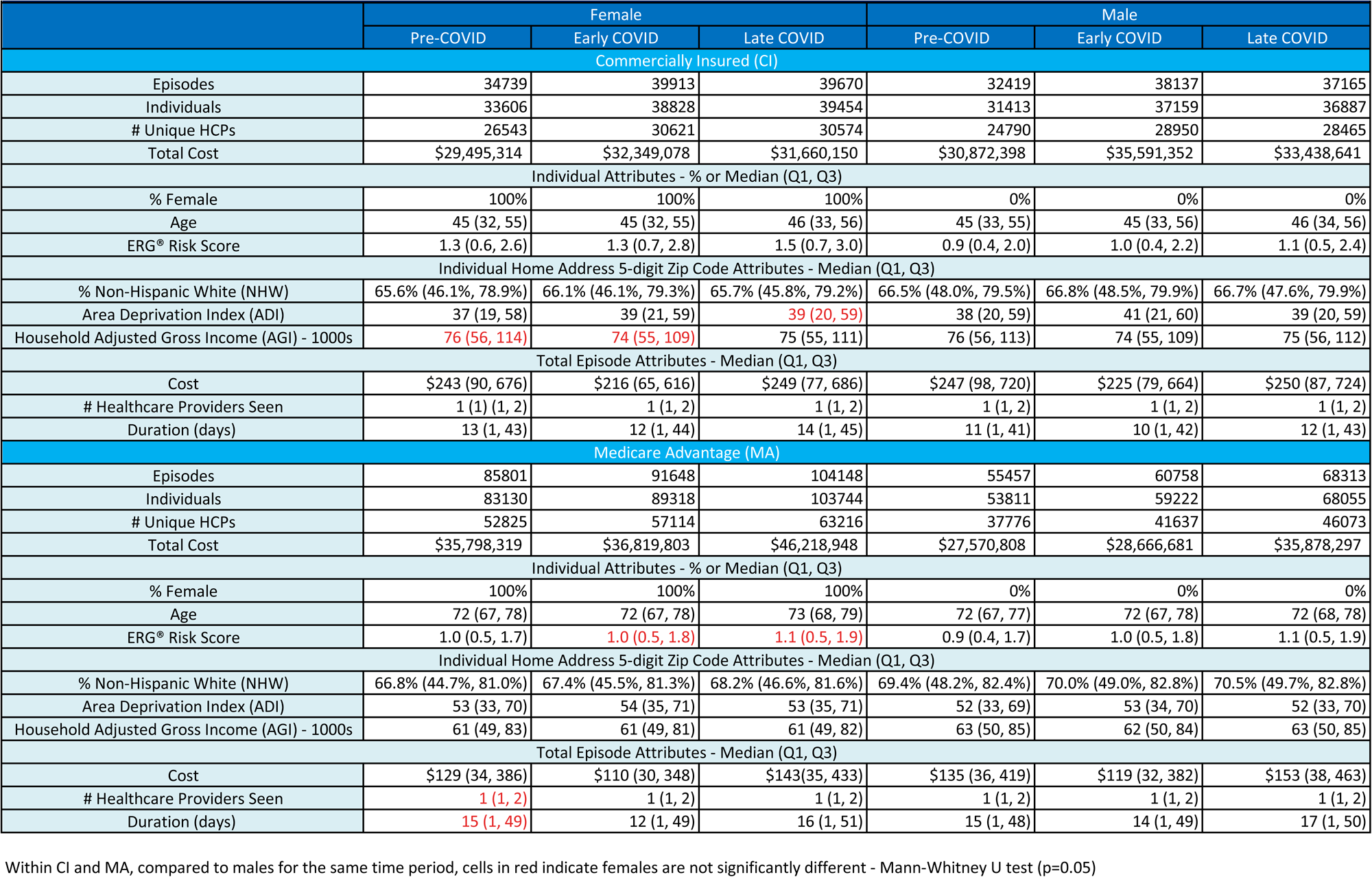
Cohort, population and episode attributes for complete low back pain episodes <91 days duration.

In the CI cohort and during all time periods DCs (24%-28% of episodes) and PCPs (20%-23%) were the types of HCP initially contacted by the highest percent of individuals with LBP. In the MA cohort PCPs (30%-35% of episodes) and Hospitals (14%-17%) were most common. [Table 2] Compared to male patients, in the CI cohort females were less likely to initially contact a DC (RR pre-COVID 0.88, early COVID 0.90, late COVID 0.86) or an Emergency Medicine (EM) physician (0.79, 0.78, 0.87). In the CI cohort females were more likely to initially contact a rheumatologist (2.72, 2.62, 3.20), LAc (1.75, 1.53, 2.21), neurologist (1.58, 1.39, 1.56), or PT (1.24, 1.17, 1.16). [Figure 1] In the MA cohort females were less likely to initially contact a DC (RR pre-COVID 0.70), early COVID 0.70, late COVID 0.73) or neurosurgeon (0.63, 0.71, 0.79). In the MA cohort females were more likely to initially contact a rheumatologist (2.15, 2.59, 2.08). [Figure 2]

**Figure 1.**
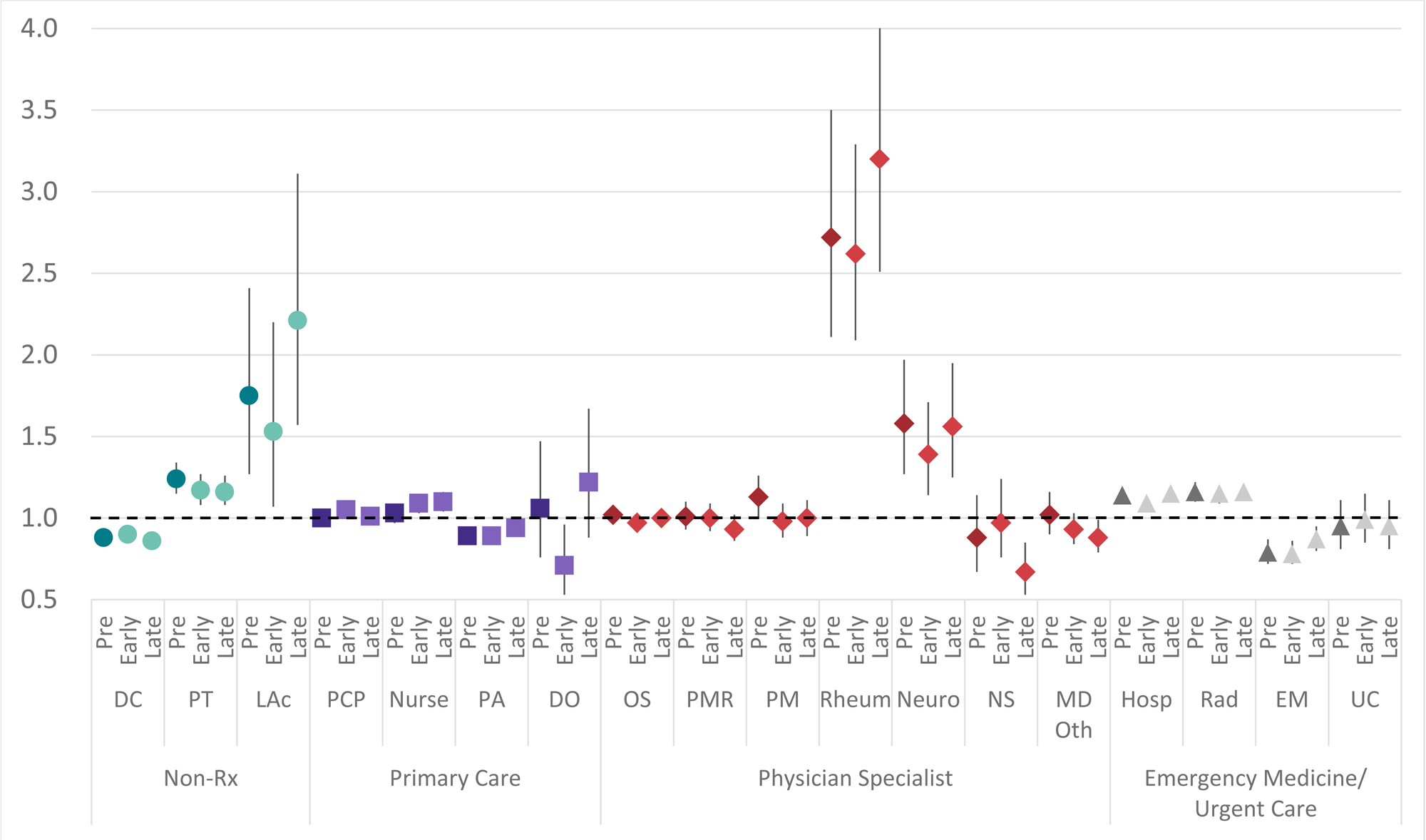
Risk ratio and 95% confidence interval comparing **females to males** for the type of health care provider initially contacted by **commercially insured** individuals with low back pain with episode lasting <91 days Pre=pre-COVID period, Early=0-12 months post-COVID, Late=13-24 months post-COVID, PCP=primary care provider, Nur=nurse, PA=physician assistant, DO=doctor of osteopathy, DC=doctor of chiropractic, PT=physical therapist, LAc=licensed acupuncturist, OS=orthopedic suregon, PMR=physical medicine and rehabilitation, PM=pain managment, Neuro=neurologist, Rheum=rheumatologist, NS=neurosurgeon, Oth=other physician specialist, Hosp=hospital, Rad=radiologist, EM=emergency medicine, UC=urgent care

**Figure 2.**
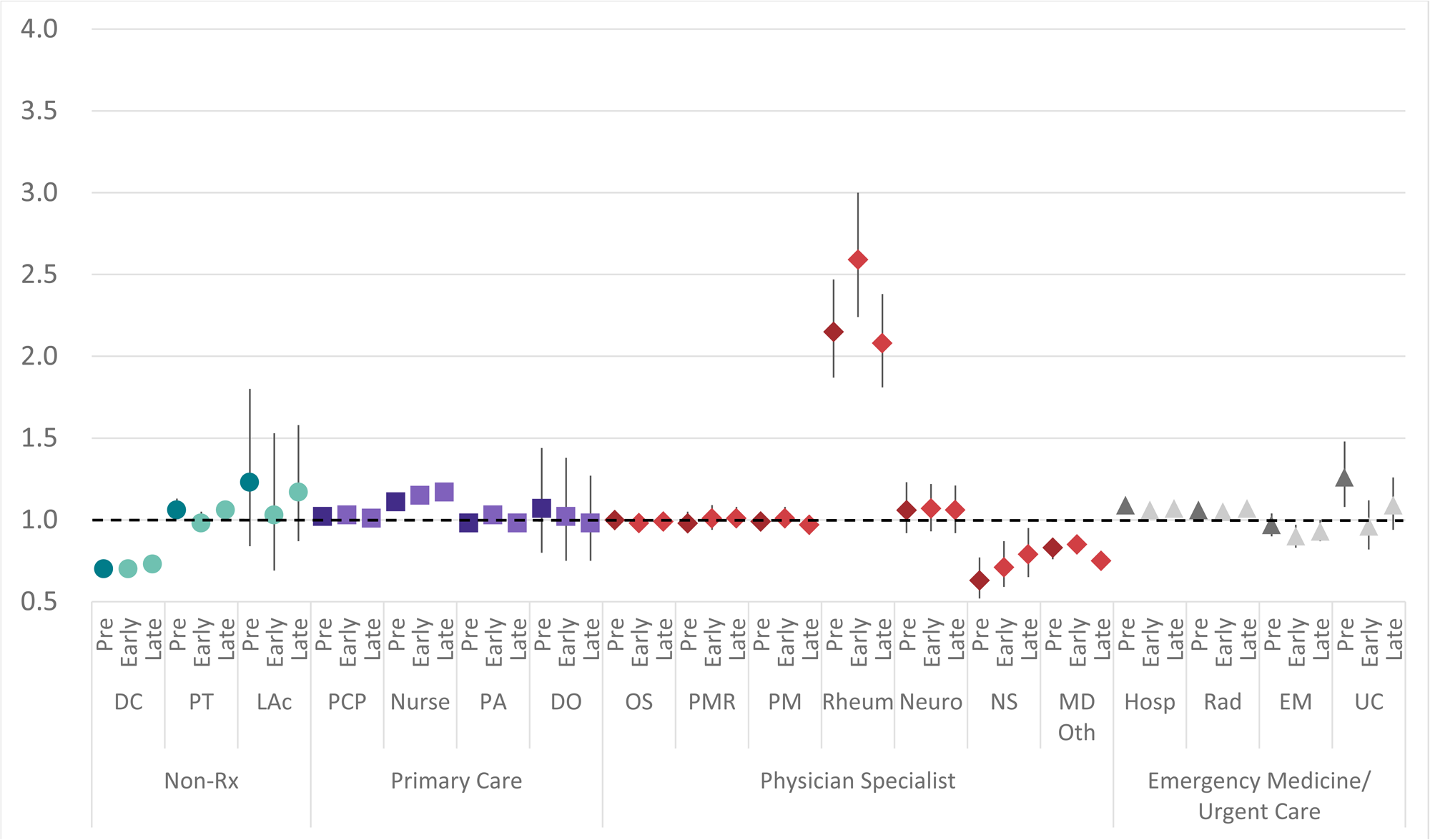
Risk ratio and 95% confidence interval comparing **females to males** for the type of health care provider initially contacted by **medicare advantage** insured individuals with low back pain with episode lasting <91 days Pre=pre-COVID period, Early=0-12 months post-COVID, Late=13-24 months post-COVID, PCP=primary care provider, Nur=nurse, PA=physician assistant, DO=doctor of osteopathy, DC=doctor of chiropractic, PT=physical therapist, LAc=licensed acupuncturist, OS=orthopedic suregon, PMR=physical medicine and rehabilitation, PM=pain managment, Neuro=neurologist, Rheum=rheumatologist, NS=neurosurgeon, Oth=other physician specialist, Hosp=hospital, Rad=radiologist, EM=emergency medicine, UC=urgent care

**Table 2.** Type of healthcare provider initially contacted for low back pain - duration <91 days.

|  |  |  |  |  |  |  |  |  |  |  |
| --- | --- | --- | --- | --- | --- | --- | --- | --- | --- | --- |
|  |  | % of Episodes Where Type of HCP Was Initially Contacted |  |  |  |  |  | Risk Ratio and 95% Confidence Interval Comparing Female to Male |  |  |
|  |  | Pre-COVID |  | Early COVID |  | Late COVID |  | Pre-COVID | Early COVID | Late COVID |
|  |  | F | M | F | M | F | M |  |  |  |
| Commercial Insurance (CI) |  |  |  |  |  |  |  |  |  |  |
| Episodes |  | 34739 | 32419 | 39913 | 38137 | 39670 | 37165 |  |  |  |
| Non-Rx | DC | 25.0% | 28.3% | 25.4% | 28.2% | 23.5% | 27.3% | 0.88 (0.86, 0.90) | 0.90 (0.88, 0.92) | 0.86 (0.84, 0.88) |
|  | PT | 4.2% | 3.4% | 3.4% | 2.9% | 3.6% | 3.1% | 1.24 (1.15, 1.34) | 1.17 (1.08, 1.27) | 1.16 (1.08, 1.26) |
|  | LAc | 0.3% | 0.2% | 0.2% | 0.1% | 0.3% | 0.1% | 1.75 (1.27, 2.41) | 1.53 (1.07, 2.20) | 2.21 (1.57, 3.11) |
| Primary Care | PCP | 20.4% | 20.4% | 22.5% | 21.5% | 19.8% | 19.7% | 1.00 (0.97, 1.03) | 1.05 (1.02, 1.07) | 1.01 (0.98, 1.04) |
|  | Nurse | 5.6% | 5.5% | 6.0% | 5.5% | 5.9% | 5.4% | 1.03 (0.97, 1.09) | 1.09 (1.03, 1.15) | 1.10 (1.04, 1.16) |
|  | PA | 4.8% | 5.4% | 4.5% | 5.0% | 4.7% | 5.0% | 0.89 (0.84, 0.95) | 0.89 (0.84, 0.95) | 0.94 (0.89, 1.00) |
|  | DO | 0.2% | 0.2% | 0.2% | 0.3% | 0.2% | 0.2% | 1.06 (0.76, 1.47) | 0.71 (0.53, 0.96) | 1.22 (0.88, 1.67) |
| Physician Specialist | OS | 8.6% | 8.5% | 8.4% | 8.7% | 8.8% | 8.9% | 1.02 (0.97, 1.07) | 0.97 (0.92, 1.01) | 1.00 (0.95, 1.04) |
|  | PMR | 3.2% | 3.2% | 2.8% | 2.8% | 2.7% | 2.9% | 1.01 (0.93, 1.10) | 1.00 (0.92, 1.09) | 0.93 (0.86, 1.02) |
|  | PM | 1.8% | 1.6% | 1.7% | 1.8% | 1.7% | 1.7% | 1.13 (1.01, 1.26) | 0.98 (0.88, 1.09) | 1.00 (0.89, 1.11) |
|  | Neuro | 0.6% | 0.4% | 0.6% | 0.4% | 0.5% | 0.3% | 1.58 (1.27, 1.97) | 1.39 (1.14, 1.71) | 1.56 (1.25, 1.95) |
|  | Rheum | 0.7% | 0.2% | 0.7% | 0.3% | 0.7% | 0.2% | 2.72 (2.11, 3.50) | 2.62 (2.09, 3.29) | 3.20 (2.51, 4.07) |
|  | NS | 0.3% | 0.4% | 0.3% | 0.3% | 0.3% | 0.4% | 0.88 (0.67, 1.14) | 0.97 (0.76, 1.24) | 0.67 (0.53, 0.85) |
|  | MD Oth | 1.4% | 1.4% | 1.7% | 1.8% | 1.5% | 1.7% | 1.02 (0.90, 1.16) | 0.93 (0.84, 1.03) | 0.88 (0.79, 0.99) |
| Emergency Medicine/<br>Urgent Care | Hosp | 10.9% | 9.5% | 9.9% | 9.1% | 11.7% | 10.2% | 1.14 (1.09, 1.19) | 1.09 (1.04, 1.13) | 1.15 (1.10, 1.19) |
|  | Rad | 8.8% | 7.6% | 8.8% | 7.7% | 10.7% | 9.2% | 1.16 (1.10, 1.22) | 1.15 (1.09, 1.20) | 1.16 (1.11, 1.21) |
|  | EM | 2.3% | 2.9% | 2.2% | 2.8% | 2.4% | 2.8% | 0.79 (0.72, 0.87) | 0.78 (0.72, 0.86) | 0.87 (0.80, 0.95) |
|  | UC | 0.9% | 1.0% | 0.8% | 0.8% | 0.8% | 0.9% | 0.95 (0.81, 1.11) | 0.99 (0.85, 1.15) | 0.95 (0.81, 1.11) |
| Medicare Advantage (MA) |  |  |  |  |  |  |  |  |  |  |
| Episodes |  | 85801 | 55457 | 91648 | 60758 | 104148 | 68313 |  |  |  |
| Non-Rx | DC | 8.2% | 11.7% | 7.4% | 10.5% | 8.0% | 10.9% | 0.70 (0.68, 0.72) | 0.70 (0.68, 0.72) | 0.73 (0.71, 0.75) |
|  | PT | 3.6% | 3.4% | 2.7% | 2.8% | 3.4% | 3.2% | 1.06 (1.01, 1.13) | 0.98 (0.93, 1.05) | 1.06 (1.01, 1.12) |
|  | LAc | 0.1% | 0.1% | 0.1% | 0.1% | 0.1% | 0.1% | 1.23 (0.84, 1.80) | 1.03 (0.69, 1.53) | 1.17 (0.87, 1.58) |
| Primary Care | PCP | 31.8% | 31.1% | 35.3% | 34.4% | 29.7% | 29.3% | 1.02 (1.00, 1.04) | 1.03 (1.01, 1.04) | 1.01 (1.00, 1.03) |
|  | Nurse | 6.4% | 5.7% | 7.0% | 6.1% | 6.6% | 5.6% | 1.11 (1.07, 1.16) | 1.15 (1.10, 1.19) | 1.17 (1.12, 1.21) |
|  | PA | 4.4% | 4.5% | 4.5% | 4.4% | 4.6% | 4.7% | 0.98 (0.94, 1.03) | 1.03 (0.98, 1.08) | 0.98 (0.94, 1.02) |
|  | DO | 0.1% | 0.1% | 0.1% | 0.1% | 0.1% | 0.1% | 1.07 (0.80, 1.44) | 1.02 (0.75, 1.38) | 0.98 (0.75, 1.27) |
| Physician Specialist | OS | 6.5% | 6.4% | 6.3% | 6.4% | 6.7% | 6.8% | 1.00 (0.96, 1.05) | 0.98 (0.95, 1.02) | 0.99 (0.95, 1.02) |
|  | PMR | 2.3% | 2.3% | 2.0% | 2.0% | 2.1% | 2.0% | 0.98 (0.92, 1.05) | 1.01 (0.94, 1.09) | 1.01 (0.95, 1.08) |
|  | PM | 2.9% | 2.9% | 2.7% | 2.6% | 2.5% | 2.6% | 0.99 (0.93, 1.05) | 1.01 (0.95, 1.08) | 0.97 (0.91, 1.03) |
|  | Neuro | 0.5% | 0.5% | 0.6% | 0.6% | 0.5% | 0.5% | 1.06 (0.92, 1.23) | 1.07 (0.93, 1.22) | 1.06 (0.92, 1.21) |
|  | Rheum | 1.0% | 0.5% | 1.0% | 0.4% | 0.8% | 0.4% | 2.15 (1.87, 2.47) | 2.59 (2.24, 3.00) | 2.08 (1.81, 2.38) |
|  | NS | 0.2% | 0.4% | 0.2% | 0.3% | 0.2% | 0.3% | 0.63 (0.52, 0.77) | 0.71 (0.59, 0.87) | 0.79 (0.65, 0.95) |
|  | MD Oth | 1.7% | 2.0% | 2.1% | 2.5% | 1.7% | 2.3% | 0.83 (0.76, 0.89) | 0.85 (0.79, 0.90) | 0.75 (0.70, 0.80) |
| Emergency Medicine/<br>Urgent Care | Hosp | 15.9% | 14.7% | 14.6% | 13.8% | 17.5% | 16.3% | 1.09 (1.06, 1.11) | 1.06 (1.03, 1.09) | 1.07 (1.05, 1.10) |
|  | Rad | 11.9% | 11.2% | 11.4% | 10.9% | 13.2% | 12.4% | 1.06 (1.03, 1.10) | 1.05 (1.02, 1.08) | 1.07 (1.04, 1.09) |
|  | EM | 1.9% | 2.0% | 1.6% | 1.8% | 1.8% | 1.9% | 0.97 (0.90, 1.04) | 0.90 (0.83, 0.97) | 0.93 (0.87, 1.00) |
|  | UC | 0.5% | 0.4% | 0.4% | 0.4% | 0.4% | 0.4% | 1.26 (1.08, 1.48) | 0.96 (0.82, 1.12) | 1.09 (0.94, 1.26) |

Pre=Pre-COVID period, Early=first 12 months post-COVID, Late=13-24 months post-COVID
PCP=primary care physician, PA=physician assistant, DO=doctor of osteopathy, DC=doctor of chiropractic, PT=physical therapist, LAc=licensed acupuncturist, OS=orthopedic surgeon, PMR=physical medicine and rehabilitation, PM=pain management, Neuro=neurologist, Rheum=rheumatologist, NS=neurosurgeon, MD-Oth=other physician specialists, Hosp=hospital, Rad=radiologist, EM=emergency medicine, UC=urgent care
For risk ratio, cells in red indicate the confidence interval includes 1 and likelihood of HCP being initially contacted is not different than the male reference for the same time period

In the CI cohort, during the pre, early, and late COVID periods plain film radiology (36%-40% of episodes), chiropractic manipulative treatment (CMT) (24%-29%), prescription NSAIDs (22%-23%), active care (AC) (20%-23%), and prescription skeletal muscle relaxants (18%-23%) were the only services provided for at least 15% of episodes. In the MA cohort, during the pre, early, and late COVID periods plain film radiology (31%-38% of episodes), prescription NSAIDs (19%-21%), and prescription opioids (15%-17%) were provided during at least 15% of episodes. In the late COVID period MRI was provided for 16%-18% of episodes in the MA cohort. [Table 3] Compared to male patients, in the CI cohort females were less likely to receive spinal surgery (RR pre-COVID 0.53, early COVID 0.53, late COVID 0.53), oral steroids (0.75, 0.73, 0.77), gabapentins (0.79, 0.79, 0.77), chiropractic manipulative therapy (0.87, 0.89, 0.86), passive therapy (0.87, 0.89, 0.85), skeletal muscle relaxants (0.85, 0.88, 0.89), MRI (0.88, 0.88, 0.89), spinal injections (0.87, 0.84, 0.87), or prescription opioids (0.88, 0.90, 0.92). In the CI cohort females were more likely to receive acupuncture (RR pre-COVID 1.41, early COVID 1.48, late COVID 1.48). [Figure 3] In the MA cohort females were less likely to receive initially spinal surgery (RR pre-COVID 0.45, early COVID 0.46, late COVID 0.42), chiropractic manipulative therapy (0.70, 0.71, 0.73), oral steroids (0.82, 0.79, 0.83), MRI (0.87, 0.87, 0.88), skeletal muscle relaxants (0.93, 0.95, 0.91), and spinal injections (0.94, 0.91, 0.94). In the MA cohort females were more likely to receive plain film radiology (RR pre-COVID 1.07, early COVID 1.05, late COVID 1.07) and prescription NSAIDs (1.07, 1.08, 1.07). [Figure 4]

**Figure 3.**
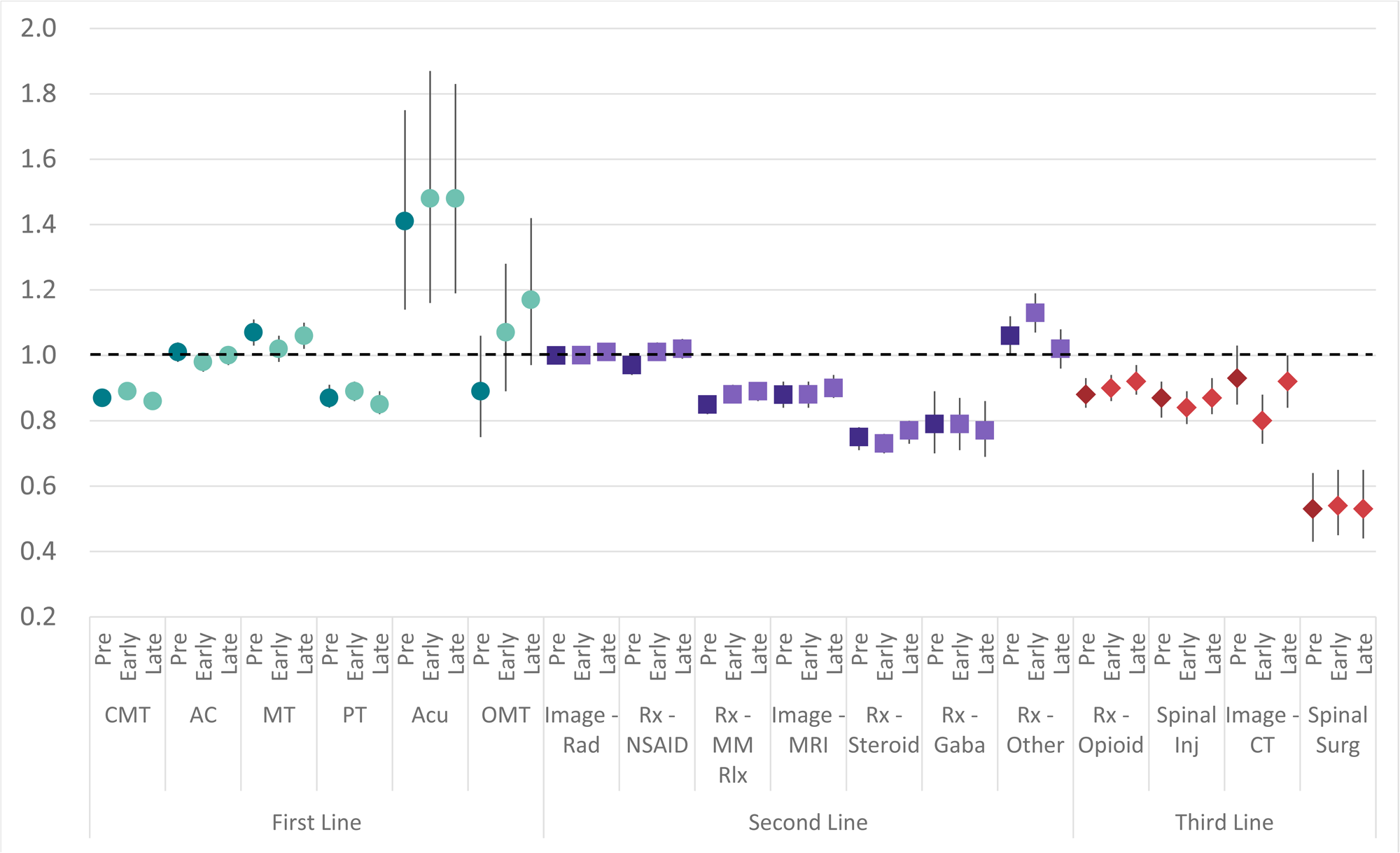
Risk ratio and 95% confidence interval comparing **females to males** for the type of health care services provided for **commercially insured** individuals with low back pain with episode lasting <91 days Pre=pre-COVID period, Early=0-12 months post-COVID, Late=13-24 months post-COVID, CMT=chiropractic manipulative therapy, AC=active care, MT=manual therapy, PT=passive therapy, Acu=acupuncture, OMT=osteopathic manipulative therapy, Imag Rad=radiology, MM Rlx=skeletal muscle relaxant, Imag MRI=MRI scan, Gaba=gabapentin, Oth=other prescription medication, Inj=injection, Imag-CT=CT scan, Surg=surgical procedure

**Figure 4.**
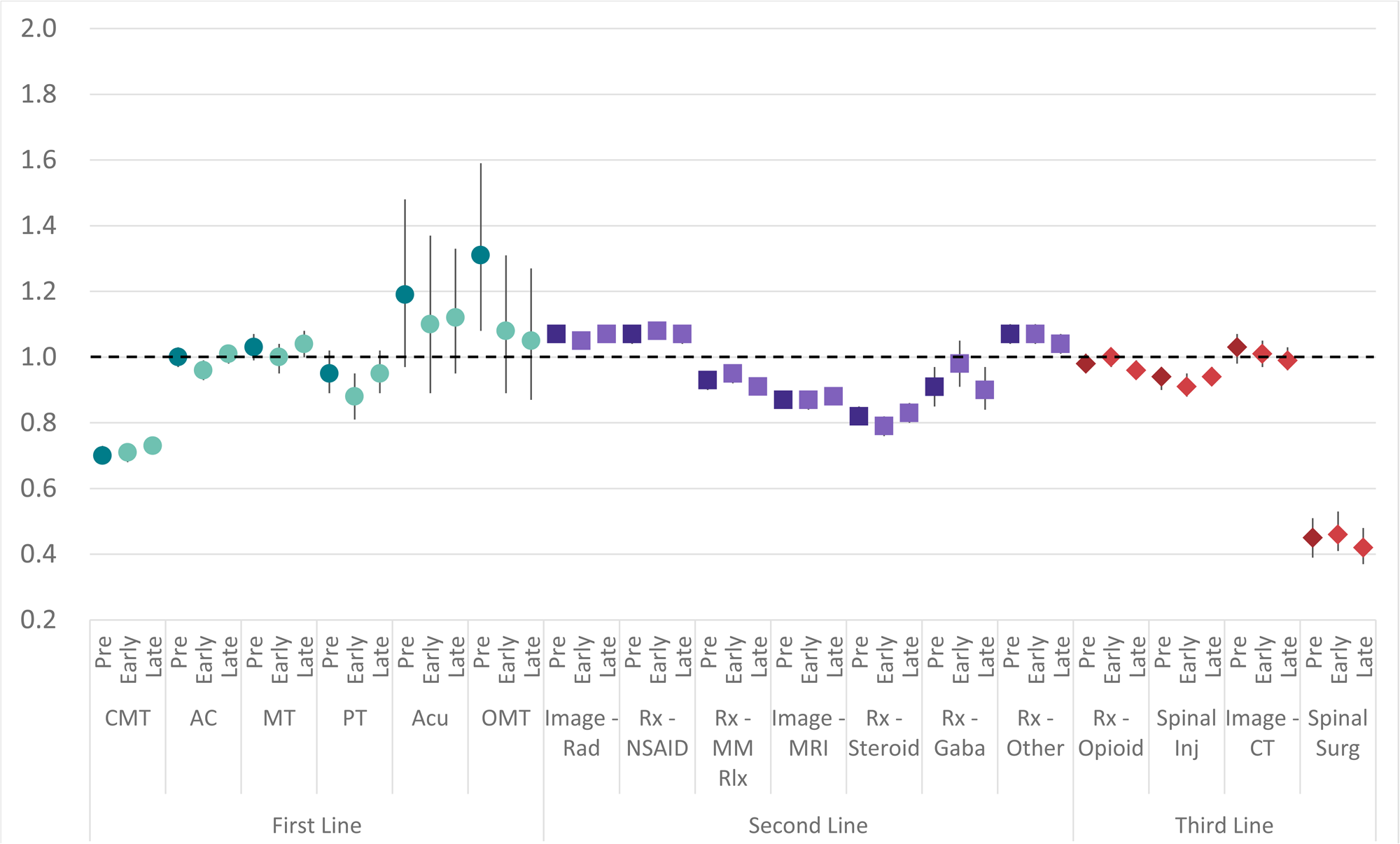
Risk ratio and 95% confidence interval comparing **females to males** for the type of health care services provided for **medicare advantage** insured individuals with low back pain with episode lasting <91 days Pre=pre-COVID period, Early=0-12 months post-COVID, Late=13-24 months post-COVID, CMT=chiropractic manipulative therapy, AC=active care, MT=manual therapy, PT=passive therapy, Acu=acupuncture, OMT=osteopathic manipulative therapy, Imag Rad=radiology, MM Rlx=skeletal muscle relaxant, Imag MRI=MRI scan, Gaba=gabapentin, Oth=other prescription medication, Inj=injection, Imag-CT=CT scan, Surg=surgical procedure

**Table 3.**
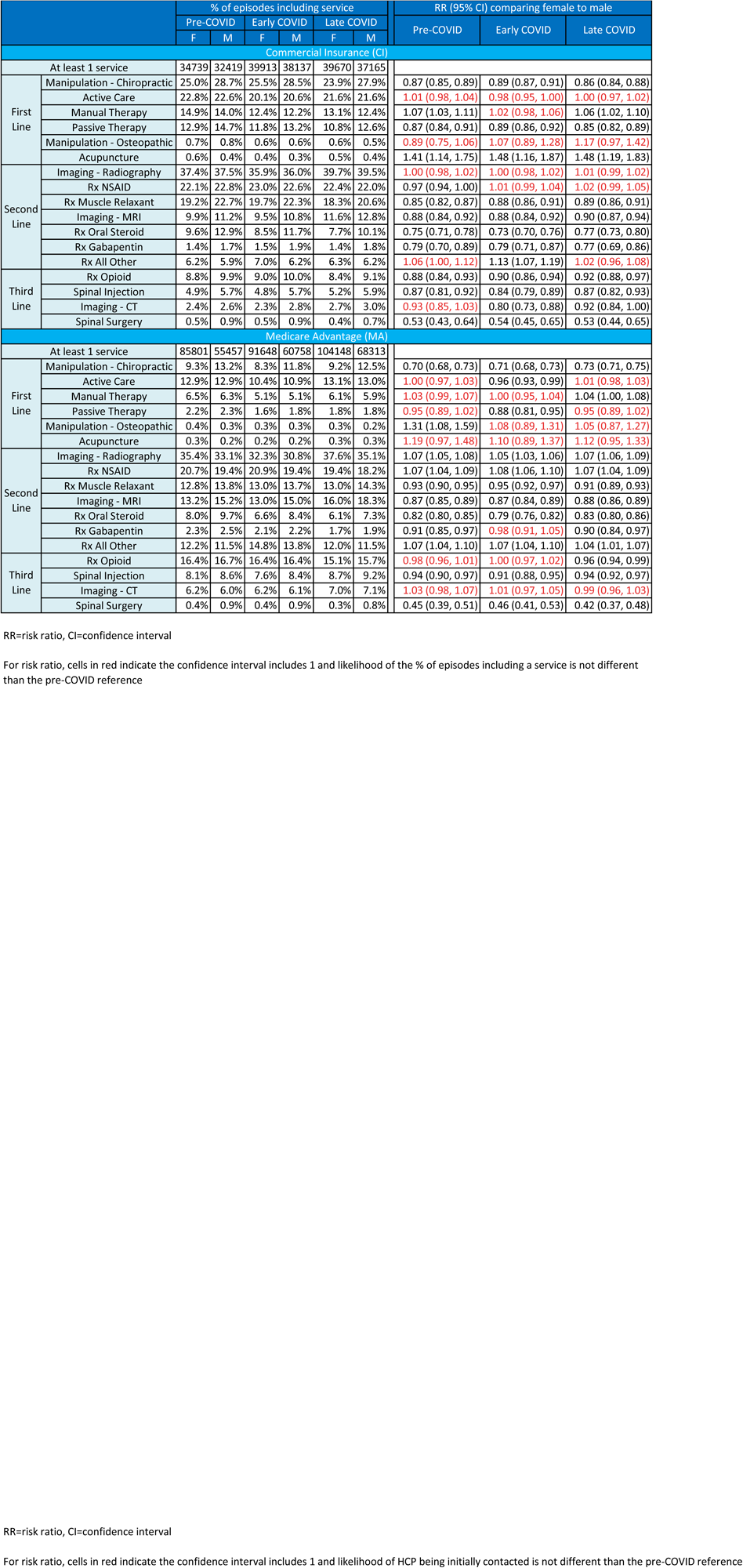
Type of healthcare services provided for low back pain with duration <91 days

The sub analysis of gender variability in the type of HCP initially contacted by individual zip code 5-digit zip code population ADI and percent NHW revealed two types of HCP, DCs and rheumatologists, with significant differences in both CI and MA during all time intervals. [Supplement – Table 2 Initial HCP By ADI] Females were significantly less likely to initially contact a DC in both CI and MA, and during all time periods. In the CI cohort and compared to males, females in zip codes with 0-50% NHW (low % white) population and 51-100 ADI (high deprivation) were least likely to initially contact a DC. This was also observed in the MA cohort during the late COVID period. [Figure 5]

**Figure 5.**
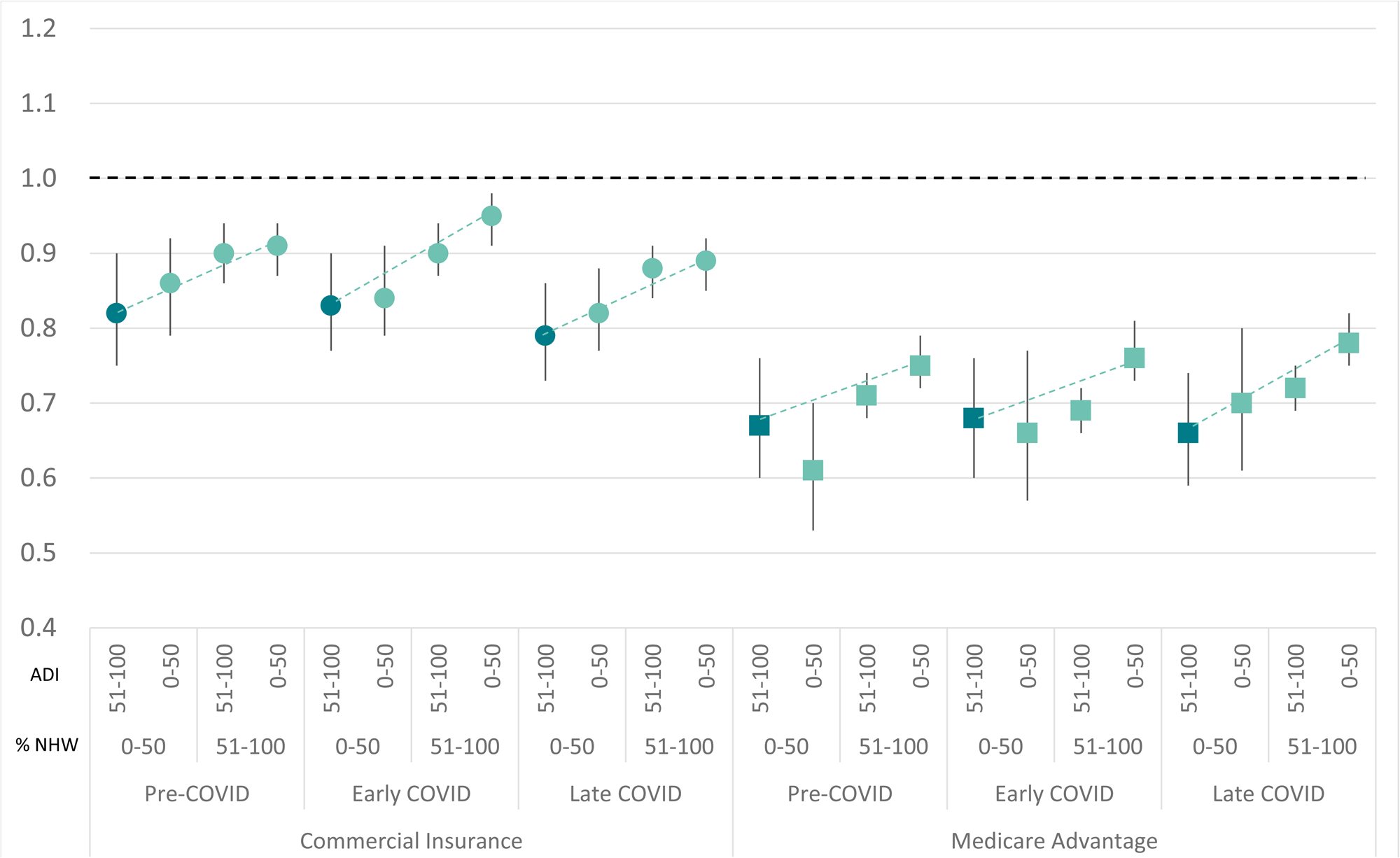
Risk ratio and 95% confidence interval comparing **commercially insured females to males** for the likelihood to select a chiropractor as the initial type of healthcare provider seen for an episode of low back pain with episode lasting <91 days Pre=pre-COVID period, Early=0-12 months post-COVID, Late=13-24 months post-COVID, ADI=Area Deprivation Index of individual’s home address 5-digit zip code, NHW=percent of the individual’s home address 5-digit zip code population that is non-Hispanic white

The sub analysis of gender variability in the type of services received for LBP by individual zip code 5-digit zip code population NHW and ADI revealed females in the 51-100% NHW (high % white) 0-50 ADI (low deprivation) were significantly less like than males to receive chiropractic manipulative treatment, prescription skeletal muscle relaxants, prescription oral steroids, MRI scans, spinal injections, or spinal surgery in both CI and MA during all time intervals. [Supplement – Table 3 Services by ADI] [Figure 6]

**Figure 6.**
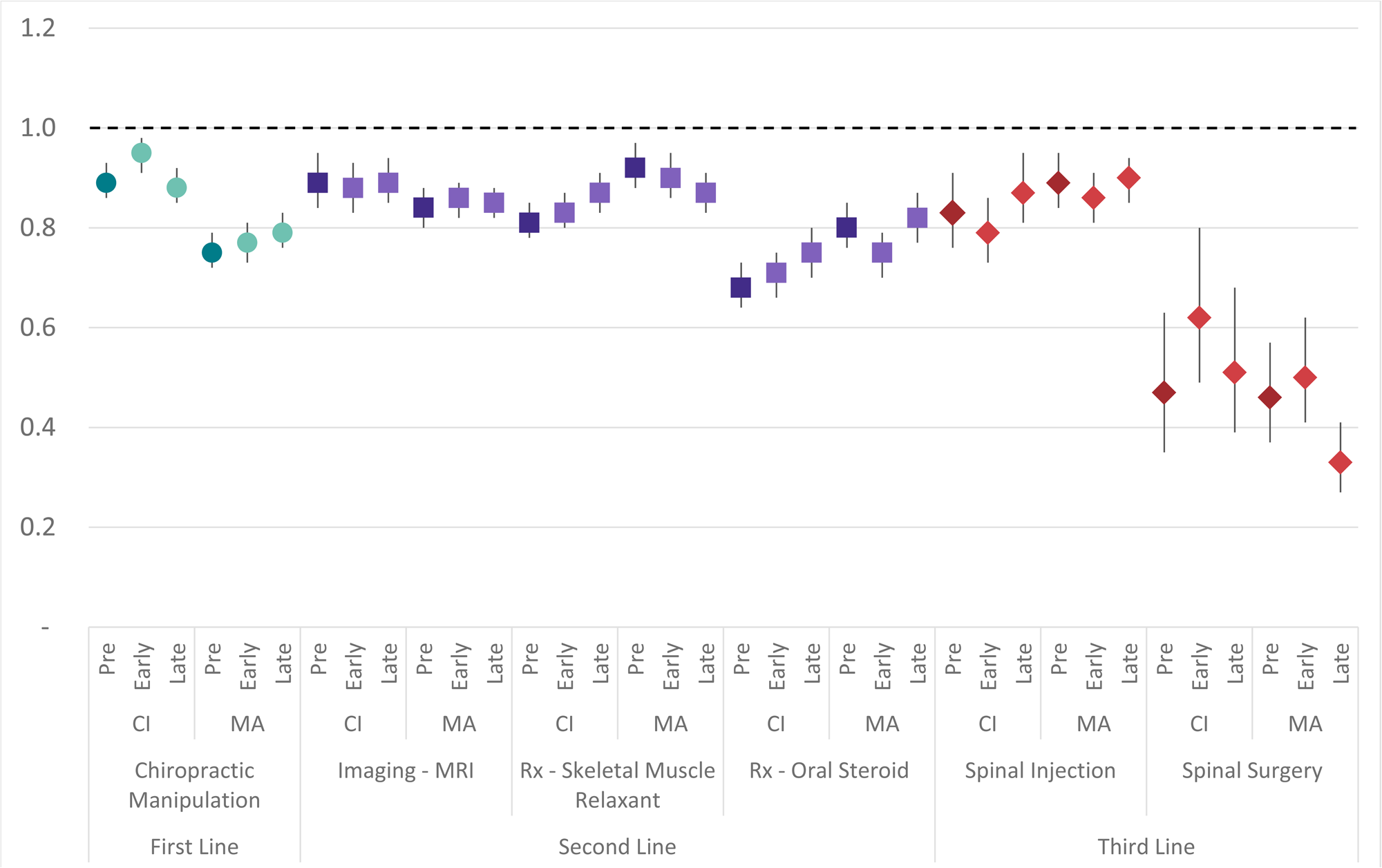
Risk ratio and 95% confidence interval comparing **females to males** living in zip codes with **51-100 Area Deprivation Index (ADI) and 0-50% non-Hispanic white (NHW) population** for the type of healthcare service provided for an episode of low back pain with a duration of <91 days Pre=pre-COVID period, Early=0-12 months post-COVID, Late=13-24 months post-COVID, CI=commercially insured, MA=medicare advantage insured

The sub analysis of gender variability in the type of services received for LBP by the type of HCP initially contacted revealed numerous significant but not clinically meaningful differences. [Supplement – Table 1 – Population By Type of HCP][Supplement – Table 3 – Services By Type of HCP] An example of a clinically meaningful difference was for the three types of HCP most commonly initially contacted, for both the CI and MA cohorts, females with LBP initially contacting a PCP or OS were significantly less likely to receive chiropractic manipulative treatment (CMT). [Figure 7]

**Figure 7.**
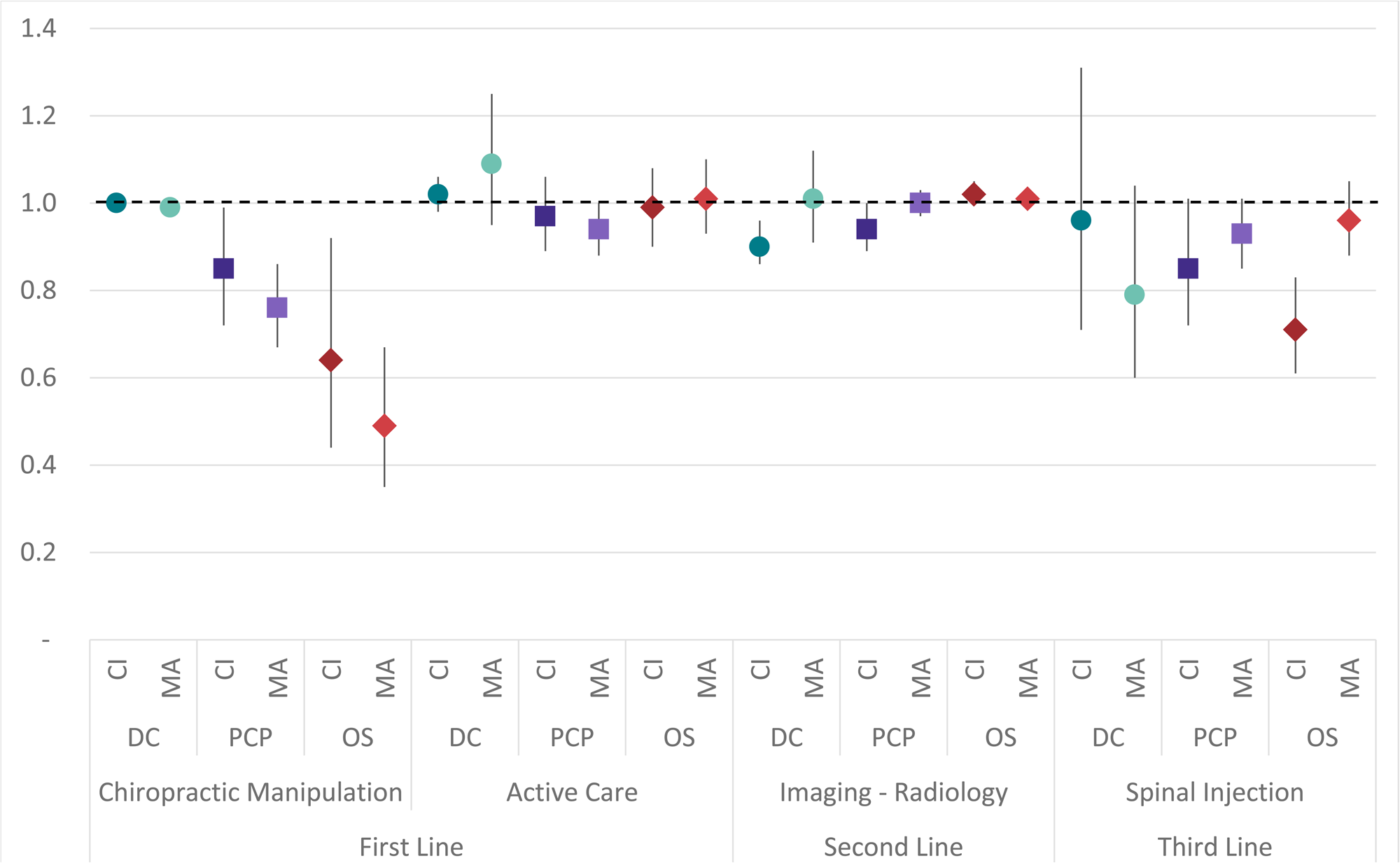
Risk ratio and 95% confidence interval comparing **females to males** for services provided during the late COVID period (March to December 2021) for episodes of low back pain with a duration of <91 days and initial contact with either a Doctor of Chiropractic (DC), Primary Care Physician (PCP), or Orthopedic Surgeon (OS) CI=commercially insured, MA=medicare advantage insured

## Discussion

The degree to which individual gender appeared to influence care seeking for and management of LBP was surprising, with results consistent across pre- and post-COVID periods and in both CI and MA cohorts. With DCs being a common type of HCP initially contacted by individuals with LBP and having total episode attributes aligned with LBP CPGs, the finding that females are less likely than males to initially contact a DC, a finding amplified in low-income, non-white zip codes, is important. Similarly, females with LBP initially contacting a PCP or OS are less likely than males to receive CMT. The degree to which these and other findings are associated with variation in the proportion of female practitioners making up a type of HCP warrants further study to examine HCP gender concordance preference in the management of LBP. This seems particularly plausible for the non-pharmaceutical “hands-on” HCP types like DC, PT and LAc. Additional significant and potentially important findings warranting additional exploration include female patients with LBP being less likely than males to receive spinal surgery, an oral steroid, an MRI scan, or a spinal injection. The degree to which this represents differences in clinical complexity vs. differences in management approaches warrants additional study.

There are numerous limitations and potential confounders to consider. This is particularly true for a study examining gender specific patterns in care seeking for and management of LBP during the pre and post COVID periods in CI and MA cohorts. Variation in the rate of COVID infections by date and geography^57^ and corresponding local COVID public health policies ^42, 43, 58–60^ are two examples. More specifically for LBP, confounders associated with the avoidance of elective spine surgery^61^, reluctance to seek in-person hands on care, reduced capacity of primary care^62^ and emergency departments^63^, and reluctance to use of public transportation ^64, 65^ are additional potential confounders. While a strength of the study is the national scope and inclusion of CI and MA cohorts, this was not a representative sample of the U.S., and the CI and MA cohort distributions were not identical. The potential confounders associated with the heterogenous nature of CI and MA coverage and cohorts and differing proportion of CI and MA episodes from each State made it impossible to directly compare and interpret observed CI and MA differences in the type of HCP initially contacted by individuals with LBP and subsequent management of LBP.

This study corroborated, expanded on, and contrasted with the findings of other studies. Previous research has demonstrated patient-physician gender concordance preference for primary care and emergency medicine HCP types.^13, 14^ The current study of care seeking patterns among individuals with LBP indicates there may be a gender concordance preference among a wider range of HCP types. An earlier study of individuals with LBP found that compared to males, females experienced a greater reduction in pain with acupuncture and less reduction in pain with spinal manipulative therapy.^15^ While the current study did not examine pain reduction of specific services, the study did find that compared to males, females are more likely to seek treatment from LAcs and less likely to seek treatment from DCs. The interaction between gender preference, gender concordance and efficacy of selected treatments warrants additional study. For the pharmaceutical management of LBP an earlier study found a similar rate of prescribing and distribution of pharmaceutical analgesic types for female and male patients. NSAIDs were the most common analgesic prescribed for both females and males, and with males more likely to be prescribed strong opioids. The current study corroborates and expands on this to include additional classes of pharmaceutical prescribed for LBP. An earlier study of individuals undergoing surgical management of lumbar degenerative conditions found that compared to males, females had worse pre- and post-operative pain and similar or greater improvement following the surgical procedure. The current study found that compared to males, in both the CI and MA cohorts and in the pre- and post-COVID periods, females with LBP were significantly less likely to undergo a surgical procedure. The study was not able to determine whether this difference represented females were less likely to have a condition for which a surgical intervention was necessary, or whether in females the pain threshold for performing surgery was higher than in males with a similar condition.

## Conclusions

Care seeking for and management of LBP is different among female and male patients. Females with LBP are less likely than males to seek initial treatment from a DC, and if initially contacting a PCP or OS are less likely to subsequently see a DC. Females are more likely than males to initially contact a Rheumatologist. Females with LBP are less likely than males to receive spinal surgery or injections, MRI, oral steroids, or CMT. The degree to which findings were associated with individual preference for HCP gender concordance could not determined.

## List of Abbreviation

LBP: Low back pain
CI: Commercial Insurance
MA: Medicare Advantage
US: United States
WHO: World Health Organization
CPG: Clinical practice guideline
HCP: Health care provider
ADI: Area Deprivation Index
AGI: Adjusted Gross Income
NHW: Non-Hispanic White
STROBE: Strengthening the Reporting of Observational Studies in Epidemiology
ETG: Episode Treatment Group
ERG: Episode Risk Group
SD: Standard deviation
IQR: Interquartile range
RR: Risk ratio
Q1: 1^st^ Quartile
Q3: 3^rd^ Quartile
PCP: Primary care provider
PA: Physician’s Assistant
DC: Doctor of Chiropractic
PT: Physical Therapist
LAc: Licensed Acupuncturist
OS: Orthopedic Surgeon
PM: Pain Management
EM: Emergency Medicine
CMT: Chiropractic manipulative treatment

## Declarations

### Ethics approval and consent to participate

Because the data was de-identified or a Limited Data Set in compliance with the Health Insurance Portability and Accountability Act and customer requirements, Institutional Review Board approval or waiver of authorization was not required.

### Consent for publication

Not applicable

### Availability of data and materials

The data are proprietary and are not available for public use but, under certain conditions, may be made available to editors and their approved auditors under a data-use agreement to confirm the findings of the current study.

### Authors’ contributions

Study conception and design; **DE**. Data acquisition; **DE, MZ**. Data analysis and interpretation; **DE, MZ**. Draft or revise manuscript; **DE, MZ**.

### Acknowledgements

N/A

## Supporting information

Supplement - Episode Duration

Supplement - Provider Type Gender

Supplement - Public Policy

Supplement - State

Supplement - STROBE Checklist

Supplement - Table 1 - Population By Type of HCP

Supplement - Table 2 - Initial HCP By ADI

Supplement - Table 3 - Services By ADI

Supplement - Table 3 - Services By Type of HCP

## Data Availability

All data produced in the present work are contained in the manuscript

