## Supplement - Episode Duration for "Impact of patient gender on low back pain management before and after the COVID-19 pandemic in commercially insured and Medicare Advantage cohorts. A retrospective cohort study"

| Supplement - Episode Duration | | | | | | |
|---|---|---|---|---|---|---|
| | Pre-COVID | | Early COVID | | Late COVID | |
| | Female | Male | Female | Male | Female | Male |
| **Original Cohort** | | | | | | |
| **Episode Count** | | | | | | |
| Commercially Insured | 81446 | 69315 | 84285 | 73838 | 66497 | 58461 |
| Medicare Advantage | 329705 | 200878 | 303007 | 190566 | 225936 | 142906 |
| **Episode Duration - Median (Q1, Q3)** | | | | | | |
| Commercially Insured | 136 (22, 300) | 111 (15, 286) | 103 (15, 258) | 84 (8, 235) | 58 (6, 156) | 49 (4, 141) |
| Medicare Advantage | 251 (84, 468) | 243 (73, 446) | 209 (62, 316) | 197 (55, 313) | 104 (22, 201) | 97 (20, 196) |
| **Episodes <91 Day Duration** | | | | | | |
| **Episode Count** | | | | | | |
| Commercially Insured | 34739 | 32419 | 39913 | 38137 | 39670 | 37165 |
| Medicare Advantage | 85801 | 55457 | 91648 | 60758 | 104148 | 68313 |
| **Episode Count - % <91 Day Duration** | | | | | | |
| Commercially Insured | 42.7% | 46.8% | 47.4% | 51.6% | 59.7% | 63.6% |
| Medicare Advantage | 26.0% | 27.6% | 30.2% | 31.9% | 46.1% | 47.8% |
| **Episode Duration - Median (Q1, Q3)** | | | | | | |
| Commercially Insured | 13 (1, 43) | 11 (1, 41) | 12 (1, 44) | 10 (1, 42) | 14 (1, 45) | 12 (1, 43) |
| Medicare Advantage | 15 (1, 49) | 15 (1, 48) | 12 (1, 49) | 14 (1, 49) | 16 (1, 51) | 17 (1, 50) |
