## Supplement - Provider Type Gender for "Impact of patient gender on low back pain management before and after the COVID-19 pandemic in commercially insured and Medicare Advantage cohorts. A retrospective cohort study"

| Supplement - Healthcare provider (HCP) gender distribution of types of HCP initially contacted by individuals with LBP | | | |
|---|---|---|---|
| Type of HCP | | % Female | Source |
| Non-Rx | Doctor of Chiropractic (DC) | 32% | 2020 - National Board of Chiropractic Examiners |
| | Physical Therapist (PT) | 65% | 2020 - American Physical Therapy Association |
| | Licensed Acupuncturist (LAc) | 65% | 2023 - Zippia |
| Primary Care | Primary Care Physician (PCP) | 40% | 2020 - American Medical Association |
| | Nurse Practitioner (Nurse) | 87% | 2023 - Zippia |
| | Physician Assistant (PA) | 67% | 2023 - Zippia |
| | Doctor of Osteopathy (DO) | 43% | 2022 - American Osteopathic Association |
| Physician Specialist | Orthopedic Surgeon (OS) | 7% | 2020 - American Medical Association |
| | Physical Medicine and Rehabilitation (PMR) | 36% | 2020 - American Medical Association |
| | Pain Management (PM) | 19% | 2020 - American Medical Association |
| | Neurologist (Neuro) | 31% | 2020 - American Medical Association |
| | Rheumatologist (Rheum) | 46% | 2020 - American Medical Association |
| | Neurosurgeon (NS) | 9% | 2020 - American Medical Association |
| | MD Other (Oth) | - | |
| Emergency Medicine/ Urgent Care | Hospital (Hosp) | - | |
| | Radiologist (Rad) | 26% | 2020 - American Medical Association |
| | Emergency Medicine (EM) | 28% | 2020 - American Medical Association |
| | Urgent Care (UC) | - | |
