## Supplement - Public Policy for "Impact of patient gender on low back pain management before and after the COVID-19 pandemic in commercially insured and Medicare Advantage cohorts. A retrospective cohort study"

### Supplement - State Public Policy Responses to COVID-19

| | Oxford Stringency on April 1 (low = more restrictive)[1] | | | | | | | Raifman (low rank = more restrictive)[2] | | | | | | | | | | | | | | |
|---|---|---|---|---|---|---|---|---|---|---|---|---|---|---|---|---|---|---|---|---|---|---|
| | Score | | Rank | | Category | | | Overall Rank | | Rank | | | | | | | Days Policy In Place | | | | | |
| State | 2020 | 2021 | 2020 | 2021 | 2020 | 2021 | Combination T25=top 25 B25=bottom 25 | Rank Category | Rank | Total | State of Emergency (SOE) | Stay at home (SAH) | Cease elective medical procedures (EMP) | Close non-essential businesses (NEB) | Mask mandate (MM) - SOE | MM - public businesses (PB) | SOE | SAH | EMP | NEB | MM - SOE | MM - PB |
| DC | 87.04 | 61.11 | 3 | 4 | T25 | T25 | *Top 25* | *Most Restrictive* | 10 | 83 | 13 | 16 | 33 | 5 | 8 | 8 | 731 | 58 | 0 | 64 | 611 | 575 |
| VT | 82.41 | 65.74 | 8 | 2 | T25 | T25 | *Top 25* | *Restrictive* | 22 | 140 | 44 | 23 | 9 | 38 | 15 | 11 | 453 | 51 | 44 | 32 | 451 | 417 |
| KY | 85.19 | 58.33 | 5 | 8 | T25 | T25 | *Top 25* | *Most Restrictive* | 6 | 63 | 4 | 4 | 6 | 20 | 12 | 17 | 736 | 93 | 48 | 45 | 455 | 390 |
| RI | 82.41 | 59.26 | 8 | 7 | T25 | T25 | *Top 25* | *Restrictive* | 16 | 119 | 8 | 30 | 33 | 30 | 9 | 9 | 733 | 41 | 0 | 39 | 477 | 438 |
| NY | 82.41 | 58.33 | 8 | 8 | T25 | T25 | *Top 25* | *Most Restrictive* | 2 | 48 | 35 | 3 | 1 | 1 | 6 | 2 | 468 | 95 | 78 | 76 | 692 | 652 |
| CA | 82.41 | 56.94 | 8 | 13 | T25 | T25 | *Top 25* | *Most Restrictive* | 3 | 52 | 2 | 1 | 24 | 16 | 4 | 5 | 738 | 306 | 33 | 49 | 701 | 640 |
| CO | 82.41 | 56.02 | 8 | 14 | T25 | T25 | *Top 25* | *Restrictive* | 27 | 156 | 31 | 35 | 23 | 24 | 22 | 21 | 477 | 31 | 34 | 42 | 424 | 382 |
| OH | 82.41 | 54.63 | 8 | 18 | T25 | T25 | *Top 25* | *Restrictive* | 18 | 130 | 41 | 18 | 11 | 28 | 16 | 16 | 459 | 56 | 43 | 40 | 443 | 393 |
| MN | 82.41 | 54.63 | 8 | 18 | T25 | T25 | *Top 25* | *Restrictive* | 30 | 167 | 35 | 24 | 7 | 42 | 25 | 34 | 468 | 50 | 47 | 29 | 421 | 289 |
| AK | 85.19 | 53.24 | 5 | 21 | T25 | T25 | *Top 25* | *Least Restrictive* | 48 | 246 | 49 | 42 | 26 | 41 | 44 | 44 | 409 | 26 | 31 | 30 | 71 | 28 |
| NM | 87.04 | 52.31 | 3 | 24 | T25 | T25 | *Top 25* | *Most Restrictive* | 4 | 60 | 13 | 2 | 24 | 12 | 5 | 4 | 731 | 246 | 33 | 52 | 696 | 641 |
| WA | 79.63 | 57.41 | 20 | 12 | T25 | T25 | *Top 25* | *Most Restrictive* | 1 | 25 | 1 | 8 | 4 | 4 | 2 | 6 | 742 | 68 | 59 | 66 | 732 | 634 |
| ME | 81.48 | 55.56 | 17 | 16 | T25 | T25 | *Top 25* | *Restrictive* | 28 | 160 | 40 | 14 | 33 | 34 | 20 | 19 | 465 | 59 | 0 | 36 | 429 | 383 |
| KS | 79.63 | 52.78 | 20 | 23 | T25 | T25 | *Top 25* | *Least Restrictive* | 38 | 216 | 44 | 34 | 33 | 36 | 33 | 36 | 453 | 34 | 0 | 34 | 379 | 261 |
| OR | 78.7 | 61.11 | 26 | 4 | B25 | T25 | *Mixed* | *Most Restrictive* | 5 | 62 | 7 | 5 | 16 | 12 | 10 | 12 | 734 | 86 | 38 | 52 | 472 | 411 |
| HI | 76.85 | 71.3 | 29 | 1 | B25 | T25 | *Mixed* | *Most Restrictive* | 8 | 75 | 2 | 12 | 33 | 24 | 1 | 3 | 738 | 66 | 0 | 42 | 742 | 645 |
| IL | 82.41 | 51.85 | 8 | 26 | T25 | B25 | *Mixed* | *Most Restrictive* | 9 | 80 | 8 | 8 | 33 | 3 | 14 | 14 | 733 | 68 | 0 | 68 | 452 | 400 |
| MD | 87.96 | 50.46 | 1 | 33 | T25 | B25 | *Mixed* | *Restrictive* | 16 | 119 | 32 | 27 | 11 | 12 | 19 | 18 | 476 | 45 | 43 | 52 | 430 | 387 |
| ID | 87.96 | 50 | 1 | 34 | T25 | B25 | *Mixed* | *Least Restrictive* | 39 | 212 | 20 | 33 | 33 | 34 | 46 | 46 | 729 | 36 | 0 | 36 | 0 | 0 |
| MA | 75.93 | 62.04 | 33 | 3 | B25 | T25 | *Mixed* | *Restrictive* | 14 | 110 | 42 | 19 | 3 | 10 | 17 | 19 | 455 | 54 | 60 | 54 | 439 | 383 |
| NV | 76.85 | 58.33 | 29 | 8 | B25 | T25 | *Mixed* | *Restrictive* | 15 | 114 | 17 | 32 | 33 | 18 | 7 | 7 | 730 | 39 | 0 | 48 | 688 | 631 |
| LA | 75.93 | 59.72 | 33 | 6 | B25 | T25 | *Mixed* | *Restrictive* | 19 | 133 | 13 | 21 | 16 | 31 | 28 | 24 | 731 | 52 | 38 | 38 | 407 | 357 |
| WV | 82.41 | 50.93 | 8 | 32 | T25 | B25 | *Mixed* | *Restrictive* | 23 | 145 | 29 | 30 | 30 | 28 | 13 | 15 | 726 | 41 | 26 | 40 | 454 | 399 |
| WI | 76.85 | 56.02 | 29 | 14 | B25 | T25 | *Mixed* | *Restrictive* | 29 | 166 | 17 | 25 | 33 | 19 | 33 | 39 | 730 | 48 | 0 | 46 | 379 | 240 |
| NC | 80.56 | 51.85 | 19 | 26 | T25 | B25 | *Mixed* | *Restrictive* | 25 | 149 | 11 | 21 | 33 | 31 | 22 | 31 | 732 | 52 | 0 | 38 | 424 | 318 |
| MI | 75.46 | 58.33 | 37 | 8 | B25 | T25 | *Mixed* | *Restrictive* | 26 | 153 | 51 | 10 | 2 | 7 | 42 | 41 | 202 | 67 | 68 | 62 | 202 | 155 |
| CT | 78.7 | 53.7 | 26 | 20 | B25 | T25 | *Mixed* | *Most Restrictive* | 7 | 73 | 11 | 17 | 33 | 8 | 3 | 1 | 732 | 57 | 0 | 57 | 730 | 707 |
| IN | 79.63 | 51.85 | 20 | 26 | T25 | B25 | *Mixed* | *Restrictive* | 13 | 107 | 4 | 20 | 13 | 11 | 31 | 28 | 736 | 53 | 41 | 53 | 390 | 335 |
| DE | 81.48 | 49.07 | 17 | 35 | T25 | B25 | *Mixed* | *Restrictive* | 20 | 138 | 30 | 10 | 33 | 22 | 21 | 22 | 479 | 67 | 0 | 44 | 428 | 380 |
| FL | 77.31 | 52.31 | 28 | 24 | B25 | T25 | *Mixed* | *Restrictive* | 24 | 146 | 38 | 27 | 9 | 20 | 27 | 25 | 467 | 45 | 44 | 45 | 415 | 353 |
| NH | 85.19 | 41.67 | 5 | 49 | T25 | B25 | *Mixed* | *Restrictive* | 21 | 139 | 20 | 6 | 33 | 23 | 30 | 27 | 729 | 78 | 0 | 43 | 393 | 345 |
| TN | 75.93 | 53.24 | 33 | 21 | B25 | T25 | *Mixed* | *Least Restrictive* | 40 | 215 | 17 | 40 | 19 | 47 | 46 | 46 | 730 | 27 | 37 | 26 | 0 | 0 |
| MT | 79.63 | 48.61 | 20 | 36 | T25 | B25 | *Mixed* | *Least Restrictive* | 42 | 226 | 35 | 37 | 33 | 42 | 39 | 40 | 468 | 28 | 0 | 29 | 330 | 207 |
| WY | 69.44 | 55.56 | 41 | 16 | B25 | T25 | *Mixed* | *Least Restrictive* | 34 | 192 | 20 | 45 | 33 | 24 | 37 | 33 | 729 | 0 | 0 | 42 | 363 | 315 |
| OK | 79.63 | 46.3 | 20 | 40 | T25 | B25 | *Mixed* | *Least Restrictive* | 46 | 239 | 49 | 29 | 20 | 49 | 46 | 46 | 409 | 44 | 36 | 23 | 0 | 0 |
| PA | 79.63 | 43.52 | 20 | 44 | T25 | B25 | *Mixed* | *Most Restrictive* | 12 | 98 | 43 | 13 | 20 | 2 | 10 | 10 | 454 | 64 | 36 | 74 | 472 | 429 |
| VA | 75.93 | 51.85 | 33 | 26 | B25 | B25 | *Bottom 25* | *Restrictive* | 17 | 123 | 34 | 14 | 20 | 5 | 24 | 26 | 469 | 59 | 36 | 64 | 423 | 346 |
| NJ | 76.85 | 47.22 | 29 | 38 | B25 | B25 | *Bottom 25* | *Most Restrictive* | 11 | 94 | 46 | 6 | 4 | 8 | 17 | 13 | 445 | 78 | 59 | 57 | 439 | 410 |
| MO | 67.59 | 51.39 | 43 | 31 | B25 | B25 | *Bottom 25* | *Least Restrictive* | 43 | 222 | 20 | 37 | 33 | 40 | 46 | 46 | 729 | 28 | 0 | 31 | 0 | 0 |
| TX | 75 | 47.69 | 38 | 37 | B25 | B25 | *Bottom 25* | *Least Restrictive* | 32 | 182 | 20 | 36 | 8 | 42 | 38 | 38 | 729 | 29 | 46 | 29 | 357 | 247 |
| GA | 54.63 | 51.85 | 49 | 26 | B25 | B25 | *Bottom 25* | *Least Restrictive* | 37 | 204 | 38 | 37 | 33 | 45 | 28 | 23 | 467 | 28 | 0 | 28 | 407 | 367 |
| SC | 74.07 | 47.22 | 39 | 38 | B25 | B25 | *Bottom 25* | *Least Restrictive* | 44 | 231 | 47 | 40 | 33 | 50 | 26 | 35 | 444 | 27 | 0 | 19 | 418 | 278 |
| AL | 73.61 | 43.52 | 40 | 44 | B25 | B25 | *Bottom 25* | *Least Restrictive* | 33 | 187 | 33 | 42 | 13 | 38 | 32 | 29 | 473 | 26 | 41 | 32 | 386 | 328 |
| UT | 69.44 | 43.98 | 41 | 43 | B25 | B25 | *Bottom 25* | *Least Restrictive* | 37 | 204 | 4 | 45 | 29 | 36 | 45 | 45 | 736 | 0 | 27 | 34 | 55 | 21 |
| AR | 65.74 | 44.44 | 45 | 42 | B25 | B25 | *Bottom 25* | *Least Restrictive* | 45 | 232 | 48 | 45 | 31 | 45 | 33 | 30 | 439 | 0 | 24 | 28 | 379 | 319 |
| AZ | 62.04 | 45.83 | 47 | 41 | B25 | B25 | *Bottom 25* | *Restrictive* | 26 | 153 | 13 | 26 | 15 | 31 | 36 | 32 | 731 | 46 | 40 | 38 | 374 | 317 |
| NE | 66.2 | 43.52 | 44 | 44 | B25 | B25 | *Bottom 25* | *Least Restrictive* | 35 | 195 | 20 | 45 | 26 | 12 | 46 | 46 | 729 | 0 | 31 | 52 | 0 | 0 |
| IA | 62.04 | 43.52 | 47 | 44 | B25 | B25 | *Bottom 25* | *Least Restrictive* | 31 | 180 | 8 | 45 | 28 | 16 | 40 | 43 | 733 | 0 | 30 | 49 | 328 | 80 |
| SD | 65.74 | 41.67 | 45 | 49 | B25 | B25 | *Bottom 25* | *Least Restrictive* | 47 | 240 | 20 | 45 | 32 | 51 | 46 | 46 | 729 | 0 | 22 | 0 | 0 | 0 |
| MS | 50.93 | 43.52 | 51 | 44 | B25 | B25 | *Bottom 25* | *Least Restrictive* | 41 | 221 | 28 | 44 | 16 | 48 | 43 | 42 | 728 | 24 | 38 | 24 | 196 | 143 |
| ND | 51.85 | 39.81 | 50 | 51 | B25 | B25 | *Bottom 25* | *Least Restrictive* | 36 | 203 | 20 | 45 | 33 | 27 | 41 | 37 | 729 | 0 | 0 | 41 | 305 | 260 |

1. Thomas Hale, Noam Angrist, Rafael Goldszmidt, Beatriz Kira, Anna Petherick, Toby Phillips, Samuel Webster, Emily Cameron-Blake, Laura Hallas, Saptarshi Majumdar, and Helen Tatlow. (2021). "A global panel database of pandemic policies (Oxford COVID-19 Government Response Tracker)." Nature Human Behaviour. https://doi.org/10.1038/s41562-021-01079-8 (accessed 3/15/2023)

2. Raifman J, Nocka K, Jones D, Bor J, Lipson S, Jay J, and Chan P. (2020). "COVID-19 US state policy database." Available at: www.tinyurl.com/statepolicies (accessed 3/20/2022)
