## Supplement - State for "Impact of patient gender on low back pain management before and after the COVID-19 pandemic in commercially insured and Medicare Advantage cohorts. A retrospective cohort study"

### Supplement State- Episode distribution by individual home address 5-digit zip code State and insurance coverage

| | Episode Count | | | | | | | | | | | | | | | | % of Episodes | | | | | | | | | | | | | |
|---|---|---|---|---|---|---|---|---|---|---|---|---|---|---|---|---|---|---|---|---|---|---|---|---|---|---|---|---|---|---|
| | Total | | | Medicare Advantage (MA) | | | | | | Commercial Insurance (CI) | | | | | | Total | | | Medicare Advantage (MA) | | | | | | Commercial Insurance (CI) | | | | | |
| | | | | Pre | | Early | | Late | | Pre | | Early | | Late | | | | | Pre | | Early | | Late | | Pre | | Early | | Late | |
| | Total | MA | CI | F | M | F | M | F | M | F | M | F | M | F | M | Total | MA | CI | F | M | F | M | F | M | F | M | F | M | F | M |
| Total | 688168 | 466125 | 222043 | 85801 | 55457 | 91648 | 60758 | 104148 | 68313 | 34739 | 32419 | 39913 | 38137 | 39670 | 37165 | 100.0% | 100.0% | 100.0% | 100.0% | 100.0% | 100.0% | 100.0% | 100.0% | 100.0% | 100.0% | 100.0% | 100.0% | 100.0% | 100.0% | 100.0% |
| FL | 70055 | 50911 | 19144 | 9581 | 6671 | 9767 | 6952 | 10652 | 7288 | 3008 | 2785 | 3503 | 3313 | 3418 | 3117 | 10.2% | 10.9% | 8.6% | 11.2% | 12.0% | 10.7% | 11.4% | 10.2% | 10.7% | 8.7% | 8.6% | 8.8% | 8.7% | 8.6% | 8.4% |
| TX | 66761 | 42595 | 24166 | 7744 | 5208 | 8458 | 5726 | 9300 | 6159 | 3682 | 3524 | 4253 | 4067 | 4466 | 4174 | 9.7% | 9.1% | 10.9% | 9.0% | 9.4% | 9.2% | 9.4% | 8.9% | 9.0% | 10.6% | 10.9% | 10.7% | 10.7% | 11.3% | 11.2% |
| NY | 44878 | 21244 | 23634 | 4054 | 2656 | 4152 | 2775 | 4606 | 3001 | 4112 | 3672 | 4148 | 3628 | 4323 | 3751 | 6.5% | 4.6% | 10.6% | 4.7% | 4.8% | 4.5% | 4.6% | 4.4% | 4.4% | 11.8% | 11.3% | 10.4% | 9.5% | 10.9% | 10.1% |
| CA | 43115 | 29131 | 13984 | 5837 | 3712 | 5852 | 3888 | 5942 | 3900 | 2267 | 2012 | 2532 | 2325 | 2497 | 2351 | 6.3% | 6.2% | 6.3% | 6.8% | 6.7% | 6.4% | 6.4% | 5.7% | 5.7% | 6.5% | 6.2% | 6.3% | 6.1% | 6.3% | 6.3% |
| GA | 34852 | 29167 | 5685 | 6116 | 3377 | 6185 | 3344 | 6546 | 3599 | 851 | 739 | 1104 | 979 | 1046 | 966 | 5.1% | 6.3% | 2.6% | 7.1% | 6.1% | 6.7% | 5.5% | 6.3% | 5.3% | 2.4% | 2.3% | 2.8% | 2.6% | 2.6% | 2.6% |
| IL | 34556 | 23072 | 11484 | 4489 | 2712 | 4396 | 2847 | 5313 | 3315 | 1657 | 1706 | 2116 | 2021 | 2000 | 1984 | 5.0% | 4.9% | 5.2% | 5.2% | 4.9% | 4.8% | 4.7% | 5.1% | 4.9% | 4.8% | 5.3% | 5.3% | 5.3% | 5.0% | 5.3% |
| NC | 27782 | 20065 | 7717 | 3679 | 2029 | 4209 | 2443 | 4802 | 2903 | 1113 | 1179 | 1338 | 1455 | 1315 | 1317 | 4.0% | 4.3% | 3.5% | 4.3% | 3.7% | 4.6% | 4.0% | 4.6% | 4.2% | 3.2% | 3.6% | 3.4% | 3.8% | 3.3% | 3.5% |
| WI | 27616 | 20906 | 6710 | 3859 | 2664 | 4026 | 2903 | 4403 | 3051 | 1037 | 960 | 1198 | 1173 | 1181 | 1161 | 4.0% | 4.5% | 3.0% | 4.5% | 4.8% | 4.4% | 4.8% | 4.2% | 4.5% | 3.0% | 3.0% | 3.0% | 3.1% | 3.0% | 3.1% |
| MO | 25637 | 16688 | 8949 | 2850 | 2032 | 3205 | 2201 | 3749 | 2651 | 1419 | 1305 | 1660 | 1562 | 1606 | 1397 | 3.7% | 3.6% | 4.0% | 3.3% | 3.7% | 3.5% | 3.6% | 3.6% | 3.9% | 4.1% | 4.0% | 4.2% | 4.1% | 4.0% | 3.8% |
| AZ | 23297 | 17290 | 6007 | 3413 | 2248 | 3281 | 2220 | 3659 | 2469 | 1007 | 946 | 1069 | 1041 | 1010 | 934 | 3.4% | 3.7% | 2.7% | 4.0% | 4.1% | 3.6% | 3.7% | 3.5% | 3.6% | 2.9% | 2.9% | 2.7% | 2.7% | 2.5% | 2.5% |
| OH | 20342 | 13208 | 7134 | 2235 | 1548 | 2589 | 1797 | 2961 | 2078 | 1057 | 1069 | 1301 | 1296 | 1217 | 1194 | 3.0% | 2.8% | 3.2% | 2.6% | 2.8% | 2.8% | 3.0% | 2.8% | 3.0% | 3.0% | 3.3% | 3.3% | 3.4% | 3.1% | 3.2% |
| CT | 20080 | 15164 | 4916 | 2799 | 1920 | 2779 | 1933 | 3484 | 2249 | 813 | 697 | 834 | 861 | 887 | 824 | 2.9% | 3.3% | 2.2% | 3.3% | 3.5% | 3.0% | 3.2% | 3.3% | 3.3% | 2.3% | 2.1% | 2.1% | 2.3% | 2.2% | 2.2% |
| NJ | 19488 | 10464 | 9024 | 2034 | 1235 | 1942 | 1316 | 2360 | 1577 | 1550 | 1411 | 1528 | 1411 | 1596 | 1528 | 2.8% | 2.2% | 4.1% | 2.4% | 2.2% | 2.1% | 2.2% | 2.3% | 2.3% | 4.5% | 4.4% | 3.8% | 3.7% | 4.0% | 4.1% |
| IN | 19039 | 14841 | 4198 | 2547 | 1589 | 2930 | 1964 | 3534 | 2277 | 668 | 590 | 753 | 782 | 714 | 691 | 2.8% | 3.2% | 1.9% | 3.0% | 2.9% | 3.2% | 3.2% | 3.4% | 3.3% | 1.9% | 1.8% | 1.9% | 2.1% | 1.8% | 1.9% |
| CO | 17613 | 11346 | 6267 | 2149 | 1381 | 2258 | 1581 | 2319 | 1658 | 1025 | 895 | 1154 | 1055 | 1097 | 1041 | 2.6% | 2.4% | 2.8% | 2.5% | 2.5% | 2.5% | 2.6% | 2.2% | 2.4% | 3.0% | 2.8% | 2.9% | 2.8% | 2.8% | 2.8% |
| SC | 14618 | 12975 | 1643 | 2594 | 1441 | 2833 | 1568 | 2937 | 1602 | 223 | 205 | 302 | 311 | 296 | 306 | 2.1% | 2.8% | 0.7% | 3.0% | 2.6% | 3.1% | 2.6% | 2.8% | 2.3% | 0.6% | 0.6% | 0.8% | 0.8% | 0.7% | 0.8% |
| WA | 13552 | 11145 | 2407 | 2145 | 1448 | 2141 | 1504 | 2280 | 1627 | 326 | 358 | 405 | 461 | 406 | 451 | 2.0% | 2.4% | 1.1% | 2.5% | 2.6% | 2.3% | 2.5% | 2.2% | 2.4% | 0.9% | 1.1% | 1.0% | 1.2% | 1.0% | 1.2% |
| AL | 13091 | 12396 | 695 | 2009 | 1238 | 2716 | 1615 | 3007 | 1811 | 93 | 101 | 110 | 143 | 114 | 134 | 1.9% | 2.7% | 0.3% | 2.3% | 2.2% | 3.0% | 2.7% | 2.9% | 2.7% | 0.3% | 0.3% | 0.3% | 0.4% | 0.3% | 0.4% |
| VA | 13052 | 6436 | 6616 | 1022 | 643 | 1216 | 877 | 1618 | 1060 | 1089 | 963 | 1165 | 1159 | 1158 | 1082 | 1.9% | 1.4% | 3.0% | 1.2% | 1.2% | 1.3% | 1.4% | 1.6% | 1.6% | 3.1% | 3.0% | 2.9% | 3.0% | 2.9% | 2.9% |
| TN | 11984 | 7155 | 4829 | 1237 | 840 | 1456 | 954 | 1574 | 1094 | 702 | 621 | 835 | 873 | 912 | 886 | 1.7% | 1.5% | 2.2% | 1.4% | 1.5% | 1.6% | 1.6% | 1.5% | 1.6% | 2.0% | 1.9% | 2.1% | 2.3% | 2.3% | 2.4% |
| OR | 11178 | 9279 | 1899 | 1698 | 1079 | 1878 | 1203 | 2103 | 1318 | 309 | 253 | 385 | 339 | 323 | 290 | 1.6% | 2.0% | 0.9% | 2.0% | 1.9% | 2.0% | 2.0% | 2.0% | 1.9% | 0.9% | 0.8% | 1.0% | 0.9% | 0.8% | 0.8% |
| UT | 10536 | 8929 | 1607 | 1619 | 1168 | 1699 | 1203 | 1877 | 1363 | 197 | 226 | 324 | 257 | 314 | 289 | 1.5% | 1.9% | 0.7% | 1.9% | 2.1% | 1.9% | 2.0% | 1.8% | 2.0% | 0.6% | 0.7% | 0.8% | 0.7% | 0.8% | 0.8% |
| PA | 8877 | 4914 | 3963 | 825 | 533 | 953 | 633 | 1131 | 839 | 584 | 529 | 669 | 717 | 729 | 735 | 1.3% | 1.1% | 1.8% | 1.0% | 1.0% | 1.0% | 1.0% | 1.1% | 1.2% | 1.7% | 1.6% | 1.7% | 1.9% | 1.8% | 2.0% |
| AR | 8688 | 6105 | 2583 | 1130 | 708 | 1281 | 716 | 1491 | 779 | 364 | 373 | 442 | 461 | 480 | 463 | 1.3% | 1.3% | 1.2% | 1.3% | 1.3% | 1.4% | 1.2% | 1.4% | 1.1% | 1.0% | 1.2% | 1.1% | 1.2% | 1.2% | 1.2% |
| IA | 8655 | 4567 | 4088 | 736 | 516 | 838 | 641 | 1055 | 781 | 608 | 611 | 787 | 765 | 657 | 660 | 1.3% | 1.0% | 1.8% | 0.9% | 0.9% | 0.9% | 1.1% | 1.0% | 1.1% | 1.8% | 1.9% | 2.0% | 2.0% | 1.7% | 1.8% |
| OK | 8457 | 6078 | 2379 | 932 | 598 | 1160 | 782 | 1602 | 1004 | 372 | 343 | 422 | 444 | 425 | 373 | 1.2% | 1.3% | 1.1% | 1.1% | 1.1% | 1.3% | 1.3% | 1.5% | 1.5% | 1.1% | 1.1% | 1.1% | 1.2% | 1.1% | 1.0% |
| MD | 7743 | 779 | 6964 | 145 | 54 | 143 | 75 | 220 | 142 | 1223 | 1018 | 1295 | 1077 | 1257 | 1094 | 1.1% | 0.2% | 3.1% | 0.2% | 0.1% | 0.2% | 0.1% | 0.2% | 0.2% | 3.5% | 3.1% | 3.2% | 2.8% | 3.2% | 2.9% |
| MA | 6072 | 4437 | 1635 | 739 | 474 | 787 | 587 | 1075 | 775 | 198 | 206 | 266 | 284 | 361 | 320 | 0.9% | 1.0% | 0.7% | 0.9% | 0.9% | 0.9% | 1.0% | 1.0% | 1.1% | 0.6% | 0.6% | 0.7% | 0.7% | 0.9% | 0.9% |
| NE | 5261 | 3402 | 1859 | 575 | 380 | 623 | 455 | 799 | 570 | 280 | 288 | 333 | 365 | 312 | 281 | 0.8% | 0.7% | 0.8% | 0.7% | 0.7% | 0.7% | 0.7% | 0.8% | 0.8% | 0.8% | 0.9% | 0.8% | 1.0% | 0.8% | 0.8% |
| LA | 4805 | 1018 | 3787 | 186 | 109 | 180 | 150 | 230 | 163 | 581 | 565 | 715 | 637 | 656 | 633 | 0.7% | 0.2% | 1.7% | 0.2% | 0.2% | 0.2% | 0.2% | 0.2% | 0.2% | 1.7% | 1.7% | 1.8% | 1.7% | 1.7% | 1.7% |
| NM | 4424 | 3752 | 672 | 657 | 444 | 730 | 517 | 853 | 551 | 88 | 96 | 128 | 121 | 139 | 100 | 0.6% | 0.8% | 0.3% | 0.8% | 0.8% | 0.8% | 0.9% | 0.8% | 0.8% | 0.3% | 0.3% | 0.3% | 0.3% | 0.4% | 0.3% |
| ID | 4240 | 3989 | 251 | 693 | 449 | 832 | 497 | 865 | 653 | 28 | 26 | 41 | 43 | 57 | 56 | 0.6% | 0.9% | 0.1% | 0.8% | 0.8% | 0.9% | 0.8% | 0.8% | 1.0% | 0.1% | 0.1% | 0.1% | 0.1% | 0.1% | 0.2% |
| MN | 3426 | 2080 | 1346 | 197 | 168 | 344 | 241 | 655 | 475 | 188 | 190 | 211 | 242 | 262 | 253 | 0.5% | 0.4% | 0.6% | 0.2% | 0.3% | 0.4% | 0.4% | 0.6% | 0.7% | 0.5% | 0.6% | 0.5% | 0.6% | 0.7% | 0.7% |
| KS | 3299 | 2002 | 1297 | 237 | 173 | 316 | 252 | 585 | 439 | 202 | 186 | 291 | 205 | 200 | 213 | 0.5% | 0.4% | 0.6% | 0.3% | 0.3% | 0.3% | 0.4% | 0.6% | 0.6% | 0.6% | 0.6% | 0.7% | 0.5% | 0.5% | 0.6% |
| RI | 3035 | 2326 | 709 | 450 | 304 | 446 | 280 | 511 | 335 | 109 | 106 | 110 | 125 | 132 | 127 | 0.4% | 0.5% | 0.3% | 0.5% | 0.5% | 0.5% | 0.5% | 0.5% | 0.5% | 0.3% | 0.3% | 0.3% | 0.3% | 0.3% | 0.3% |
| NV | 2997 | 1690 | 1307 | 198 | 106 | 355 | 235 | 480 | 316 | 205 | 167 | 243 | 238 | 242 | 212 | 0.4% | 0.4% | 0.6% | 0.2% | 0.2% | 0.4% | 0.4% | 0.5% | 0.5% | 0.6% | 0.5% | 0.6% | 0.6% | 0.6% | 0.6% |
| MI | 2886 | 687 | 2199 | 145 | 79 | 127 | 74 | 156 | 106 | 330 | 315 | 421 | 401 | 393 | 339 | 0.4% | 0.1% | 1.0% | 0.2% | 0.1% | 0.1% | 0.1% | 0.1% | 0.2% | 0.9% | 1.0% | 1.1% | 1.1% | 1.0% | 0.9% |
| ME | 2812 | 2545 | 267 | 434 | 328 | 479 | 337 | 585 | 382 | 45 | 30 | 44 | 57 | 35 | 56 | 0.4% | 0.5% | 0.1% | 0.5% | 0.6% | 0.5% | 0.6% | 0.6% | 0.6% | 0.1% | 0.1% | 0.1% | 0.1% | 0.1% | 0.2% |
| KY | 2752 | 1107 | 1645 | 175 | 138 | 210 | 151 | 257 | 176 | 219 | 226 | 319 | 289 | 294 | 298 | 0.4% | 0.2% | 0.7% | 0.2% | 0.2% | 0.2% | 0.2% | 0.2% | 0.3% | 0.6% | 0.7% | 0.8% | 0.8% | 0.7% | 0.8% |
| NH | 2655 | 2310 | 345 | 371 | 260 | 450 | 290 | 566 | 373 | 43 | 33 | 62 | 61 | 75 | 71 | 0.4% | 0.5% | 0.2% | 0.4% | 0.5% | 0.5% | 0.5% | 0.5% | 0.5% | 0.1% | 0.1% | 0.2% | 0.2% | 0.2% | 0.2% |
| MS | 2258 | 397 | 1861 | 55 | 24 | 59 | 35 | 136 | 88 | 233 | 286 | 333 | 376 | 317 | 316 | 0.3% | 0.1% | 0.8% | 0.1% | 0.0% | 0.1% | 0.1% | 0.1% | 0.1% | 0.7% | 0.9% | 0.8% | 1.0% | 0.8% | 0.9% |
| WV | 1475 | 953 | 522 | 114 | 65 | 186 | 121 | 273 | 194 | 66 | 87 | 91 | 97 | 88 | 93 | 0.2% | 0.2% | 0.2% | 0.1% | 0.1% | 0.2% | 0.2% | 0.3% | 0.3% | 0.2% | 0.3% | 0.2% | 0.3% | 0.2% | 0.3% |
| VT | 1401 | 1352 | 49 | 223 | 163 | 268 | 184 | 291 | 223 | 5 | 6 | 10 | 6 | 11 | 11 | 0.2% | 0.3% | 0.0% | 0.3% | 0.3% | 0.3% | 0.3% | 0.3% | 0.3% | 0.0% | 0.0% | 0.0% | 0.0% | 0.0% | 0.0% |
| HI | 1086 | 1030 | 56 | 176 | 131 | 181 | 155 | 234 | 153 | 7 | 9 | 8 | 15 | 13 | 4 | 0.2% | 0.2% | 0.0% | 0.2% | 0.2% | 0.2% | 0.3% | 0.2% | 0.2% | 0.0% | 0.0% | 0.0% | 0.0% | 0.0% | 0.0% |
| DC | 790 | 38 | 752 | 9 | 5 | 5 | 6 | 9 | 4 | 164 | 101 | 142 | 90 | 141 | 114 | 0.1% | 0.0% | 0.3% | 0.0% | 0.0% | 0.0% | 0.0% | 0.0% | 0.0% | 0.5% | 0.3% | 0.4% | 0.2% | 0.4% | 0.3% |
| DE | 600 | 308 | 292 | 23 | 25 | 34 | 25 | 106 | 95 | 39 | 32 | 64 | 58 | 46 | 53 | 0.1% | 0.1% | 0.1% | 0.0% | 0.0% | 0.0% | 0.0% | 0.1% | 0.1% | 0.1% | 0.1% | 0.2% | 0.2% | 0.1% | 0.1% |
| SD | 451 | 297 | 154 | 18 | 17 | 46 | 45 | 91 | 80 | 22 | 19 | 23 | 28 | 32 | 30 | 0.1% | 0.1% | 0.1% | 0.0% | 0.0% | 0.1% | 0.1% | 0.1% | 0.1% | 0.1% | 0.1% | 0.1% | 0.1% | 0.1% | 0.1% |
| VI | 359 | 135 | 224 | 0 | 1 | 21 | 7 | 72 | 34 | 36 | 34 | 45 | 40 | 35 | 34 | 0.1% | 0.0% | 0.1% | 0.0% | 0.0% | 0.0% | 0.0% | 0.1% | 0.0% | 0.1% | 0.1% | 0.1% | 0.1% | 0.1% | 0.1% |
| WY | 355 | 199 | 156 | 38 | 22 | 32 | 35 | 35 | 37 | 27 | 25 | 21 | 39 | 19 | 25 | 0.1% | 0.0% | 0.1% | 0.0% | 0.0% | 0.0% | 0.1% | 0.0% | 0.1% | 0.1% | 0.1% | 0.1% | 0.1% | 0.0% | 0.1% |
| MT | 302 | 185 | 117 | 32 | 16 | 26 | 26 | 50 | 35 | 10 | 16 | 18 | 25 | 21 | 27 | 0.0% | 0.0% | 0.1% | 0.0% | 0.0% | 0.0% | 0.0% | 0.0% | 0.1% | 0.0% | 0.0% | 0.0% | 0.1% | 0.1% | 0.1% |
| ND | 253 | 116 | 137 | 13 | 4 | 23 | 16 | 37 | 23 | 13 | 14 | 29 | 27 | 28 | 26 | 0.0% | 0.0% | 0.1% | 0.0% | 0.0% | 0.0% | 0.0% | 0.0% | 0.0% | 0.0% | 0.0% | 0.1% | 0.1% | 0.1% | 0.1% |
| AK | 65 | 24 | 41 | 3 | 4 | 6 | 3 | 3 | 5 | 8 | 10 | 6 | 9 | 3 | 5 | 0.0% | 0.0% | 0.0% | 0.0% | 0.0% | 0.0% | 0.0% | 0.0% | 0.0% | 0.0% | 0.0% | 0.0% | 0.0% | 0.0% | 0.0% |
| PR | 62 | 18 | 44 | 5 | 1 | 4 | 3 | 4 | 1 | 2 | 4 | 6 | 14 | 8 | 10 | 0.0% | 0.0% | 0.0% | 0.0% | 0.0% | 0.0% | 0.0% | 0.0% | 0.0% | 0.0% | 0.0% | 0.0% | 0.0% | 0.0% | 0.0% |
| Unknown | 4505 | 2878 | 1627 | 557 | 319 | 535 | 370 | 665 | 432 | 235 | 251 | 301 | 269 | 306 | 265 | 0.7% | 0.6% | 0.7% | 0.6% | 0.6% | 0.6% | 0.6% | 0.6% | 0.6% | 0.7% | 0.8% | 0.8% | 0.7% | 0.8% | 0.7% |
