## Supplement - Table 1 - Population By Type of HCP for "Impact of patient gender on low back pain management before and after the COVID-19 pandemic in commercially insured and Medicare Advantage cohorts. A retrospective cohort study"

| Supplement - Table 1 - Cohort, population and episode attributes for complete low back pain episodes <91 days duration by type of healthcare provider (HCP) initially contacted | | | | | | |
|---|---|---|---|---|---|---|
| | Female | | | Male | | |
| | Pre-COVID | Early COVID | Late COVID | Pre-COVID | Early COVID | Late COVID |
| Commercial | | | | | | |
| Type of HCP initially contacted - Doctor of Chiropractic (DC) | | | | | | |
| Episodes | 8682 | 10118 | 9338 | 9185 | 10761 | 10158 |
| Individuals | 8356 | 9751 | 9214 | 8862 | 10393 | 9970 |
| # Unique HCPs | 5914 | 6620 | 6352 | 6204 | 6885 | 6731 |
| Total Cost | $3,291,636 | $3,504,309 | $3,296,250 | $3,635,743 | $3,973,736 | $3,768,840 |
| Individual Attributes - % of Median (Q1, Q3) | | | | | | |
| % Female | 100% | 100% | 100% | 0% | 0% | 0% |
| Age | 40 (30, 51) | 39 (29, 51) | 40 (30, 52) | 41 (31, 51) | 40 (30, 51) | 41 (31, 52) |
| ERG® Risk Score | 0.9 (0.5, 1.8) | 0.9 (0.5, 1.9) | 1.0 (0.5, 2.1) | 0.6 (0.3, 1.4) | 0.6 (0.3, 1.3) | 0.7 (0.3, 1.5) |
| Individual Home Address 5-digit Zip Code Attributes - Median (Q1, Q3) | | | | | | |
| % Non-Hispanic White (NHW) | 70.4% (53.7%, 82.4%) | 71.4% (53.9%, 83.0%) | 71.0% (53.5%, 83.0%) | 71.0% (53.9%, 82.9%) | 71.4% (54.0%, 83.7%) | 71.0% (53.5%, 83.0%) |
| Area Deprivation Index (ADI) | 42 (24, 60) | 44 (26, 62) | 44 (26, 62) | 44 (25, 62) | 46 (27, 63) | 44 (25, 63) |
| Household Adjusted Gross Income (AGI) - 1000s | 76 (58, 109) | 74 (57, 104) | 74 (57, 104) | 75 (57, 107) | 72 (56, 102) | 74 (57, 105) |
| Total Episode Attributes - Median (Q1, Q3) | | | | | | |
| Cost | $180 (84, 385) | $176 (80, 371) | $180 (80, 385) | $167 (74, 363) | $161 (72, 353) | $165 (71, 360) |
| # Healthcare Providers Seen | 1 (1, 1) | 1 (1, 1) | 1 (1, 1) | 1 (1, 1) | 1 (1, 1) | 1 (1, 1) |
| Duration (days) | 17 (2, 44) | 17 (3, 47) | 18 (3, 45) | 14 (1, 40) | 13 (1, 41) | 14 (1, 42) |
| Type of HCP initially contacted - Primary Care Physician (PCP) | | | | | | |
| Episodes | 7071 | 8992 | 7860 | 6626 | 8207 | 7304 |
| Individuals | 6964 | 8871 | 7851 | 6524 | 8103 | 7290 |
| # Unique HCPs | 6208 | 7915 | 6977 | 5826 | 7120 | 6383 |
| Total Cost | $3,147,397 | $4,027,695 | $3,575,960 | $3,731,185 | $4,669,495 | $4,337,049 |
| Individual Attributes - % of Median (Q1, Q3) | | | | | | |
| % Female | 100% | 100% | 100% | 0% | 0% | 0% |
| Age | 48 (37, 56) | 48 (37, 57) | 49 (37, 58) | 49 (38, 57) | 49 (39, 58) | 50 (40, 58) |
| ERG® Risk Score | 1.3 (0.7, 2.7) | 1.4 (0.7, 2.9) | 1.6 (0.8, 3.0) | 1.0 (0.5, 2.0) | 1.1 (0.5, 2.3) | 1.2 (0.6, 2.4) |
| Individual Home Address 5-digit Zip Code Attributes - Median (Q1, Q3) | | | | | | |
| % Non-Hispanic White (NHW) | 60.3% (39.1%, 76.6%) | 61.2% (38.2%, 76.6%) | 60.4% (38.5%, 76.2%) | 62.4% (41.9%, 77.3%) | 63.1% (43.1%, 78.4%) | 62.7% (41.8%, 78.2%) |
| Area Deprivation Index (ADI) | 39 (21, 59) | 40 (22, 60) | 41 (21, 60) | 40 (21, 59) | 41 (22, 60) | 40 (21, 61) |
| Household Adjusted Gross Income (AGI) - 1000s | 70 (53, 102) | 69 (52, 98) | 70 (52, 99) | 71 (54, 102) | 70 (53, 102) | 70 (53, 102) |
| Total Episode Attributes - Median (Q1, Q3) | | | | | | |
| Cost | $118 (19, 318) | $85 (15, 265) | $98 (16, 311) | $141 (30, 363) | $117 (23, 310) | $122 (20, 362) |
| # Healthcare Providers Seen | 1 (1, 2) | 1 (1, 2) | 1 (1, 2) | 1 (1, 2) | 1 (1, 2) | 1 (1, 2) |
| Duration (days) | 6 (1, 43) | 4 (1, 40) | 4 (1, 43) | 7 (1, 39) | 4 (1, 39) | 6 (1, 42) |
| Type of HCP initially contacted - Orthopedic Surgeon (OS) | | | | | | |
| Episodes | 2987 | 3346 | 3509 | 2744 | 3302 | 3299 |
| Individuals | 2953 | 3316 | 3508 | 2722 | 3283 | 3297 |
| # Unique HCPs | 2033 | 2266 | 2342 | 1913 | 2251 | 2270 |
| Total Cost | $3,378,164 | $2,842,037 | $2,997,549 | $3,037,761 | $3,525,228 | $3,646,879 |
| Individual Attributes - % of Median (Q1, Q3) | | | | | | |
| % Female | 100% | 100% | 100% | 0% | 0% | 0% |
| Age | 46 (27, 56) | 47 (28, 57) | 48 (31, 58) | 47 (29, 57) | 46 (30, 57) | 48 (32, 58) |
| ERG® Risk Score | 1.4 (0.7, 2.7) | 1.5 (0.7, 2.9) | 1.6 (0.8, 3.1) | 1.2 (0.5, 2.5) | 1.2 (0.5, 2.6) | 1.4 (0.6, 2.8) |
| Individual Home Address 5-digit Zip Code Attributes - Median (Q1, Q3) | | | | | | |
| % Non-Hispanic White (NHW) | 64.7% (48.2%, 77.2%) | 64.1% (45.8%, 76.6%) | 65.0% (47.5%, 76.2%) | 65.1% (49.4%, 77.3%) | 64.6% (47.9%, 76.9%) | 65.1% (48.8%, 77.3%) |
| Area Deprivation Index (ADI) | 26 (12, 46) | 27 (14, 47) | 27 (14, 50) | 26 (13, 46) | 27 (14, 48) | 27 (13, 48) |
| Household Adjusted Gross Income (AGI) - 1000s | 94 (65, 145) | 89 (64, 141) | 89 (62, 139) | 95 (66, 149) | 92 (64, 141) | 92 (64, 143) |
| Total Episode Attributes - Median (Q1, Q3) | | | | | | |
| Cost | $289 (151, 750) | $298 (148, 760) | $321 (160, 757) | $297 (157, 783) | $327 (160, 828) | $342 (165, 936) |
| # Healthcare Providers Seen | 1 (1, 2) | 1 (1, 2) | 1 (1, 2) | 1 (1, 2) | 1 (1, 2) | 1 (1, 2) |
| Duration (days) | 8 (1, 40) | 10 (1, 43) | 12 (1, 43) | 11 (1, 38) | 11 (1, 40) | 14 (1, 44) |
| Medicare Advantage | | | | | | |
| Type of HCP initially contacted - Doctor of Chiropractic (DC) | | | | | | |
| Episodes | 7059 | 6767 | 8292 | 6506 | 6393 | 7480 |
| Individuals | 6802 | 6569 | 8187 | 6309 | 6189 | 7394 |
| # Unique HCPs | 4754 | 4638 | 5716 | 4476 | 4392 | 5168 |
| Total Cost | $1,574,291 | $1,647,355 | $2,093,569 | $1,594,978 | $1,543,041 | $1,888,137 |
| Individual Attributes - % of Median (Q1, Q3) | | | | | | |
| % Female | 100% | 100% | 100% | 0% | 0% | 0% |
| Age | 72 (68, 76) | 72 (68, 77) | 72 (68, 77) | 72 (68, 77) | 72 (68, 77) | 72 (68, 77) |
| ERG® Risk Score | 0.7 (0.3, 1.2) | 0.7 (0.4, 1.2) | 0.8 (0.4, 1.3) | 0.7 (0.4, 1.3) | 0.8 (0.4, 1.3) | 0.8 (0.4, 1.4) |
| Individual Home Address 5-digit Zip Code Attributes - Median (Q1, Q3) | | | | | | |
| % Non-Hispanic White (NHW) | 77.6% (63.1%, 87.1%) | 78.5% (64.4%, 87.7%) | 78.1% (64.0%, 87.3%) | 78.6% (63.8%, 87.8%) | 79.7% (65.6%, 88.6%) | 79.3% (65.4%, 88.1%) |
| Area Deprivation Index (ADI) | 53 (36, 69) | 54 (38, 70) | 54 (38, 70) | 53 (36, 68) | 54 (38, 70) | 54 (38, 70) |
| Household Adjusted Gross Income (AGI) - 1000s | 65 (53, 86) | 64 (53, 83) | 64 (53, 84) | 66 (53, 86) | 64 (53, 84) | 64 (53, 84) |
| Total Episode Attributes - Median (Q1, Q3) | | | | | | |
| Cost | $123 (58, 248) | $131 (58, 267) | $135 (61, 277) | $117 (56, 245) | $122 (57, 255) | $128 (59, 268) |
| # Healthcare Providers Seen | 1 (1, 1) | 1 (1, 1) | 1 (1, 1) | 1 (1, 1) | 1 (1, 1) | 1 (1, 1) |
| Duration (days) | 22 (5, 50) | 22 (5, 50) | 22 (6, 51) | 19 (4, 46) | 19 (4, 48) | 20 (5, 48) |
| Type of HCP initially contacted - Primary Care Physician (PCP) | | | | | | |
| Episodes | 27246 | 32374 | 30894 | 17255 | 20906 | 19987 |
| Individuals | 26677 | 31748 | 30844 | 16890 | 20525 | 19960 |
| # Unique HCPs | 19077 | 22114 | 21960 | 13083 | 15565 | 15416 |
| Total Cost | $5,621,082 | $6,176,616 | $6,685,227 | $3,793,046 | $4,501,602 | $5,054,857 |
| Individual Attributes - % of Median (Q1, Q3) | | | | | | |
| % Female | 100% | 100% | 100% | 0% | 0% | 0% |
| Age | 72 (68, 79) | 73 (68, 79) | 73 (68, 79) | 72 (67, 77) | 72 (67, 78) | 72 (67, 78) |
| ERG® Risk Score | 0.9 (0.5, 1.6) | 1.0 (0.5, 1.7) | 1.1 (0.5, 1.8) | 0.9 (0.4, 1.6) | 1.0 (0.4, 1.7) | 1.0 (0.5, 1.8) |
| Individual Home Address 5-digit Zip Code Attributes - Median (Q1, Q3) | | | | | | |
| % Non-Hispanic White (NHW) | 62.7% (39.4%, 79.1%) | 63.1% (39.8%, 79.2%) | 63.0% (40.0%, 79.0%) | 65.0% (42.6%, 80.5%) | 65.6% (43.2%, 80.7%) | 65.9% (43.5%, 80.6%) |
| Area Deprivation Index (ADI) | 53 (32, 71) | 53 (33, 71) | 53 (33, 71) | 52 (32, 70) | 52 (32, 70) | 52 (32, 70) |
| Household Adjusted Gross Income (AGI) - 1000s | 60 (48, 80) | 60 (48, 79) | 60 (48, 79) | 61 (48, 83) | 61 (49, 82) | 61 (49, 83) |
| Total Episode Attributes - Median (Q1, Q3) | | | | | | |
| Cost | $43 (15, 149) | $41 (15, 126) | $46 (16, 163) | $45 (16, 156) | $44 (16, 132) | $46 (16, 171) |
| # Healthcare Providers Seen | 1 (1, 2) | 1 (1, 1) | 1 (1, 2) | 1 (1, 2) | 1 (1, 2) | 1 (1, 2) |
| Duration (days) | 10 (1, 51) | 5 (1, 50) | 12 (1, 53) | 9 (1, 49) | 6 (1, 50) | 12 (1, 53) |
| Type of HCP initially contacted - Orthopedic Surgeon (OS) | | | | | | |
| Episodes | 5546 | 5816 | 7022 | 3569 | 3918 | 4673 |
| Individuals | 5502 | 5777 | 7017 | 3549 | 3900 | 4672 |
| # Unique HCPs | 3417 | 3607 | 4073 | 2457 | 2703 | 3123 |
| Total Cost | $2,699,439 | $3,296,508 | $3,972,274 | $2,276,110 | $2,433,154 | $2,920,096 |
| Individual Attributes - % of Median (Q1, Q3) | | | | | | |
| % Female | 100% | 100% | 100% | 0% | 0% | 0% |
| Age | 72 (68, 78) | 73 (68, 78) | 73 (68, 78) | 73 (68, 78) | 73 (68, 78) | 73 (69, 78) |
| ERG® Risk Score | 1.0 (0.6, 1.7) | 1.1 (0.6, 1.7) | 1.1 (0.6, 1.8) | 1.1 (0.6, 1.7) | 1.1 (0.6, 1.8) | 1.1 (0.6, 1.9) |
| Individual Home Address 5-digit Zip Code Attributes - Median (Q1, Q3) | | | | | | |
| % Non-Hispanic White (NHW) | 65.7% (45.4%, 79.4%) | 65.9% (46.1%, 79.5%) | 66.5% (47.2%, 79.8%) | 68.2% (49.6%, 80.8%) | 68.9% (50.0%, 80.7%) | 69.0% (51.3%, 81.0%) |
| Area Deprivation Index (ADI) | 51 (32, 68) | 53 (34, 69) | 52 (34, 69) | 49 (31, 66) | 51 (32, 68) | 50 (31, 67) |
| Household Adjusted Gross Income (AGI) - 1000s | 64 (50, 89) | 62 (49, 87) | 63 (50, 88) | 67 (52, 93) | 65 (51, 91) | 67 (52, 93) |
| Total Episode Attributes - Median (Q1, Q3) | | | | | | |
| Cost | $200 (78, 515) | $197 (55, 521) | $237 (92, 609) | $224 (98, 582) | $229 (95, 585) | $252 (110, 637) |
| # Healthcare Providers Seen | 1 (1, 2) | 1 (1, 2) | 1 (1, 2) | 1 (1, 2) | 1 (1, 2) | 1 (1, 2) |
| Duration (days) | 18 (1, 49) | 16 (1, 50) | 21 (1, 51) | 19 (1, 46) | 21 (1, 49) | 21 (1, 50) |

Within CI and MA, compared to males for the same time period, cells in red indicate females are not significantly different - Mann-Whitney U test (p=0.05)
