## Supplement - Table 2 - Initial HCP By ADI for "Impact of patient gender on low back pain management before and after the COVID-19 pandemic in commercially insured and Medicare Advantage cohorts. A retrospective cohort study"

**Supplement - Table 2 - Type of healthcare provider initially contacted by patient home address 5 digit zip code population measures of race/ethnicity and socioeconomic status**

| | | % of Episodes Where Type of HCP Was Initially Contacted | | | | | | | | | | | | Risk Ratio and 95% Confidence Interval Comparing Female to Male Baseline | | | | | |
|---|---|---|---|---|---|---|---|---|---|---|---|---|---|---|---|---|---|---|---|
| | | Commercial Insurance (CI) | | | | | | Medicare Advantage (MA) | | | | | | Commercial Insurance (CI) | | | Medicare Advantage (MA) | | |
| | | Pre-COVID | | Early COVID | | Late COVID | | Pre-COVID | | Early COVID | | Late COVID | | Pre-COVID | Early COVID | Late COVID | Pre-COVID | Early COVID | Late COVID |
| | | F | M | F | M | F | M | F | M | F | M | F | M | | | | | | |
| % of Population that is non-Hispanic white (NHW 0%-50%) and Area Deprivation Index (ADI 51-100) - **Non-White, Low Income** | | | | | | | | | | | | | | | | | | | |
| Episodes | | 3453 | 3208 | 4313 | 3972 | 4239 | 3739 | 14592 | 8275 | 15752 | 8998 | 17272 | 9870 | | | | | | |
| Non-Rx | DC | 19.8% | 24.1% | 19.7% | 23.6% | 18.8% | 23.8% | 3.7% | 5.5% | 3.4% | 5.1% | 3.8% | 5.7% | **0.82 (0.75, 0.90)** | **0.83 (0.77, 0.90)** | **0.79 (0.73, 0.86)** | **0.67 (0.60, 0.76)** | **0.68 (0.60, 0.76)** | **0.66 (0.59, 0.74)** |
| | PT | 2.5% | 1.3% | 1.7% | 1.5% | 2.1% | 1.9% | 2.4% | 2.4% | 1.9% | 1.8% | 2.3% | 2.1% | 1.88 (1.30, 2.71) | 1.19 (0.85, 1.67) | 1.11 (0.81, 1.51) | 0.98 (0.83, 1.17) | 1.07 (0.88, 1.30) | 1.08 (0.92, 1.28) |
| | LAc | 0.1% | 0.1% | 0.1% | 0.1% | 0.2% | 0.1% | 0.1% | 0.0% | 0.0% | 0.0% | 0.1% | 0.1% | 0.93 (0.23, 3.71) | 2.30 (0.45, 11.86) | 2.94 (0.81, 10.68) | 9.07 (1.20, 68.41) | 0.86 (0.24, 3.04) | 1.83 (0.67, 4.99) |
| Primary Care | PCP | 25.0% | 24.1% | 27.8% | 24.8% | 25.3% | 24.6% | 35.9% | 35.5% | 39.9% | 39.1% | 35.2% | 34.0% | 1.04 (0.95, 1.13) | 1.12 (1.04, 1.20) | 1.03 (0.96, 1.11) | 1.01 (0.98, 1.05) | 1.02 (0.99, 1.05) | 1.04 (1.00, 1.07) |
| | Nurse | 7.7% | 7.5% | 8.9% | 7.4% | 7.9% | 7.2% | 7.5% | 7.1% | 8.1% | 6.8% | 7.7% | 7.3% | 1.02 (0.86, 1.21) | 1.21 (1.05, 1.40) | 1.10 (0.94, 1.28) | 1.05 (0.96, 1.16) | 1.19 (1.08, 1.30) | 1.05 (0.96, 1.15) |
| | PA | 6.0% | 6.7% | 5.1% | 6.1% | 5.1% | 5.2% | 5.0% | 5.2% | 4.6% | 4.9% | 5.1% | 4.9% | 0.89 (0.74, 1.07) | 0.83 (0.70, 1.00) | 0.98 (0.81, 1.18) | 0.96 (0.85, 1.07) | 0.94 (0.83, 1.05) | 1.03 (0.92, 1.15) |
| | DO | 0.2% | 0.2% | 0.3% | 0.3% | 0.2% | 0.3% | 0.2% | 0.1% | 0.1% | 0.1% | 0.1% | 0.1% | 1.08 (0.36, 3.22) | 1.01 (0.43, 2.38) | 0.47 (0.19, 1.19) | 1.42 (0.68, 2.95) | 1.03 (0.47, 2.23) | 1.01 (0.51, 1.99) |
| Physician Specialist | OS | 6.7% | 6.0% | 6.3% | 7.0% | 7.0% | 6.4% | 6.4% | 6.1% | 6.4% | 6.3% | 6.8% | 6.3% | 1.13 (0.94, 1.36) | 0.91 (0.77, 1.07) | 1.09 (0.93, 1.29) | 1.05 (0.94, 1.16) | 1.01 (0.92, 1.12) | 1.06 (0.97, 1.17) |
| | PMR | 2.3% | 1.4% | 1.9% | 1.5% | 1.7% | 2.0% | 1.9% | 1.8% | 1.5% | 1.6% | 1.7% | 1.4% | 1.69 (1.17, 2.43) | 1.26 (0.91, 1.76) | 0.85 (0.61, 1.17) | 1.03 (0.85, 1.25) | 0.97 (0.79, 1.20) | 1.18 (0.97, 1.44) |
| | PM | 1.9% | 1.1% | 1.5% | 1.5% | 1.6% | 1.4% | 3.0% | 3.2% | 3.1% | 3.0% | 3.0% | 3.0% | 1.75 (1.16, 2.64) | 1.00 (0.71, 1.41) | 1.18 (0.82, 1.69) | 0.93 (0.80, 1.08) | 1.01 (0.88, 1.17) | 1.01 (0.88, 1.16) |
| | Neuro | 0.5% | 0.1% | 0.4% | 0.3% | 0.3% | 0.3% | 0.5% | 0.4% | 0.6% | 0.5% | 0.5% | 0.4% | 4.18 (1.42, 12.34) | 1.47 (0.67, 3.24) | 1.12 (0.51, 2.47) | 1.26 (0.85, 1.88) | 1.18 (0.82, 1.71) | 1.10 (0.76, 1.59) |
| | Rheum | 0.5% | 0.2% | 0.8% | 0.3% | 0.5% | 0.1% | 1.1% | 0.5% | 1.1% | 0.4% | 0.9% | 0.3% | 2.79 (1.11, 7.01) | 2.69 (1.40, 5.17) | 5.07 (1.76, 14.65) | 2.35 (1.67, 3.32) | 2.73 (1.90, 3.92) | 2.93 (2.01, 4.27) |
| | NS | 0.2% | 0.4% | 0.3% | 0.4% | 0.3% | 0.4% | 0.3% | 0.5% | 0.3% | 0.3% | 0.2% | 0.3% | 0.62 (0.25, 1.51) | 0.70 (0.34, 1.45) | 0.71 (0.33, 1.51) | 0.52 (0.34, 0.82) | 0.94 (0.57, 1.54) | 0.88 (0.55, 1.41) |
| | MD Oth | 1.4% | 1.1% | 1.5% | 1.4% | 1.4% | 1.1% | 1.7% | 2.2% | 2.3% | 2.6% | 1.8% | 2.6% | 1.30 (0.85, 2.00) | 1.13 (0.79, 1.61) | 1.27 (0.85, 1.89) | 0.76 (0.63, 0.92) | 0.90 (0.77, 1.06) | 0.71 (0.60, 0.84) |
| Emergency Medicine/ Urgent Care | Hosp | 11.4% | 12.3% | 10.5% | 11.7% | 12.8% | 12.0% | 16.2% | 15.2% | 13.9% | 13.7% | 16.3% | 16.2% | 0.92 (0.81, 1.05) | 0.89 (0.79, 1.01) | 1.06 (0.95, 1.19) | 1.06 (1.00, 1.13) | 1.01 (0.95, 1.08) | 1.00 (0.95, 1.06) |
| | Rad | 9.2% | 8.4% | 9.3% | 7.1% | 10.3% | 8.3% | 11.4% | 11.3% | 10.6% | 11.0% | 12.1% | 12.4% | 1.09 (0.93, 1.27) | 1.31 (1.13, 1.52) | 1.24 (1.08, 1.43) | 1.01 (0.93, 1.09) | 0.97 (0.90, 1.04) | 0.97 (0.91, 1.04) |
| | EM | 3.6% | 3.9% | 2.9% | 4.0% | 3.5% | 3.9% | 2.4% | 2.6% | 1.9% | 2.5% | 2.1% | 2.5% | 0.91 (0.72, 1.17) | 0.74 (0.59, 0.93) | 0.89 (0.71, 1.11) | 0.93 (0.79, 1.10) | 0.77 (0.65, 0.92) | 0.82 (0.70, 0.96) |
| | UC | 1.1% | 1.1% | 1.0% | 1.2% | 0.9% | 1.0% | 0.4% | 0.3% | 0.3% | 0.4% | 0.4% | 0.3% | 0.95 (0.61, 1.51) | 0.81 (0.54, 1.21) | 0.90 (0.58, 1.40) | 1.47 (0.90, 2.40) | 0.84 (0.55, 1.28) | 1.51 (0.98, 2.33) |
| NHW 0-50 and ADI 0-50 - **Non-White, High Income** | | | | | | | | | | | | | | | | | | | |
| Episodes | | 6344 | 5480 | 6970 | 6131 | 7189 | 6362 | 10708 | 6432 | 10606 | 6706 | 11677 | 7371 | | | | | | |
| Non-Rx | DC | 17.6% | 20.6% | 17.9% | 21.3% | 16.8% | 20.5% | 3.8% | 6.2% | 3.1% | 4.7% | 3.6% | 5.1% | **0.86 (0.79, 0.92)** | **0.84 (0.79, 0.91)** | **0.82 (0.77, 0.88)** | **0.61 (0.53, 0.70)** | **0.66 (0.57, 0.77)** | **0.70 (0.61, 0.80)** |
| | PT | 5.0% | 3.9% | 3.7% | 3.3% | 4.1% | 3.0% | 4.0% | 3.9% | 2.6% | 2.6% | 3.7% | 3.4% | 1.29 (1.09, 1.53) | 1.13 (0.94, 1.35) | 1.39 (1.16, 1.66) | 1.02 (0.87, 1.18) | 1.00 (0.83, 1.20) | 1.08 (0.93, 1.26) |
| | LAc | 0.4% | 0.2% | 0.4% | 0.2% | 0.5% | 0.2% | 0.2% | 0.3% | 0.2% | 0.1% | 0.3% | 0.3% | 1.96 (0.97, 3.99) | 1.83 (0.94, 3.54) | 3.14 (1.61, 6.12) | 0.74 (0.42, 1.32) | 1.26 (0.59, 2.70) | 1.01 (0.61, 1.66) |
| Primary Care | PCP | 26.3% | 25.6% | 29.1% | 26.4% | 25.2% | 23.6% | 39.8% | 39.4% | 45.4% | 45.6% | 38.7% | 38.6% | 1.03 (0.97, 1.09) | 1.10 (1.04, 1.17) | 1.07 (1.00, 1.13) | 1.01 (0.97, 1.05) | 1.00 (0.96, 1.03) | 1.00 (0.97, 1.04) |
| | Nurse | 3.8% | 3.4% | 3.2% | 3.2% | 3.4% | 3.4% | 3.8% | 3.6% | 4.1% | 3.6% | 3.3% | 3.3% | 1.11 (0.92, 1.34) | 1.00 (0.83, 1.21) | 0.99 (0.83, 1.19) | 1.05 (0.90, 1.23) | 1.14 (0.98, 1.33) | 1.00 (0.86, 1.18) |
| | PA | 4.2% | 4.9% | 3.7% | 3.9% | 3.4% | 3.9% | 3.4% | 3.8% | 3.5% | 3.3% | 3.5% | 3.6% | 0.86 (0.73, 1.02) | 0.96 (0.81, 1.15) | 0.86 (0.72, 1.02) | 0.89 (0.76, 1.05) | 1.06 (0.90, 1.25) | 0.97 (0.83, 1.12) |
| | DO | 0.2% | 0.2% | 0.2% | 0.3% | 0.2% | 0.2% | 0.2% | 0.1% | 0.1% | 0.1% | 0.1% | 0.1% | 1.02 (0.46, 2.28) | 0.53 (0.26, 1.08) | 1.25 (0.60, 2.62) | 1.13 (0.51, 2.54) | 1.37 (0.52, 3.60) | 1.68 (0.66, 4.30) |
| Physician Specialist | OS | 9.0% | 9.4% | 10.2% | 10.1% | 9.4% | 10.1% | 6.5% | 6.2% | 6.1% | 6.1% | 6.4% | 6.8% | 0.95 (0.85, 1.07) | 1.01 (0.92, 1.12) | 0.93 (0.84, 1.03) | 1.04 (0.92, 1.17) | 1.00 (0.89, 1.13) | 0.95 (0.85, 1.06) |
| | PMR | 3.8% | 5.0% | 3.6% | 3.6% | 3.6% | 3.9% | 3.2% | 2.9% | 2.7% | 2.4% | 2.9% | 2.7% | 0.76 (0.64, 0.90) | 0.98 (0.82, 1.17) | 0.93 (0.79, 1.11) | 1.11 (0.93, 1.32) | 1.12 (0.93, 1.35) | 1.06 (0.89, 1.26) |
| | PM | 2.4% | 1.5% | 1.7% | 2.4% | 1.9% | 1.6% | 2.5% | 3.0% | 2.4% | 2.2% | 2.3% | 2.4% | 1.55 (1.19, 2.02) | 0.72 (0.57, 0.91) | 1.15 (0.89, 1.48) | 0.82 (0.68, 0.99) | 1.06 (0.87, 1.30) | 0.93 (0.77, 1.12) |
| | Neuro | 0.9% | 0.6% | 0.9% | 0.8% | 0.8% | 0.6% | 0.9% | 0.8% | 0.8% | 1.0% | 0.7% | 0.8% | 1.51 (0.98, 2.33) | 1.10 (0.75, 1.59) | 1.38 (0.91, 2.09) | 1.03 (0.74, 1.44) | 0.80 (0.58, 1.10) | 0.83 (0.59, 1.15) |
| | Rheum | 0.9% | 0.4% | 0.9% | 0.4% | 1.1% | 0.2% | 1.9% | 0.7% | 1.8% | 0.6% | 1.3% | 0.7% | 2.32 (1.42, 3.77) | 2.42 (1.52, 3.85) | 5.31 (2.95, 9.54) | 2.60 (1.90, 3.56) | 3.30 (2.32, 4.68) | 1.94 (1.42, 2.66) |
| | NS | 0.3% | 0.3% | 0.3% | 0.2% | 0.2% | 0.4% | 0.1% | 0.2% | 0.2% | 0.2% | 0.1% | 0.2% | 1.12 (0.59, 2.10) | 1.39 (0.68, 2.87) | 0.58 (0.31, 1.07) | 0.53 (0.26, 1.08) | 1.22 (0.62, 2.38) | 0.63 (0.32, 1.24) |
| | MD Oth | 1.8% | 1.7% | 2.1% | 2.2% | 1.5% | 2.5% | 2.4% | 3.3% | 2.9% | 3.4% | 2.5% | 3.6% | 1.06 (0.81, 1.39) | 0.96 (0.76, 1.21) | 0.61 (0.48, 0.78) | 0.74 (0.62, 0.88) | 0.87 (0.73, 1.03) | 0.71 (0.60, 0.83) |
| Emergency Medicine/ Urgent Care | Hosp | 9.8% | 9.4% | 8.3% | 8.6% | 10.8% | 9.9% | 11.4% | 11.3% | 10.0% | 9.6% | 13.5% | 12.1% | 1.04 (0.93, 1.16) | 0.96 (0.86, 1.08) | 1.09 (0.99, 1.20) | 1.01 (0.92, 1.10) | 1.04 (0.95, 1.14) | 1.12 (1.04, 1.21) |
| | Rad | 9.5% | 8.3% | 10.1% | 9.1% | 13.2% | 11.9% | 12.9% | 11.3% | 11.9% | 12.0% | 14.5% | 13.5% | 1.14 (1.01, 1.28) | 1.11 (1.00, 1.23) | 1.10 (1.01, 1.20) | 1.13 (1.04, 1.23) | 0.99 (0.91, 1.07) | 1.07 (1.00, 1.15) |
| | EM | 2.9% | 3.2% | 2.3% | 2.7% | 2.6% | 3.1% | 2.1% | 2.0% | 1.4% | 1.7% | 1.6% | 1.9% | 0.90 (0.73, 1.10) | 0.83 (0.67, 1.03) | 0.85 (0.70, 1.03) | 1.05 (0.85, 1.30) | 0.83 (0.65, 1.06) | 0.86 (0.69, 1.07) |
| | UC | 1.1% | 1.3% | 1.3% | 1.2% | 1.1% | 0.9% | 1.0% | 0.7% | 0.9% | 0.9% | 0.8% | 0.8% | 0.86 (0.62, 1.20) | 1.05 (0.77, 1.42) | 1.21 (0.87, 1.70) | 1.42 (1.00, 2.02) | 1.02 (0.74, 1.42) | 0.94 (0.68, 1.30) |
| NHW 51-100 and ADI 51-100 - **White, Low Income** | | | | | | | | | | | | | | | | | | | |
| Episodes | | 8280 | 8202 | 10278 | 10508 | 9850 | 9564 | 31659 | 20912 | 35275 | 23747 | 39887 | 26521 | | | | | | |
| Non-Rx | DC | 32.6% | 36.3% | 32.7% | 36.3% | 30.6% | 34.8% | 10.5% | 14.8% | 9.3% | 13.5% | 10.2% | 14.0% | **0.90 (0.86, 0.94)** | **0.90 (0.87, 0.94)** | **0.88 (0.84, 0.91)** | **0.71 (0.68, 0.74)** | **0.69 (0.66, 0.72)** | **0.72 (0.69, 0.75)** |
| | PT | 1.9% | 1.8% | 1.7% | 1.3% | 1.8% | 1.6% | 2.6% | 2.3% | 2.1% | 2.0% | 2.7% | 2.3% | 1.04 (0.83, 1.29) | 1.23 (0.99, 1.54) | 1.13 (0.92, 1.40) | 1.16 (1.04, 1.29) | 1.09 (0.97, 1.22) | 1.16 (1.05, 1.28) |
| | LAc | 0.0% | 0.0% | 0.0% | 0.0% | 0.0% | 0.0% | 0.0% | 0.0% | 0.0% | 0.0% | 0.0% | 0.0% | 1.49 (0.25, 8.89) | 3.07 (0.32, 29.48) | 1.46 (0.24, 8.71) | 0.22 (0.02, 2.12) | 0.22 (0.05, 1.11) | 1.77 (0.47, 6.68) |
| Primary Care | PCP | 20.2% | 19.8% | 21.6% | 20.3% | 18.9% | 19.1% | 29.5% | 29.4% | 32.4% | 31.7% | 26.9% | 27.4% | 1.02 (0.96, 1.08) | 1.07 (1.01, 1.12) | 0.99 (0.93, 1.05) | 1.01 (0.98, 1.03) | 1.02 (1.00, 1.05) | 0.98 (0.96, 1.01) |
| | Nurse | 9.2% | 8.9% | 9.8% | 8.6% | 10.0% | 8.7% | 8.0% | 7.1% | 8.6% | 7.8% | 8.1% | 6.9% | 1.04 (0.94, 1.14) | 1.14 (1.05, 1.24) | 1.15 (1.05, 1.26) | 1.12 (1.05, 1.19) | 1.11 (1.05, 1.17) | 1.17 (1.11, 1.24) |
| | PA | 5.0% | 5.3% | 4.5% | 5.3% | 4.9% | 5.5% | 4.2% | 4.3% | 4.5% | 4.4% | 4.6% | 4.7% | 0.95 (0.83, 1.08) | 0.86 (0.76, 0.97) | 0.89 (0.79, 1.00) | 0.97 (0.90, 1.06) | 1.04 (0.96, 1.12) | 0.98 (0.91, 1.05) |
| | DO | 0.2% | 0.1% | 0.1% | 0.2% | 0.2% | 0.1% | 0.1% | 0.1% | 0.1% | 0.1% | 0.1% | 0.2% | 1.68 (0.77, 3.68) | 0.60 (0.30, 1.20) | 1.62 (0.79, 3.31) | 1.10 (0.64, 1.89) | 0.87 (0.53, 1.42) | 0.78 (0.51, 1.17) |
| Physician Specialist | OS | 5.2% | 4.7% | 4.6% | 4.8% | 5.8% | 5.5% | 6.0% | 5.9% | 6.1% | 6.1% | 6.4% | 6.3% | 1.09 (0.96, 1.25) | 0.96 (0.85, 1.08) | 1.05 (0.93, 1.18) | 1.02 (0.95, 1.09) | 1.00 (0.94, 1.07) | 1.02 (0.96, 1.08) |
| | PMR | 1.3% | 1.2% | 1.1% | 1.3% | 1.0% | 1.3% | 1.8% | 1.9% | 1.6% | 1.5% | 1.6% | 1.7% | 1.06 (0.81, 1.40) | 0.90 (0.70, 1.15) | 0.81 (0.63, 1.05) | 0.99 (0.88, 1.13) | 1.05 (0.92, 1.19) | 0.94 (0.83, 1.06) |
| | PM | 1.4% | 1.1% | 1.4% | 1.1% | 1.5% | 1.2% | 3.1% | 2.8% | 2.6% | 2.6% | 2.5% | 2.5% | 1.30 (0.99, 1.71) | 1.25 (0.98, 1.60) | 1.21 (0.95, 1.54) | 1.08 (0.97, 1.19) | 0.99 (0.90, 1.10) | 0.98 (0.89, 1.08) |
| | Neuro | 0.4% | 0.2% | 0.4% | 0.2% | 0.3% | 0.2% | 0.5% | 0.4% | 0.5% | 0.5% | 0.5% | 0.4% | 2.29 (1.19, 4.38) | 1.57 (0.96, 2.58) | 1.44 (0.85, 2.43) | 1.17 (0.91, 1.51) | 1.01 (0.81, 1.27) | 1.14 (0.90, 1.44) |
| | Rheum | 0.6% | 0.2% | 0.5% | 0.2% | 0.6% | 0.2% | 0.7% | 0.5% | 0.8% | 0.3% | 0.6% | 0.3% | 3.68 (2.04, 6.63) | 3.13 (1.78, 5.50) | 3.18 (1.88, 5.39) | 1.45 (1.14, 1.84) | 2.47 (1.91, 3.20) | 1.93 (1.51, 2.47) |
| | NS | 0.3% | 0.3% | 0.4% | 0.3% | 0.2% | 0.3% | 0.3% | 0.4% | 0.2% | 0.4% | 0.2% | 0.4% | 0.91 (0.52, 1.60) | 1.28 (0.80, 2.03) | 0.77 (0.42, 1.40) | 0.80 (0.59, 1.07) | 0.60 (0.45, 0.80) | 0.68 (0.51, 0.90) |
| | MD Oth | 0.9% | 1.0% | 1.2% | 1.3% | 1.1% | 1.1% | 1.5% | 1.5% | 1.8% | 2.0% | 1.4% | 1.9% | 0.95 (0.70, 1.30) | 0.90 (0.71, 1.14) | 1.04 (0.80, 1.36) | 0.95 (0.82, 1.09) | 0.88 (0.78, 0.99) | 0.72 (0.64, 0.82) |
| Emergency Medicine/ Urgent Care | Hosp | 11.8% | 9.8% | 10.6% | 9.5% | 13.2% | 11.0% | 17.9% | 16.3% | 16.7% | 15.3% | 19.7% | 18.0% | 1.20 (1.10, 1.32) | 1.11 (1.02, 1.21) | 1.19 (1.11, 1.29) | 1.10 (1.06, 1.15) | 1.09 (1.05, 1.13) | 1.10 (1.06, 1.13) |
| | Rad | 6.1% | 5.3% | 6.2% | 5.7% | 6.7% | 5.4% | 11.1% | 10.0% | 10.7% | 9.7% | 12.3% | 10.8% | 1.15 (1.01, 1.30) | 1.10 (0.99, 1.23) | 1.24 (1.11, 1.39) | 1.11 (1.06, 1.17) | 1.10 (1.05, 1.16) | 1.14 (1.09, 1.19) |
| | EM | 1.8% | 2.8% | 2.4% | 2.9% | 2.3% | 3.0% | 1.8% | 2.0% | 1.7% | 1.9% | 1.8% | 1.9% | 0.63 (0.52, 0.78) | 0.83 (0.70, 0.98) | 0.78 (0.65, 0.92) | 0.92 (0.81, 1.04) | 0.89 (0.78, 1.00) | 0.93 (0.83, 1.04) |
| | UC | 0.9% | 1.0% | 0.7% | 0.7% | 0.6% | 0.8% | 0.3% | 0.3% | 0.3% | 0.3% | 0.4% | 0.3% | 0.90 (0.66, 1.22) | 1.05 (0.76, 1.46) | 0.83 (0.59, 1.16) | 1.15 (0.84, 1.57) | 0.93 (0.68, 1.27) | 1.21 (0.92, 1.59) |
| NHW 51-100 and ADI 0-50 - **White, High Income** | | | | | | | | | | | | | | | | | | | |
| Episodes | | 16563 | 15413 | 18213 | 17405 | 18252 | 17393 | 28717 | 19765 | 29861 | 21221 | 35026 | 24409 | | | | | | |
| Non-Rx | DC | 25.1% | 27.7% | 25.4% | 26.8% | 23.4% | 26.4% | 9.7% | 12.9% | 8.7% | 11.4% | 9.0% | 11.5% | **0.91 (0.87, 0.94)** | **0.95 (0.91, 0.98)** | **0.89 (0.85, 0.92)** | **0.75 (0.72, 0.79)** | **0.76 (0.73, 0.81)** | **0.78 (0.75, 0.82)** |
| | PT | 5.5% | 4.5% | 4.6% | 4.0% | 4.6% | 4.1% | 5.1% | 4.8% | 3.9% | 4.1% | 4.6% | 4.5% | 1.21 (1.10, 1.33) | 1.16 (1.05, 1.28) | 1.12 (1.01, 1.23) | 1.08 (0.99, 1.17) | 0.94 (0.86, 1.03) | 1.02 (0.95, 1.10) |
| | LAc | 0.5% | 0.3% | 0.2% | 0.2% | 0.3% | 0.2% | 0.1% | 0.1% | 0.1% | 0.1% | 0.2% | 0.1% | 1.74 (1.19, 2.56) | 1.25 (0.79, 1.99) | 1.81 (1.17, 2.80) | 1.47 (0.80, 2.71) | 1.21 (0.70, 2.10) | 1.15 (0.75, 1.76) |
| Primary Care | PCP | 17.2% | 18.2% | 19.3% | 19.7% | 16.8% | 17.4% | 29.1% | 28.4% | 32.8% | 31.9% | 27.0% | 26.6% | 0.94 (0.90, 0.99) | 0.98 (0.94, 1.02) | 0.96 (0.92, 1.01) | 1.02 (0.99, 1.05) | 1.03 (1.00, 1.05) | 1.02 (0.99, 1.04) |
| | Nurse | 4.1% | 4.0% | 4.2% | 4.1% | 4.3% | 4.0% | 5.1% | 4.4% | 5.6% | 4.7% | 5.4% | 4.3% | 1.03 (0.92, 1.14) | 1.04 (0.94, 1.15) | 1.09 (0.99, 1.20) | 1.15 (1.06, 1.25) | 1.18 (1.09, 1.27) | 1.25 (1.16, 1.35) |
| | PA | 4.7% | 5.3% | 4.6% | 5.0% | 5.0% | 5.0% | 4.8% | 4.7% | 4.8% | 4.5% | 4.8% | 5.0% | 0.88 (0.80, 0.97) | 0.91 (0.83, 0.99) | 0.99 (0.90, 1.08) | 1.03 (0.95, 1.12) | 1.06 (0.98, 1.15) | 0.96 (0.90, 1.04) |
| | DO | 0.2% | 0.3% | 0.2% | 0.3% | 0.2% | 0.2% | 0.1% | 0.2% | 0.1% | 0.1% | 0.1% | 0.1% | 0.91 (0.58, 1.41) | 0.77 (0.50, 1.18) | 1.33 (0.84, 2.13) | 0.91 (0.57, 1.45) | 1.11 (0.67, 1.83) | 1.09 (0.70, 1.70) |
| Physician Specialist | OS | 10.6% | 10.6% | 10.3% | 10.9% | 10.7% | 10.8% | 7.0% | 7.2% | 6.7% | 7.0% | 7.1% | 7.6% | 0.99 (0.93, 1.06) | 0.94 (0.89, 1.00) | 0.99 (0.94, 1.05) | 0.97 (0.91, 1.04) | 0.95 (0.89, 1.02) | 0.94 (0.88, 0.99) |
| | PMR | 4.1% | 4.0% | 3.6% | 3.7% | 3.5% | 3.6% | 2.6% | 2.8% | 2.4% | 2.5% | 2.5% | 2.4% | 1.04 (0.94, 1.16) | 0.98 (0.88, 1.10) | 0.96 (0.86, 1.07) | 0.92 (0.83, 1.03) | 0.98 (0.87, 1.09) | 1.04 (0.94, 1.15) |
| | PM | 1.9% | 2.1% | 2.0% | 2.0% | 1.7% | 2.1% | 2.8% | 2.9% | 2.7% | 2.7% | 2.5% | 2.6% | 0.89 (0.76, 1.04) | 0.99 (0.86, 1.14) | 0.85 (0.73, 0.98) | 0.99 (0.89, 1.10) | 1.02 (0.91, 1.13) | 0.94 (0.85, 1.04) |
| | Neuro | 0.7% | 0.5% | 0.6% | 0.4% | 0.5% | 0.3% | 0.4% | 0.5% | 0.6% | 0.5% | 0.5% | 0.4% | 1.32 (0.98, 1.76) | 1.49 (1.11, 2.00) | 1.80 (1.28, 2.51) | 0.87 (0.67, 1.13) | 1.25 (0.99, 1.59) | 1.10 (0.87, 1.40) |
| | Rheum | 0.6% | 0.2% | 0.7% | 0.3% | 0.7% | 0.3% | 0.9% | 0.3% | 0.9% | 0.4% | 0.8% | 0.4% | 2.50 (1.72, 3.62) | 2.47 (1.78, 3.42) | 2.46 (1.77, 3.41) | 2.61 (2.01, 3.38) | 2.25 (1.76, 2.88) | 1.95 (1.54, 2.45) |
| | NS | 0.3% | 0.4% | 0.3% | 0.4% | 0.4% | 0.5% | 0.2% | 0.4% | 0.2% | 0.3% | 0.3% | 0.3% | 0.84 (0.58, 1.21) | 0.84 (0.59, 1.19) | 0.67 (0.49, 0.91) | 0.56 (0.40, 0.77) | 0.69 (0.49, 0.98) | 0.96 (0.69, 1.32) |
| | MD Oth | 1.5% | 1.5% | 1.9% | 2.1% | 1.7% | 1.8% | 1.7% | 2.1% | 2.1% | 2.7% | 1.8% | 2.2% | 0.97 (0.81, 1.16) | 0.89 (0.77, 1.02) | 0.92 (0.79, 1.08) | 0.80 (0.70, 0.91) | 0.78 (0.69, 0.87) | 0.80 (0.72, 0.90) |
| Emergency Medicine/ Urgent Care | Hosp | 10.7% | 8.9% | 10.0% | 8.5% | 11.1% | 9.5% | 15.3% | 13.8% | 14.1% | 13.5% | 16.8% | 15.8% | 1.21 (1.13, 1.29) | 1.18 (1.11, 1.26) | 1.17 (1.10, 1.24) | 1.11 (1.06, 1.16) | 1.05 (1.00, 1.10) | 1.07 (1.03, 1.11) |
| | Rad | 9.7% | 8.3% | 9.6% | 8.5% | 12.1% | 10.6% | 12.7% | 12.3% | 12.4% | 11.8% | 14.4% | 13.8% | 1.17 (1.09, 1.25) | 1.13 (1.05, 1.20) | 1.14 (1.08, 1.21) | 1.03 (0.98, 1.08) | 1.05 (1.00, 1.11) | 1.04 (1.00, 1.08) |
| | EM | 2.0% | 2.5% | 1.8% | 2.5% | 2.2% | 2.3% | 1.6% | 1.7% | 1.5% | 1.4% | 1.7% | 1.7% | 0.78 (0.67, 0.90) | 0.74 (0.64, 0.85) | 0.93 (0.81, 1.07) | 0.98 (0.86, 1.13) | 1.03 (0.89, 1.20) | 0.99 (0.88, 1.12) |
| | UC | 0.8% | 0.8% | 0.7% | 0.7% | 0.8% | 0.9% | 0.6% | 0.5% | 0.4% | 0.4% | 0.5% | 0.5% | 1.03 (0.80, 1.32) | 0.97 (0.76, 1.25) | 0.89 (0.71, 1.12) | 1.22 (0.95, 1.56) | 0.97 (0.74, 1.26) | 0.97 (0.76, 1.23) |

Pre=Pre-COVID period, Early=first 12 months post-COVID, Late=13-24 months post-COVID

PCP=primary care physician, PA=physician assistant, DO=doctor of osteopathy, DC=doctor of chiropractic, PT=physical therapist, LAc=licensed acupuncturist, OS=orthopedic surgeon, PMR=physical medicine and rehabilitation, PM=pain management, Neuro=neurologist, Rheum=rheumatologist, NS=neurosurgeon, MD-Oth=other physician specialtists, Hosp=hospital, Rad=radiologist, EM=emergency medicine, UC=urgent care

For risk ratio, cells in red indicate the confidence interval includes 1 and likelihood of HCP being initially contacted is not different than the male reference for the same time period
