## Supplement - Table 3 - Services By ADI for "Impact of patient gender on low back pain management before and after the COVID-19 pandemic in commercially insured and Medicare Advantage cohorts. A retrospective cohort study"

### Supplement - Table 3 - Type of healthcare service provided by patient home address 5 digit zip code population measures of race/ethnicity and socioeconomic status

| | | % of Episodes Including Service | | | | | | | | | | | | Risk Ratio and 95% Confidence Interval Comparing Female to Male Baseline | | | | | |
|---|---|---|---|---|---|---|---|---|---|---|---|---|---|---|---|---|---|---|---|
| | | Commercial Insurance (CI) | | | | | | Medicare Advantage (MA) | | | | | | Commercial Insurance (CI) | | | Medicare Advantage (MA) | | |
| | | Pre-COVID | | Early COVID | | Late COVID | | Pre-COVID | | Early COVID | | Late COVID | | Pre-COVID | Early COVID | Late COVID | Pre-COVID | Early COVID | Late COVID |
| | | F | M | F | M | F | M | F | M | F | M | F | M | | | | | | |
| % of Population that is non-Hispanic white (NHW 0%-50%) and Area Deprivation Index (ADI 51-100) - **Non-White, Low Income** | | | | | | | | | | | | | | | | | | | |
| | At least one service | 3453 | 3208 | 4313 | 3972 | 4239 | 3739 | 14592 | 8275 | 15752 | 8998 | 17272 | 9870 | | | | | | |
| First Line | Manipulation - Chiropractic | 18.3% | 22.7% | 18.7% | 23.2% | 18.1% | 23.5% | 4.3% | 6.2% | 3.9% | 5.6% | 4.5% | 6.7% | 0.81 (0.73, 0.89) | 0.81 (0.74, 0.88) | 0.77 (0.71, 0.84) | 0.68 (0.61, 0.76) | 0.70 (0.63, 0.79) | 0.67 (0.60, 0.74) |
| | Active Care | 18.2% | 18.8% | 16.3% | 17.5% | 18.1% | 19.0% | 9.9% | 9.6% | 8.0% | 7.9% | 10.0% | 10.2% | 0.97 (0.87, 1.07) | 0.93 (0.84, 1.02) | 0.95 (0.87, 1.05) | 1.03 (0.94, 1.11) | 1.02 (0.93, 1.11) | 0.98 (0.91, 1.06) |
| | Manual Therapy | 10.0% | 9.6% | 8.2% | 8.5% | 9.5% | 8.8% | 4.1% | 4.2% | 3.4% | 3.1% | 4.0% | 3.8% | 1.05 (0.90, 1.21) | 0.97 (0.84, 1.12) | 1.08 (0.94, 1.24) | 0.97 (0.85, 1.10) | 1.10 (0.95, 1.26) | 1.04 (0.92, 1.17) |
| | Passive Therapy | 10.3% | 12.7% | 10.1% | 13.8% | 9.4% | 12.9% | 1.8% | 1.8% | 1.3% | 1.6% | 1.5% | 1.7% | 0.81 (0.71, 0.93) | 0.73 (0.65, 0.82) | 0.73 (0.64, 0.83) | 1.03 (0.84, 1.25) | 0.82 (0.67, 1.02) | 0.87 (0.72, 1.05) |
| | Manipulation - Osteopathic | 0.6% | 0.6% | 0.3% | 0.3% | 0.4% | 0.4% | 0.2% | 0.2% | 0.2% | 0.2% | 0.2% | 0.2% | 1.14 (0.61, 2.11) | 0.85 (0.39, 1.86) | 1.13 (0.56, 2.28) | 1.10 (0.60, 2.01) | 1.02 (0.53, 1.96) | 1.07 (0.57, 2.00) |
| | Acupuncture | 0.2% | 0.2% | 0.2% | 0.2% | 0.4% | 0.2% | 0.1% | 0.1% | 0.1% | 0.1% | 0.2% | 0.1% | 1.11 (0.34, 3.65) | 0.92 (0.37, 2.32) | 2.21 (0.86, 5.68) | 2.15 (0.80, 5.77) | 1.07 (0.45, 2.53) | 1.24 (0.63, 2.45) |
| Second Line | Imaging - Radiography | 38.7% | 40.9% | 37.7% | 37.7% | 41.0% | 40.9% | 36.5% | 35.0% | 32.8% | 32.7% | 36.9% | 36.5% | 0.95 (0.89, 1.00) | 1.00 (0.95, 1.06) | 1.00 (0.95, 1.06) | 1.04 (1.01, 1.08) | 1.00 (0.97, 1.04) | 1.01 (0.98, 1.05) |
| | Rx NSAID | 29.2% | 29.5% | 30.4% | 29.4% | 29.1% | 29.0% | 25.9% | 25.3% | 26.8% | 25.6% | 25.0% | 24.7% | 0.99 (0.92, 1.07) | 1.03 (0.97, 1.11) | 1.00 (0.94, 1.08) | 1.02 (0.98, 1.07) | 1.05 (1.00, 1.09) | 1.01 (0.97, 1.06) |
| | Rx Muscle Relaxant | 25.0% | 29.6% | 26.4% | 28.4% | 23.6% | 27.2% | 16.7% | 17.7% | 16.4% | 18.4% | 16.8% | 18.1% | 0.84 (0.78, 0.91) | 0.93 (0.87, 1.00) | 0.87 (0.80, 0.93) | 0.95 (0.89, 1.00) | 0.89 (0.84, 0.94) | 0.93 (0.88, 0.98) |
| | Rx Oral Steroid | 10.7% | 13.2% | 9.0% | 12.2% | 8.4% | 10.2% | 7.3% | 8.6% | 6.2% | 7.9% | 5.4% | 6.8% | 0.81 (0.71, 0.93) | 0.74 (0.65, 0.84) | 0.82 (0.71, 0.94) | 0.85 (0.78, 0.94) | 0.79 (0.72, 0.87) | 0.80 (0.73, 0.88) |
| | Imaging - MRI | 8.9% | 10.6% | 9.1% | 8.9% | 10.3% | 10.4% | 11.9% | 14.1% | 11.6% | 13.5% | 14.2% | 15.8% | 0.84 (0.72, 0.97) | 1.02 (0.89, 1.17) | 0.99 (0.87, 1.13) | 0.85 (0.79, 0.91) | 0.86 (0.81, 0.92) | 0.90 (0.85, 0.95) |
| | Rx Gabapentin | 1.9% | 2.1% | 1.8% | 2.4% | 1.8% | 2.4% | 2.3% | 2.5% | 2.4% | 2.1% | 1.8% | 1.9% | 0.90 (0.64, 1.26) | 0.76 (0.56, 1.02) | 0.76 (0.56, 1.02) | 0.90 (0.76, 1.07) | 1.14 (0.96, 1.36) | 0.96 (0.80, 1.15) |
| | Rx Other | 6.4% | 5.5% | 6.9% | 5.8% | 6.8% | 6.2% | 12.4% | 11.0% | 14.8% | 13.8% | 12.5% | 11.4% | 1.16 (0.96, 1.41) | 1.18 (1.00, 1.39) | 1.09 (0.92, 1.29) | 1.13 (1.05, 1.21) | 1.07 (1.01, 1.14) | 1.10 (1.03, 1.18) |
| Third Line | Rx Opioid | 11.1% | 12.4% | 10.6% | 11.3% | 10.2% | 10.0% | 18.6% | 20.3% | 17.8% | 19.1% | 16.9% | 17.7% | 0.89 (0.78, 1.01) | 0.94 (0.83, 1.07) | 1.02 (0.89, 1.16) | 0.92 (0.87, 0.97) | 0.93 (0.88, 0.99) | 0.95 (0.90, 1.00) |
| | Spinal Injection | 4.6% | 4.6% | 3.9% | 4.6% | 4.5% | 4.5% | 6.9% | 7.2% | 6.5% | 6.7% | 7.5% | 7.3% | 0.99 (0.80, 1.23) | 0.86 (0.70, 1.06) | 1.00 (0.81, 1.22) | 0.97 (0.88, 1.07) | 0.98 (0.89, 1.08) | 1.02 (0.93, 1.11) |
| | Imaging - CT | 3.4% | 4.2% | 3.0% | 4.3% | 4.2% | 4.3% | 7.0% | 7.4% | 6.7% | 7.0% | 7.5% | 8.6% | 0.82 (0.64, 1.04) | 0.69 (0.55, 0.87) | 0.99 (0.80, 1.22) | 0.96 (0.87, 1.05) | 0.95 (0.86, 1.04) | 0.87 (0.80, 0.94) |
| | Surgery | 0.4% | 0.9% | 0.4% | 0.5% | 0.5% | 0.6% | 0.2% | 0.8% | 0.3% | 0.6% | 0.3% | 0.6% | 0.40 (0.21, 0.77) | 0.83 (0.45, 1.55) | 0.88 (0.48, 1.61) | 0.30 (0.20, 0.45) | 0.51 (0.35, 0.75) | 0.44 (0.30, 0.65) |
| NHW 0-50 and ADI 0-50 - **Non-White, High Income** | | | | | | | | | | | | | | | | | | | |
| | At least one service | 6344 | 5480 | 6970 | 6131 | 7189 | 6362 | 10708 | 6432 | 10606 | 6706 | 11677 | 7371 | | | | | | |
| First Line | Manipulation - Chiropractic | 16.6% | 20.5% | 16.9% | 20.8% | 16.5% | 20.4% | 4.2% | 7.1% | 3.8% | 5.8% | 4.2% | 6.1% | 0.81 (0.75, 0.87) | 0.81 (0.76, 0.87) | 0.81 (0.76, 0.87) | 0.60 (0.53, 0.68) | 0.64 (0.56, 0.74) | 0.69 (0.61, 0.78) |
| | Active Care | 24.9% | 25.4% | 21.1% | 22.3% | 23.2% | 23.0% | 14.2% | 15.8% | 10.2% | 11.4% | 14.0% | 13.6% | 0.98 (0.92, 1.04) | 0.95 (0.89, 1.01) | 1.01 (0.95, 1.07) | 0.90 (0.83, 0.97) | 0.89 (0.82, 0.97) | 1.02 (0.95, 1.10) |
| | Manual Therapy | 16.9% | 16.0% | 14.0% | 13.7% | 14.8% | 13.8% | 8.5% | 9.5% | 5.7% | 6.3% | 7.1% | 7.2% | 1.05 (0.97, 1.14) | 1.02 (0.94, 1.12) | 1.07 (0.98, 1.16) | 0.90 (0.82, 1.00) | 0.91 (0.80, 1.02) | 0.98 (0.88, 1.08) |
| | Passive Therapy | 11.7% | 13.9% | 10.8% | 11.8% | 10.1% | 11.6% | 2.9% | 4.0% | 1.9% | 2.4% | 2.5% | 2.7% | 0.84 (0.77, 0.93) | 0.92 (0.83, 1.01) | 0.88 (0.80, 0.97) | 0.73 (0.62, 0.85) | 0.76 (0.62, 0.93) | 0.92 (0.77, 1.10) |
| | Manipulation - Osteopathic | 0.6% | 0.7% | 0.4% | 0.6% | 0.6% | 0.3% | 0.3% | 0.2% | 0.3% | 0.2% | 0.2% | 0.1% | 0.84 (0.55, 1.30) | 0.71 (0.44, 1.15) | 1.61 (0.96, 2.70) | 1.13 (0.61, 2.06) | 1.56 (0.82, 2.96) | 1.43 (0.71, 2.91) |
| | Acupuncture | 0.7% | 0.6% | 0.8% | 0.4% | 0.8% | 0.6% | 1.2% | 1.2% | 0.9% | 0.8% | 1.3% | 1.4% | 1.16 (0.75, 1.79) | 1.79 (1.13, 2.84) | 1.37 (0.92, 2.06) | 1.02 (0.78, 1.35) | 1.11 (0.80, 1.55) | 0.94 (0.73, 1.21) |
| Second Line | Imaging - Radiography | 37.4% | 36.9% | 36.2% | 36.6% | 40.7% | 41.3% | 33.3% | 31.2% | 29.7% | 28.5% | 35.9% | 32.7% | 1.01 (0.97, 1.06) | 0.99 (0.94, 1.03) | 0.99 (0.95, 1.03) | 1.07 (1.02, 1.12) | 1.04 (0.99, 1.09) | 1.10 (1.05, 1.14) |
| | Rx NSAID | 29.2% | 30.2% | 30.3% | 30.7% | 27.4% | 27.4% | 27.9% | 29.0% | 28.9% | 27.1% | 26.4% | 26.0% | 0.97 (0.91, 1.02) | 0.99 (0.94, 1.04) | 1.00 (0.95, 1.06) | 0.96 (0.92, 1.01) | 1.06 (1.01, 1.12) | 1.01 (0.96, 1.06) |
| | Rx Muscle Relaxant | 22.7% | 25.2% | 22.5% | 23.7% | 20.6% | 22.1% | 13.5% | 15.1% | 14.2% | 14.2% | 13.8% | 15.5% | 0.90 (0.85, 0.96) | 0.95 (0.89, 1.01) | 0.93 (0.87, 0.99) | 0.90 (0.83, 0.97) | 1.00 (0.93, 1.08) | 0.89 (0.83, 0.96) |
| | Rx Oral Steroid | 8.6% | 12.1% | 7.7% | 9.9% | 6.9% | 8.8% | 7.1% | 8.3% | 5.3% | 7.1% | 4.6% | 6.1% | 0.71 (0.64, 0.79) | 0.78 (0.70, 0.88) | 0.78 (0.70, 0.88) | 0.85 (0.76, 0.95) | 0.74 (0.66, 0.83) | 0.75 (0.66, 0.85) |
| | Imaging - MRI | 10.0% | 11.6% | 9.8% | 11.5% | 12.2% | 13.4% | 13.6% | 14.8% | 13.3% | 14.7% | 16.1% | 18.1% | 0.86 (0.78, 0.96) | 0.85 (0.77, 0.94) | 0.91 (0.83, 0.99) | 0.92 (0.85, 0.99) | 0.90 (0.83, 0.97) | 0.89 (0.83, 0.95) |
| | Rx Gabapentin | 1.9% | 2.6% | 2.1% | 2.1% | 2.0% | 2.4% | 2.8% | 2.8% | 2.5% | 2.8% | 2.3% | 2.4% | 0.75 (0.59, 0.96) | 0.99 (0.79, 1.26) | 0.83 (0.67, 1.04) | 0.97 (0.81, 1.17) | 0.89 (0.74, 1.07) | 0.94 (0.78, 1.13) |
| | Rx Other | 6.5% | 6.7% | 7.3% | 6.4% | 6.7% | 6.9% | 14.7% | 13.2% | 17.8% | 16.7% | 14.1% | 14.0% | 0.97 (0.84, 1.11) | 1.15 (1.01, 1.30) | 0.98 (0.86, 1.11) | 1.11 (1.03, 1.20) | 1.07 (1.00, 1.14) | 1.01 (0.94, 1.08) |
| Third Line | Rx Opioid | 8.9% | 9.6% | 8.3% | 9.4% | 8.2% | 8.4% | 15.7% | 16.9% | 15.3% | 16.4% | 13.8% | 15.5% | 0.92 (0.83, 1.04) | 0.88 (0.79, 0.99) | 0.98 (0.88, 1.10) | 0.93 (0.86, 0.99) | 0.93 (0.87, 1.00) | 0.89 (0.83, 0.96) |
| | Spinal Injection | 4.8% | 5.8% | 4.7% | 5.6% | 4.9% | 5.4% | 6.8% | 7.0% | 6.2% | 6.6% | 6.9% | 7.6% | 0.83 (0.71, 0.97) | 0.83 (0.72, 0.97) | 0.90 (0.78, 1.04) | 0.98 (0.87, 1.10) | 0.94 (0.83, 1.05) | 0.91 (0.82, 1.01) |
| | Imaging - CT | 2.3% | 2.6% | 2.4% | 2.9% | 3.0% | 2.9% | 6.2% | 5.7% | 5.4% | 5.8% | 7.6% | 7.2% | 0.88 (0.70, 1.10) | 0.84 (0.69, 1.04) | 1.01 (0.83, 1.23) | 1.08 (0.95, 1.22) | 0.92 (0.82, 1.05) | 1.06 (0.95, 1.17) |
| | Surgery | 0.3% | 0.7% | 0.3% | 0.7% | 0.4% | 0.5% | 0.3% | 0.7% | 0.3% | 0.7% | 0.3% | 0.6% | 0.48 (0.28, 0.80) | 0.41 (0.24, 0.69) | 0.86 (0.51, 1.42) | 0.49 (0.32, 0.76) | 0.34 (0.21, 0.54) | 0.51 (0.33, 0.78) |
| NHW 51-100 and ADI 51-100 - **White, Low Income** | | | | | | | | | | | | | | | | | | | |
| | At least one service | 8280 | 8202 | 10278 | 10508 | 9850 | 9564 | 31659 | 20912 | 35275 | 23747 | 39887 | 26521 | | | | | | |
| First Line | Manipulation - Chiropractic | 34.0% | 37.8% | 33.9% | 37.6% | 32.2% | 36.5% | 11.7% | 16.5% | 10.3% | 15.0% | 11.6% | 15.9% | 0.90 (0.86, 0.94) | 0.90 (0.87, 0.94) | 0.88 (0.85, 0.92) | 0.71 (0.68, 0.74) | 0.69 (0.66, 0.72) | 0.73 (0.70, 0.76) |
| | Active Care | 16.2% | 16.0% | 15.3% | 15.0% | 16.2% | 15.5% | 10.4% | 10.0% | 9.0% | 9.0% | 11.1% | 10.3% | 1.01 (0.94, 1.08) | 1.02 (0.96, 1.09) | 1.04 (0.98, 1.11) | 1.05 (0.99, 1.10) | 1.00 (0.95, 1.05) | 1.07 (1.03, 1.12) |
| | Manual Therapy | 9.8% | 9.2% | 9.2% | 8.4% | 9.1% | 8.3% | 4.7% | 4.1% | 4.0% | 3.8% | 4.7% | 4.2% | 1.07 (0.97, 1.18) | 1.09 (1.00, 1.19) | 1.09 (1.00, 1.20) | 1.16 (1.07, 1.26) | 1.05 (0.97, 1.15) | 1.12 (1.04, 1.20) |
| | Passive Therapy | 13.7% | 16.1% | 13.4% | 14.6% | 11.8% | 14.1% | 1.8% | 1.8% | 1.3% | 1.6% | 1.5% | 1.4% | 0.85 (0.79, 0.92) | 0.92 (0.86, 0.98) | 0.84 (0.78, 0.90) | 1.03 (0.90, 1.17) | 0.86 (0.75, 0.99) | 1.04 (0.91, 1.18) |
| | Manipulation - Osteopathic | 0.6% | 0.7% | 0.3% | 0.4% | 0.5% | 0.5% | 0.4% | 0.3% | 0.2% | 0.2% | 0.3% | 0.2% | 0.83 (0.56, 1.22) | 0.81 (0.52, 1.27) | 0.91 (0.62, 1.35) | 1.38 (1.00, 1.90) | 1.08 (0.77, 1.52) | 1.15 (0.83, 1.58) |
| | Acupuncture | 0.2% | 0.1% | 0.2% | 0.1% | 0.2% | 0.1% | 0.0% | 0.0% | 0.1% | 0.1% | 0.1% | 0.1% | 1.49 (0.67, 3.31) | 2.17 (0.94, 5.03) | 1.50 (0.70, 3.20) | 0.75 (0.27, 2.08) | 0.75 (0.44, 1.26) | 1.60 (0.91, 2.82) |
| Second Line | Imaging - Radiography | 34.7% | 35.2% | 33.9% | 34.3% | 37.4% | 37.4% | 35.7% | 33.1% | 33.0% | 30.9% | 37.9% | 34.7% | 0.98 (0.94, 1.03) | 0.99 (0.95, 1.02) | 1.00 (0.97, 1.04) | 1.08 (1.05, 1.10) | 1.07 (1.04, 1.10) | 1.09 (1.07, 1.12) |
| | Rx NSAID | 20.4% | 20.9% | 20.2% | 19.6% | 20.8% | 20.3% | 18.3% | 16.7% | 18.4% | 17.1% | 17.0% | 15.7% | 0.97 (0.92, 1.03) | 1.03 (0.98, 1.09) | 1.02 (0.97, 1.08) | 1.10 (1.06, 1.14) | 1.07 (1.03, 1.11) | 1.08 (1.05, 1.12) |
| | Rx Muscle Relaxant | 19.7% | 23.3% | 20.1% | 22.6% | 19.3% | 21.9% | 12.6% | 13.8% | 12.8% | 13.1% | 13.0% | 14.1% | 0.85 (0.80, 0.90) | 0.89 (0.84, 0.94) | 0.88 (0.84, 0.93) | 0.91 (0.87, 0.95) | 0.98 (0.94, 1.02) | 0.92 (0.88, 0.95) |
| | Rx Oral Steroid | 11.1% | 12.7% | 9.1% | 12.5% | 8.6% | 11.1% | 8.5% | 10.2% | 7.4% | 8.9% | 6.9% | 7.9% | 0.87 (0.80, 0.95) | 0.73 (0.67, 0.79) | 0.77 (0.71, 0.84) | 0.83 (0.79, 0.88) | 0.84 (0.79, 0.88) | 0.87 (0.82, 0.92) |
| | Imaging - MRI | 8.3% | 9.4% | 7.4% | 9.0% | 8.8% | 9.9% | 12.9% | 14.2% | 12.5% | 14.2% | 15.4% | 17.1% | 0.88 (0.80, 0.97) | 0.83 (0.76, 0.91) | 0.88 (0.81, 0.97) | 0.91 (0.87, 0.95) | 0.88 (0.84, 0.91) | 0.90 (0.87, 0.93) |
| | Rx Gabapentin | 1.2% | 1.4% | 1.2% | 1.6% | 0.8% | 1.5% | 2.0% | 2.3% | 1.7% | 1.8% | 1.3% | 1.6% | 0.81 (0.62, 1.06) | 0.75 (0.59, 0.94) | 0.57 (0.44, 0.75) | 0.85 (0.76, 0.96) | 0.98 (0.87, 1.11) | 0.80 (0.71, 0.91) |
| | Rx Other | 6.5% | 5.5% | 7.2% | 6.0% | 6.6% | 5.9% | 12.0% | 11.7% | 14.3% | 13.4% | 11.7% | 11.3% | 1.19 (1.06, 1.35) | 1.20 (1.09, 1.33) | 1.11 (1.00, 1.24) | 1.03 (0.98, 1.08) | 1.07 (1.02, 1.11) | 1.03 (0.99, 1.08) |
| Third Line | Rx Opioid | 10.0% | 11.0% | 10.0% | 10.8% | 9.3% | 10.1% | 17.0% | 17.0% | 17.0% | 17.1% | 15.9% | 16.7% | 0.91 (0.83, 1.00) | 0.93 (0.86, 1.01) | 0.92 (0.85, 1.01) | 1.00 (0.96, 1.04) | 0.99 (0.96, 1.03) | 0.95 (0.92, 0.99) |
| | Spinal Injection | 4.3% | 4.5% | 4.1% | 4.3% | 4.1% | 5.1% | 8.5% | 8.7% | 7.9% | 8.1% | 9.1% | 9.2% | 0.94 (0.81, 1.08) | 0.95 (0.83, 1.08) | 0.81 (0.71, 0.92) | 0.98 (0.92, 1.03) | 0.97 (0.91, 1.02) | 0.99 (0.95, 1.04) |
| | Imaging - CT | 2.7% | 3.1% | 2.4% | 3.2% | 3.0% | 3.3% | 6.4% | 6.5% | 6.7% | 6.3% | 7.3% | 7.3% | 0.88 (0.74, 1.05) | 0.75 (0.64, 0.88) | 0.90 (0.77, 1.05) | 0.98 (0.92, 1.05) | 1.06 (1.00, 1.13) | 1.01 (0.95, 1.06) |
| | Surgery | 0.7% | 0.9% | 0.4% | 1.0% | 0.2% | 0.7% | 0.5% | 1.0% | 0.5% | 1.0% | 0.4% | 0.8% | 0.73 (0.51, 1.03) | 0.42 (0.29, 0.60) | 0.32 (0.20, 0.52) | 0.50 (0.40, 0.62) | 0.46 (0.38, 0.56) | 0.50 (0.41, 0.61) |
| NHW 51-100 and ADI 0-50 - **White, High Income** | | | | | | | | | | | | | | | | | | | |
| | At least one service | 16563 | 15413 | 18213 | 17405 | 18252 | 17393 | 28717 | 19765 | 29861 | 21221 | 35026 | 24409 | | | | | | |
| First Line | Manipulation - Chiropractic | 25.1% | 28.1% | 25.5% | 26.9% | 23.7% | 26.8% | 11.0% | 14.6% | 9.9% | 12.8% | 10.5% | 13.2% | **0.89 (0.86, 0.93)** | **0.95 (0.91, 0.98)** | **0.88 (0.85, 0.92)** | **0.75 (0.72, 0.79)** | **0.77 (0.73, 0.81)** | **0.79 (0.76, 0.83)** |
| | Active Care | 26.3% | 25.9% | 23.3% | 24.0% | 24.5% | 25.1% | 16.7% | 16.5% | 13.5% | 14.1% | 16.6% | 16.8% | 1.01 (0.98, 1.05) | 0.97 (0.93, 1.01) | 0.98 (0.94, 1.01) | 1.01 (0.97, 1.05) | 0.96 (0.92, 1.00) | 0.99 (0.95, 1.02) |
| | Manual Therapy | 17.8% | 16.8% | 14.6% | 14.8% | 15.4% | 14.8% | 9.0% | 8.6% | 7.1% | 7.1% | 8.6% | 8.2% | 1.06 (1.01, 1.11) | 0.99 (0.94, 1.04) | 1.04 (0.99, 1.09) | 1.04 (0.98, 1.10) | 1.00 (0.94, 1.07) | 1.04 (0.98, 1.10) |
| | Passive Therapy | 13.4% | 14.7% | 11.5% | 12.7% | 10.6% | 12.1% | 2.6% | 2.6% | 2.0% | 2.1% | 2.0% | 2.1% | 0.91 (0.86, 0.96) | 0.91 (0.86, 0.96) | 0.88 (0.83, 0.93) | 1.00 (0.90, 1.12) | 0.97 (0.86, 1.10) | 0.94 (0.84, 1.05) |
| | Manipulation - Osteopathic | 0.8% | 0.9% | 0.9% | 0.7% | 0.7% | 0.6% | 0.5% | 0.3% | 0.4% | 0.4% | 0.3% | 0.3% | 0.90 (0.71, 1.15) | 1.27 (1.01, 1.60) | 1.23 (0.95, 1.58) | 1.39 (1.04, 1.85) | 1.05 (0.79, 1.39) | 0.97 (0.73, 1.29) |
| | Acupuncture | 0.9% | 0.6% | 0.5% | 0.4% | 0.7% | 0.5% | 0.3% | 0.2% | 0.3% | 0.2% | 0.4% | 0.3% | 1.50 (1.15, 1.96) | 1.31 (0.96, 1.80) | 1.44 (1.08, 1.91) | 1.39 (0.95, 2.03) | 1.31 (0.91, 1.89) | 1.22 (0.93, 1.60) |
| Second Line | Imaging - Radiography | 38.4% | 38.1% | 36.5% | 36.4% | 40.3% | 39.8% | 35.3% | 32.9% | 32.2% | 30.7% | 38.0% | 35.6% | 1.01 (0.98, 1.04) | 1.00 (0.98, 1.03) | 1.01 (0.99, 1.04) | 1.07 (1.04, 1.10) | 1.05 (1.02, 1.08) | 1.07 (1.05, 1.09) |
| | Rx NSAID | 18.8% | 19.8% | 19.9% | 20.1% | 19.7% | 19.4% | 17.9% | 16.7% | 18.1% | 16.8% | 17.0% | 16.0% | 0.95 (0.91, 0.99) | 0.99 (0.95, 1.03) | 1.01 (0.97, 1.06) | 1.08 (1.03, 1.12) | 1.07 (1.03, 1.12) | 1.06 (1.03, 1.10) |
| | Rx Muscle Relaxant | 16.5% | 20.2% | 16.8% | 20.2% | 15.7% | 18.0% | 10.9% | 11.8% | 11.0% | 12.2% | 10.9% | 12.6% | **0.81 (0.78, 0.85)** | **0.83 (0.80, 0.87)** | **0.87 (0.83, 0.91)** | **0.92 (0.88, 0.97)** | **0.90 (0.86, 0.95)** | **0.87 (0.83, 0.91)** |
| | Rx Oral Steroid | 9.0% | 13.2% | 8.3% | 11.8% | 7.5% | 10.0% | 8.1% | 10.1% | 6.4% | 8.5% | 5.9% | 7.2% | **0.68 (0.64, 0.73)** | **0.71 (0.66, 0.75)** | **0.75 (0.70, 0.80)** | **0.80 (0.76, 0.85)** | **0.75 (0.70, 0.79)** | **0.82 (0.77, 0.87)** |
| | Imaging - MRI | 10.9% | 12.3% | 10.7% | 12.2% | 13.1% | 14.7% | 14.1% | 16.8% | 14.1% | 16.5% | 17.5% | 20.6% | **0.89 (0.84, 0.95)** | **0.88 (0.83, 0.93)** | **0.89 (0.85, 0.94)** | **0.84 (0.80, 0.88)** | **0.86 (0.82, 0.89)** | **0.85 (0.82, 0.88)** |
| | Rx Gabapentin | 1.1% | 1.5% | 1.4% | 1.9% | 1.4% | 1.7% | 2.4% | 2.6% | 2.3% | 2.5% | 1.8% | 1.9% | 0.75 (0.62, 0.90) | 0.72 (0.61, 0.85) | 0.82 (0.69, 0.97) | 0.94 (0.84, 1.05) | 0.94 (0.84, 1.05) | 0.95 (0.84, 1.07) |
| | Rx Other | 5.9% | 5.9% | 6.9% | 6.4% | 5.9% | 6.1% | 11.5% | 10.9% | 14.4% | 13.4% | 11.4% | 11.2% | 1.01 (0.93, 1.10) | 1.07 (0.99, 1.16) | 0.97 (0.89, 1.05) | 1.06 (1.00, 1.11) | 1.07 (1.03, 1.12) | 1.02 (0.98, 1.07) |
| Third Line | Rx Opioid | 7.7% | 9.0% | 8.3% | 9.4% | 7.6% | 8.6% | 14.8% | 14.7% | 15.4% | 14.6% | 13.7% | 13.8% | 0.85 (0.79, 0.92) | 0.88 (0.83, 0.95) | 0.88 (0.82, 0.94) | 1.01 (0.96, 1.05) | 1.05 (1.01, 1.10) | 1.00 (0.96, 1.04) |
| | Spinal Injection | 5.4% | 6.5% | 5.3% | 6.7% | 6.0% | 6.9% | 8.7% | 9.8% | 8.5% | 9.9% | 9.3% | 10.4% | **0.83 (0.76, 0.91)** | **0.79 (0.73, 0.86)** | **0.87 (0.81, 0.95)** | **0.89 (0.84, 0.95)** | **0.86 (0.81, 0.91)** | **0.90 (0.85, 0.94)** |
| | Imaging - CT | 2.0% | 1.9% | 2.0% | 2.2% | 2.2% | 2.6% | 5.5% | 5.0% | 5.6% | 5.6% | 6.3% | 6.2% | 1.05 (0.90, 1.23) | 0.89 (0.77, 1.02) | 0.86 (0.75, 0.98) | 1.09 (1.01, 1.17) | 1.00 (0.93, 1.08) | 1.00 (0.94, 1.07) |
| | Surgery | 0.4% | 0.9% | 0.6% | 0.9% | 0.4% | 0.8% | 0.5% | 1.1% | 0.5% | 1.0% | 0.3% | 1.0% | **0.47 (0.35, 0.63)** | **0.62 (0.49, 0.80)** | **0.51 (0.39, 0.68)** | **0.46 (0.37, 0.57)** | **0.50 (0.41, 0.62)** | **0.33 (0.27, 0.41)** |

Pre=Pre-COVID period, Early=first 12 months post-COVID, Late=13-24 months post-COVID

For risk ratio, cells in red indicate the confidence interval includes 1 and likelihood of HCP being initially contacted is not different than the male reference for the same time period
