## Supplement - Table 3 - Services By Type of HCP for "Impact of patient gender on low back pain management before and after the COVID-19 pandemic in commercially insured and Medicare Advantage cohorts. A retrospective cohort study"

**Supplement - Table 3 - Type of healthcare service provided by type of healthcare provider (HCP) initially contacted**

| | | % of Episodes Including Service | | | | | | | | | | | | Risk Ratio and 95% Confidence Interval Comparing Female to Male Baseline | | | | | |
|---|---|---|---|---|---|---|---|---|---|---|---|---|---|---|---|---|---|---|---|
| | | Commercial Insurance (CI) | | | | | | Medicare Advantage (MA) | | | | | | Commercial Insurance (CI) | | | Medicare Advantage (MA) | | |
| | | Pre-COVID | | Early COVID | | Late COVID | | Pre-COVID | | Early COVID | | Late COVID | | Pre-COVID | Early COVID | Late COVID | Pre-COVID | Early COVID | Late COVID |
| | | F | M | F | M | F | M | F | M | F | M | F | M | | | | | | |
| **Type of HCP Initially Contacted - Chiropractor (DC)** | | | | | | | | | | | | | | | | | | | |
| | At least one service | 8682 | 9185 | 10118 | 10761 | 9338 | 10158 | 7059 | 6506 | 6767 | 6393 | 8292 | 7480 | | | | | | |
| First Line | Manipulation - Chiropractic | 91.5% | 92.9% | 92.2% | 93.3% | 93.1% | 93.1% | 98.1% | 98.4% | 98.3% | 99.0% | 98.5% | 99.1% | 0.98 (0.98, 0.99) | 0.99 (0.98, 0.99) | 1.00 (0.99, 1.01) | 1.00 (0.99, 1.00) | 0.99 (0.99, 1.00) | 0.99 (0.99, 1.00) |
| | Active Care | 34.5% | 34.5% | 32.7% | 31.6% | 33.1% | 32.4% | 5.4% | 5.2% | 4.7% | 4.3% | 5.0% | 4.6% | 1.00 (0.96, 1.04) | 1.03 (0.99, 1.08) | 1.02 (0.98, 1.06) | 1.04 (0.91, 1.20) | 1.09 (0.93, 1.28) | 1.09 (0.95, 1.25) |
| | Manual Therapy | 17.4% | 16.6% | 15.9% | 15.2% | 14.3% | 13.9% | 3.3% | 3.0% | 3.0% | 2.5% | 3.0% | 3.0% | 1.04 (0.98, 1.11) | 1.05 (0.98, 1.11) | 1.03 (0.96, 1.10) | 1.10 (0.91, 1.33) | 1.16 (0.95, 1.42) | 0.97 (0.81, 1.16) |
| | Passive Therapy | 33.9% | 37.1% | 33.5% | 35.0% | 31.3% | 34.1% | 4.6% | 4.6% | 4.1% | 4.5% | 3.5% | 3.8% | 0.91 (0.88, 0.95) | 0.96 (0.92, 1.00) | 0.92 (0.88, 0.96) | 1.01 (0.87, 1.18) | 0.91 (0.78, 1.07) | 0.94 (0.80, 1.10) |
| | Manipulation - Osteopathic | 0.1% | 0.1% | 0.1% | 0.1% | 0.1% | 0.1% | 0.0% | 0.1% | 0.1% | 0.1% | 0.1% | 0.1% | 0.79 (0.28, 2.29) | 0.83 (0.31, 2.22) | 1.40 (0.52, 3.75) | 0.46 (0.08, 2.52) | 1.89 (0.65, 5.52) | 0.90 (0.32, 2.57) |
| | Acupuncture | 0.7% | 0.6% | 0.6% | 0.3% | 0.6% | 0.5% | 0.4% | 0.3% | 0.6% | 0.4% | 0.7% | 0.3% | 1.18 (0.81, 1.72) | 1.73 (1.14, 2.64) | 1.24 (0.85, 1.82) | 1.20 (0.67, 2.14) | 1.28 (0.79, 2.09) | 2.41 (1.46, 3.97) |
| Second Line | Imaging - Radiography | 17.6% | 20.8% | 18.9% | 21.0% | 19.5% | 21.5% | 7.2% | 7.1% | 7.6% | 7.7% | 8.0% | 7.9% | 0.85 (0.80, 0.90) | 0.90 (0.85, 0.95) | 0.90 (0.86, 0.96) | 1.01 (0.89, 1.14) | 0.99 (0.88, 1.11) | 1.01 (0.91, 1.12) |
| | Rx NSAID | 4.4% | 3.2% | 3.7% | 3.1% | 4.0% | 3.4% | 3.3% | 3.3% | 3.5% | 3.2% | 3.9% | 3.3% | 1.37 (1.18, 1.59) | 1.20 (1.04, 1.38) | 1.18 (1.02, 1.37) | 1.01 (0.84, 1.21) | 1.10 (0.92, 1.33) | 1.19 (1.01, 1.40) |
| | Rx Muscle Relaxant | 2.4% | 2.3% | 2.0% | 2.5% | 1.8% | 2.4% | 1.4% | 1.8% | 1.3% | 1.9% | 1.5% | 1.5% | 1.02 (0.84, 1.23) | 0.81 (0.68, 0.97) | 0.76 (0.63, 0.92) | 0.80 (0.61, 1.05) | 0.71 (0.54, 0.94) | 1.05 (0.82, 1.35) |
| | Rx Oral Steroid | 2.2% | 2.3% | 1.5% | 2.0% | 1.7% | 2.2% | 1.9% | 2.2% | 1.7% | 2.0% | 1.7% | 1.9% | 0.97 (0.80, 1.18) | 0.75 (0.61, 0.92) | 0.76 (0.62, 0.93) | 0.83 (0.66, 1.05) | 0.84 (0.66, 1.08) | 0.93 (0.74, 1.18) |
| | Imaging - MRI | 1.4% | 1.9% | 1.2% | 1.7% | 1.5% | 1.9% | 1.5% | 2.0% | 1.9% | 1.9% | 2.0% | 2.6% | 0.71 (0.56, 0.89) | 0.71 (0.57, 0.89) | 0.83 (0.67, 1.03) | 0.77 (0.59, 0.99) | 1.02 (0.79, 1.30) | 0.75 (0.61, 0.93) |
| | Rx Gabapentin | 0.1% | 0.2% | 0.1% | 0.2% | 0.2% | 0.2% | 0.3% | 0.4% | 0.2% | 0.3% | 0.2% | 0.3% | 0.50 (0.23, 1.11) | 0.53 (0.27, 1.06) | 0.90 (0.49, 1.65) | 0.69 (0.38, 1.27) | 0.52 (0.25, 1.08) | 0.53 (0.27, 1.02) |
| | Rx Other | 1.3% | 1.0% | 1.1% | 0.9% | 1.4% | 1.2% | 2.1% | 1.7% | 2.0% | 1.5% | 1.7% | 1.8% | 1.30 (0.99, 1.71) | 1.29 (0.98, 1.69) | 1.18 (0.92, 1.50) | 1.24 (0.98, 1.58) | 1.36 (1.05, 1.76) | 0.96 (0.76, 1.22) |
| Third Line | Rx Opioid | 2.6% | 2.1% | 2.7% | 2.3% | 2.5% | 2.2% | 3.1% | 3.6% | 3.2% | 3.5% | 3.1% | 3.6% | 1.25 (1.03, 1.51) | 1.18 (1.00, 1.40) | 1.16 (0.97, 1.39) | 0.86 (0.72, 1.03) | 0.92 (0.76, 1.10) | 0.86 (0.72, 1.01) |
| | Spinal Injection | 0.8% | 1.1% | 0.8% | 1.0% | 0.8% | 0.8% | 1.2% | 1.5% | 1.2% | 1.6% | 1.1% | 1.4% | 0.68 (0.50, 0.93) | 0.78 (0.59, 1.04) | 0.96 (0.71, 1.31) | 0.80 (0.60, 1.07) | 0.71 (0.53, 0.95) | 0.79 (0.60, 1.04) |
| | Imaging - CT | 0.3% | 0.2% | 0.2% | 0.3% | 0.1% | 0.3% | 0.6% | 0.5% | 0.5% | 0.5% | 0.8% | 0.9% | 1.22 (0.66, 2.26) | 0.60 (0.33, 1.09) | 0.45 (0.24, 0.83) | 1.11 (0.71, 1.73) | 0.89 (0.55, 1.43) | 0.94 (0.68, 1.32) |
| | Spinal Surgery | 0.1% | 0.2% | 0.1% | 0.2% | 0.1% | 0.2% | 0.0% | 0.1% | 0.1% | 0.2% | 0.0% | 0.1% | 0.73 (0.34, 1.57) | 0.56 (0.27, 1.15) | 0.29 (0.11, 0.77) | 0.18 (0.02, 1.58) | 0.71 (0.30, 1.68) | 0.45 (0.11, 1.80) |
| **Type of HCP Initially Contacted - Primary Care Provider (PCP)** | | | | | | | | | | | | | | | | | | | |
| | At least one service | 7071 | 6626 | 8992 | 8207 | 7860 | 7304 | 27246 | 17255 | 32374 | 20906 | 30894 | 19987 | | | | | | |
| First Line | Manipulation - Chiropractic | 3.8% | 4.4% | 3.8% | 3.8% | 3.6% | 4.3% | 1.6% | 2.1% | 1.4% | 1.9% | 1.8% | 2.4% | 0.87 (0.74, 1.03) | 0.99 (0.85, 1.14) | 0.85 (0.72, 0.99) | 0.74 (0.64, 0.85) | 0.72 (0.63, 0.82) | 0.76 (0.67, 0.86) |
| | Active Care | 11.4% | 11.2% | 9.2% | 10.1% | 11.1% | 11.4% | 6.7% | 7.4% | 5.0% | 5.8% | 7.2% | 7.6% | 1.02 (0.92, 1.12) | 0.91 (0.83, 0.99) | 0.97 (0.89, 1.06) | 0.90 (0.84, 0.97) | 0.86 (0.80, 0.92) | 0.94 (0.88, 1.00) |
| | Manual Therapy | 8.1% | 7.7% | 6.5% | 6.5% | 7.4% | 7.6% | 3.7% | 3.9% | 2.8% | 3.1% | 3.6% | 3.7% | 1.06 (0.94, 1.18) | 0.99 (0.88, 1.11) | 0.98 (0.88, 1.10) | 0.95 (0.86, 1.04) | 0.89 (0.80, 0.98) | 0.97 (0.88, 1.06) |
| | Passive Therapy | 5.0% | 5.2% | 3.5% | 4.1% | 3.6% | 4.1% | 1.4% | 1.5% | 1.0% | 1.2% | 1.1% | 1.3% | 0.95 (0.82, 1.10) | 0.87 (0.75, 1.01) | 0.87 (0.74, 1.02) | 0.88 (0.75, 1.03) | 0.85 (0.72, 1.01) | 0.89 (0.76, 1.05) |
| | Manipulation - Osteopathic | 1.9% | 2.1% | 1.6% | 1.5% | 1.5% | 1.4% | 0.7% | 0.6% | 0.5% | 0.5% | 0.5% | 0.5% | 0.90 (0.72, 1.14) | 1.10 (0.86, 1.39) | 1.08 (0.83, 1.41) | 1.22 (0.96, 1.54) | 1.07 (0.84, 1.37) | 0.97 (0.75, 1.25) |
| | Acupuncture | 0.4% | 0.2% | 0.2% | 0.2% | 0.2% | 0.1% | 0.1% | 0.1% | 0.1% | 0.1% | 0.2% | 0.2% | 2.13 (1.05, 4.32) | 0.97 (0.50, 1.87) | 1.76 (0.78, 3.94) | 1.61 (0.85, 3.05) | 1.23 (0.73, 2.09) | 1.00 (0.67, 1.49) |
| Second Line | Imaging - Radiography | 23.9% | 25.3% | 20.8% | 22.9% | 24.7% | 26.2% | 20.7% | 20.6% | 17.3% | 18.0% | 22.4% | 22.4% | 0.95 (0.89, 1.00) | 0.91 (0.86, 0.96) | 0.94 (0.89, 1.00) | 1.01 (0.97, 1.04) | 0.97 (0.93, 1.00) | 1.00 (0.97, 1.03) |
| | Rx NSAID | 43.7% | 43.3% | 44.2% | 41.6% | 43.1% | 41.9% | 33.2% | 31.7% | 32.0% | 30.6% | 32.7% | 30.6% | 1.01 (0.97, 1.05) | 1.06 (1.03, 1.10) | 1.03 (0.99, 1.07) | 1.05 (1.02, 1.08) | 1.04 (1.02, 1.07) | 1.07 (1.04, 1.09) |
| | Rx Muscle Relaxant | 38.3% | 44.6% | 38.1% | 43.2% | 35.6% | 40.7% | 19.8% | 21.5% | 19.1% | 20.7% | 20.5% | 23.3% | 0.86 (0.83, 0.90) | 0.88 (0.85, 0.91) | 0.87 (0.84, 0.91) | 0.92 (0.89, 0.96) | 0.92 (0.89, 0.95) | 0.88 (0.85, 0.91) |
| | Rx Oral Steroid | 16.8% | 22.6% | 14.5% | 20.2% | 13.3% | 17.9% | 11.2% | 13.8% | 8.8% | 11.2% | 8.8% | 10.5% | 0.74 (0.69, 0.80) | 0.72 (0.67, 0.77) | 0.74 (0.69, 0.80) | 0.82 (0.78, 0.86) | 0.79 (0.75, 0.83) | 0.84 (0.79, 0.88) |
| | Imaging - MRI | 5.5% | 7.3% | 4.8% | 6.5% | 5.8% | 7.7% | 6.1% | 7.5% | 5.5% | 6.9% | 7.5% | 9.1% | 0.75 (0.66, 0.86) | 0.73 (0.64, 0.82) | 0.75 (0.67, 0.85) | 0.82 (0.76, 0.87) | 0.80 (0.75, 0.86) | 0.82 (0.78, 0.87) |
| | Rx Gabapentin | 2.6% | 2.5% | 2.6% | 3.1% | 2.4% | 3.3% | 3.9% | 4.2% | 3.3% | 3.5% | 2.8% | 2.9% | 1.04 (0.84, 1.28) | 0.83 (0.70, 0.99) | 0.74 (0.61, 0.89) | 0.93 (0.84, 1.02) | 0.96 (0.88, 1.05) | 0.95 (0.86, 1.05) |
| | Rx Other | 13.9% | 12.2% | 14.8% | 13.4% | 14.2% | 13.8% | 23.8% | 23.1% | 27.3% | 26.4% | 24.3% | 23.7% | 1.13 (1.04, 1.24) | 1.11 (1.03, 1.19) | 1.03 (0.95, 1.11) | 1.03 (1.00, 1.07) | 1.03 (1.00, 1.06) | 1.02 (0.99, 1.06) |
| Third Line | Rx Opioid | 15.6% | 16.2% | 15.1% | 15.9% | 14.3% | 14.4% | 23.8% | 24.4% | 22.8% | 23.2% | 22.4% | 23.3% | 0.96 (0.89, 1.04) | 0.95 (0.89, 1.02) | 1.00 (0.92, 1.08) | 0.98 (0.94, 1.01) | 0.98 (0.95, 1.01) | 0.96 (0.93, 0.99) |
| | Spinal Injection | 2.6% | 3.6% | 2.6% | 3.4% | 3.2% | 3.7% | 3.6% | 3.9% | 3.2% | 3.5% | 4.0% | 4.4% | 0.70 (0.58, 0.85) | 0.75 (0.63, 0.89) | 0.85 (0.72, 1.01) | 0.91 (0.83, 1.01) | 0.90 (0.82, 0.99) | 0.93 (0.85, 1.01) |
| | Imaging - CT | 1.0% | 0.9% | 1.1% | 1.1% | 1.1% | 1.6% | 1.9% | 2.1% | 1.8% | 1.8% | 2.3% | 2.5% | 1.12 (0.80, 1.57) | 1.00 (0.76, 1.34) | 0.69 (0.52, 0.91) | 0.90 (0.79, 1.03) | 1.03 (0.91, 1.17) | 0.93 (0.83, 1.04) |
| | Spinal Surgery | 0.2% | 0.6% | 0.3% | 0.5% | 0.3% | 0.6% | 0.2% | 0.4% | 0.2% | 0.5% | 0.2% | 0.5% | 0.37 (0.21, 0.65) | 0.54 (0.33, 0.87) | 0.53 (0.33, 0.86) | 0.59 (0.42, 0.84) | 0.42 (0.31, 0.58) | 0.35 (0.25, 0.50) |
| **Type of HCP Initially Contacted - Orthopedic Surgeon (OS)** | | | | | | | | | | | | | | | | | | | |
| | At least one service | 2987 | 2744 | 3346 | 3302 | 3509 | 3299 | 5546 | 3569 | 5816 | 3918 | 7022 | 4673 | | | | | | |
| First Line | Manipulation - Chiropractic | 1.3% | 1.6% | 0.8% | 1.3% | 1.3% | 2.1% | 1.0% | 1.6% | 1.0% | 1.1% | 0.9% | 1.9% | 0.78 (0.51, 1.19) | 0.66 (0.41, 1.06) | 0.64 (0.44, 0.92) | 0.64 (0.45, 0.92) | 0.91 (0.61, 1.36) | 0.49 (0.35, 0.67) |
| | Active Care | 20.0% | 20.9% | 18.9% | 19.7% | 20.3% | 20.6% | 14.8% | 16.4% | 14.2% | 15.3% | 16.7% | 16.5% | 0.95 (0.86, 1.06) | 0.96 (0.87, 1.06) | 0.99 (0.90, 1.08) | 0.90 (0.82, 0.99) | 0.93 (0.84, 1.02) | 1.01 (0.93, 1.10) |
| | Manual Therapy | 13.6% | 14.6% | 13.3% | 13.5% | 13.8% | 13.2% | 7.1% | 7.4% | 6.9% | 7.2% | 8.0% | 7.5% | 0.93 (0.82, 1.06) | 0.98 (0.87, 1.11) | 1.05 (0.93, 1.19) | 0.96 (0.82, 1.11) | 0.96 (0.83, 1.11) | 1.07 (0.94, 1.21) |
| | Passive Therapy | 5.4% | 5.2% | 4.4% | 4.0% | 4.1% | 4.5% | 1.8% | 1.9% | 1.6% | 1.7% | 1.5% | 1.4% | 1.04 (0.83, 1.29) | 1.08 (0.86, 1.36) | 0.93 (0.74, 1.16) | 0.95 (0.70, 1.28) | 0.94 (0.69, 1.28) | 1.11 (0.81, 1.50) |
| | Manipulation - Osteopathic | 0.4% | 0.3% | 0.2% | 0.3% | 0.3% | 0.4% | 0.1% | 0.1% | 0.1% | 0.1% | 0.1% | 0.1% | 1.71 (0.68, 4.27) | 0.63 (0.24, 1.62) | 0.80 (0.36, 1.77) | 1.72 (0.46, 6.46) | 0.40 (0.10, 1.69) | 1.55 (0.40, 6.00) |
| | Acupuncture | 0.1% | 0.1% | 0.0% | 0.1% | 0.2% | 0.1% | 0.1% | 0.0% | 0.0% | 0.0% | 0.1% | 0.0% | 1.84 (0.34, 10.02) | nan | 1.88 (0.47, 7.51) | 3.22 (0.38, 27.53) | 1.35 (0.12, 14.85) | 5.32 (0.67, 42.55) |
| Second Line | Imaging - Radiography | 79.7% | 77.4% | 78.0% | 75.5% | 79.9% | 78.0% | 74.3% | 75.6% | 73.3% | 75.7% | 76.6% | 75.8% | 1.03 (1.00, 1.06) | 1.03 (1.01, 1.06) | 1.02 (1.00, 1.05) | 0.98 (0.96, 1.01) | 0.97 (0.95, 0.99) | 1.01 (0.99, 1.03) |
| | Rx NSAID | 21.7% | 25.2% | 24.4% | 25.8% | 26.3% | 27.0% | 21.8% | 20.6% | 22.1% | 20.2% | 20.7% | 20.7% | 0.86 (0.78, 0.94) | 0.95 (0.87, 1.03) | 0.97 (0.90, 1.05) | 1.06 (0.98, 1.15) | 1.09 (1.01, 1.18) | 1.00 (0.93, 1.08) |
| | Rx Muscle Relaxant | 10.6% | 15.2% | 13.2% | 15.5% | 11.4% | 13.1% | 5.4% | 5.8% | 6.1% | 6.2% | 5.9% | 6.8% | 0.70 (0.61, 0.80) | 0.85 (0.76, 0.96) | 0.87 (0.77, 0.99) | 0.92 (0.78, 1.10) | 0.99 (0.84, 1.16) | 0.87 (0.75, 1.00) |
| | Rx Oral Steroid | 11.1% | 16.2% | 11.1% | 15.0% | 8.6% | 11.3% | 8.3% | 10.5% | 7.3% | 9.4% | 6.2% | 8.5% | 0.68 (0.60, 0.78) | 0.74 (0.65, 0.84) | 0.76 (0.66, 0.88) | 0.79 (0.70, 0.90) | 0.77 (0.67, 0.88) | 0.72 (0.64, 0.83) |
| | Imaging - MRI | 15.4% | 18.1% | 15.9% | 18.4% | 17.0% | 21.3% | 19.6% | 22.4% | 20.4% | 23.1% | 23.5% | 25.3% | 0.85 (0.76, 0.95) | 0.86 (0.78, 0.96) | 0.79 (0.72, 0.88) | 0.87 (0.81, 0.95) | 0.88 (0.82, 0.95) | 0.93 (0.87, 0.99) |
| | Rx Gabapentin | 1.3% | 2.4% | 1.4% | 2.3% | 1.7% | 2.2% | 2.1% | 2.4% | 2.0% | 2.6% | 2.0% | 2.5% | 0.57 (0.38, 0.84) | 0.63 (0.44, 0.90) | 0.74 (0.52, 1.04) | 0.90 (0.68, 1.19) | 0.75 (0.58, 0.98) | 0.79 (0.62, 1.01) |
| | Rx Other | 3.9% | 3.9% | 3.5% | 4.1% | 3.7% | 4.2% | 4.5% | 4.2% | 4.9% | 4.3% | 5.0% | 4.8% | 1.00 (0.78, 1.30) | 0.86 (0.68, 1.10) | 0.88 (0.69, 1.11) | 1.06 (0.87, 1.30) | 1.13 (0.94, 1.36) | 1.04 (0.89, 1.23) |
| Third Line | Rx Opioid | 6.0% | 8.3% | 6.2% | 8.9% | 7.0% | 7.6% | 11.2% | 13.1% | 12.5% | 13.5% | 12.1% | 13.1% | 0.72 (0.60, 0.87) | 0.70 (0.59, 0.83) | 0.92 (0.78, 1.09) | 0.86 (0.77, 0.96) | 0.93 (0.83, 1.03) | 0.92 (0.83, 1.01) |
| | Spinal Injection | 7.6% | 8.6% | 6.7% | 9.4% | 7.2% | 10.0% | 13.1% | 16.0% | 13.5% | 15.8% | 15.1% | 15.6% | 0.89 (0.75, 1.06) | 0.71 (0.60, 0.84) | 0.71 (0.61, 0.83) | 0.82 (0.74, 0.91) | 0.85 (0.78, 0.94) | 0.96 (0.88, 1.05) |
| | Imaging - CT | 0.6% | 0.4% | 0.7% | 0.8% | 0.7% | 1.1% | 2.0% | 2.6% | 1.8% | 2.5% | 2.0% | 2.4% | 1.30 (0.62, 2.72) | 0.80 (0.46, 1.41) | 0.58 (0.35, 0.98) | 0.76 (0.58, 1.00) | 0.72 (0.55, 0.95) | 0.84 (0.66, 1.08) |
| | Spinal Surgery | 1.0% | 1.6% | 0.6% | 1.7% | 0.5% | 1.2% | 0.7% | 1.4% | 0.9% | 1.5% | 0.5% | 1.2% | 0.66 (0.42, 1.05) | 0.37 (0.22, 0.61) | 0.42 (0.24, 0.74) | 0.51 (0.34, 0.78) | 0.63 (0.43, 0.91) | 0.40 (0.26, 0.62) |
